# MIRS: an AI scoring system for predicting the prognosis and therapy of breast cancer

**DOI:** 10.1101/2021.12.16.21267775

**Authors:** Chen Huang, Min Deng, Dongliang Leng, Elaine Lai-Han Leung, Baoqing Sun, Peiyan Zheng, Xiaohua Douglas Zhang

## Abstract

Current scoring systems for prognosis of breast cancer are available but usually consider only one prognostic feature. We aim to develop a novel prognostic scoring system based on both immune-infiltration and metastatic features to not only assess the patient prognoses more accurately but also guide therapy for patients with breast cancer. Computational immune-infiltration and gene profiling analysis identified a 12-gene panel firstly characterizing immune-infiltrating and metastatic features. Neural network model yielded a precise prognostic scoring system called metastatic and immunogenomic risk score (MIRS). The influence of MIRS on the prognosis and therapy of breast cancer was then comprehensively investigated. MIRS significantly stratifies patients into high risk-group (MIRS^high^) and low risk-group (MIRS^low^) in both training and test cohorts. The MIRS^low^ patients exhibit significantly improved survival rate compared with MIRS^high^ patients. A series of analyses demonstrates that MIRS can well characterize the metastatic and immune landscape of breast cancer. Further analysis on the usage of MIRS in chemotherapy suggests that MIRS^high^ patients may benefit from three chemotherapeutic drugs (Cisplatin, Tamoxifen and Vincristine). Higher immune infiltration and significantly prolonged survival are observed in MIRS^low^ patients, indicating a better response in immune checkpoint inhibitor therapy. Our analysis demonstrates that MIRS could effectively improve the accuracy of prognosis for patients with breast cancer. Also, MIRS is a useful webtool, which is deposited at https://lva85.github.io/MIRS/, to help clinicians in designing personalized therapies for patients with breast cancer.

## Introduction

Cancer has long history in mankind and remains the leading cause of death, with breast cancer being one of the most common malignancies in women worldwide (1, 2). Breast cancer is also the second most common cause of death in cancer-related deaths among women. (3, 4). Despite tremendous advancement of medicine over the years has lowered the mortality rate, the high level of heterogeneity in breast cancer still makes the prognosis and treatment challenging.

Over the decade, a considerable amount of work has been done to develop prognostic measures on the progression of breast cancer (5). The majority (∼80%) of breast cancer becomes invasive (6) and approximately 20∼30% of them results in distant metastasis after treatment (7). Metastasis is thereby the most fatal development of breast cancer, which greatly reduces the rate of long-term survival from 90% to 5% (8). However, most metastasis-based signatures were developed based on organ-specific metastatic events, yet breast cancer consists of tumors with extremely heterogeneous cell types, resulting in the discrepancy between prognosis and survival (9, 10). Hence currently available metastasis-based prognostic measures have poor performance (11). On the other side, tumor-infiltrating lymphocytes have already been reported to be inextricably linked to therapeutic efficacy and patient survival in various cancers (12, 13). Many prognostic predictors were developed by assessing the level of the infiltration of immune cells into tumor and were preferably adopted for prognosis in cancers (14, 15). These histological strategies based on the analysis of a small proportions of immune cell marker genes support the prognostic significance of immune infiltration but still have limitations. Firstly, strategies for describing the level of immune infiltrate are the first limitation of the current studies (16). Specifically, each immune cell subset is computationally estimated by reference profiles based on bulk analysis of tissue samples. This is the main drawback because the transcriptional program of immunocytes exhibits high plasticity under tumor microenvironments (17). Secondly, while most studies were used the immune-related characteristics to improve cancer prognosis, only one or two subsets of immunocyte are included and these subsets lack functional variation, thus the treatments based on these indicators fail to achieve satisfactory immune response effects (18). Therefore, prognostic indicator based on only one characteristic without considering other crucial features is insufficient to accurately assess risk stratification and direct treatment strategies.

Given the limitations of the aforementioned work a more comprehensive approach should be developed to assess prognostic value and translate it into clinical practice. For the first time, we develop a prognostic signature for breast cancer patients, integrating immune-related gene signatures involved in metastasis, to classify patients with breast cancer into groups of high and low risk for potential therapeutic strategies. We construct a Neuron network to estimate gene weights, which exhibit outstanding performance in binary classification. A metastatic and immunogenomic risk score (MIRS) is then established, which has conspicuous power to predict survival status compared with previously published indicators based on single feature. Ultimately, the ability of MIRS to predict the treatment is identified, suggesting its potential to guide therapeutic tactics in breast cancer.

## Materials and Methods

### Collection and pre-processing of breast cancer data

All analyzed expression profiles and the corresponding clinical datasets were collected from Gene Expression Omnibus (GEO), The Cancer Genome Atlas (TCGA, https://www.cbioportal.org/datasets), and Molecular Taxonomy of Breast Cancer International Consortium (METABRIC, https://www.cbioportal.org/datasets). Only the datasets available with sufficient overall survival information were included, consisting of 8,424 patients from 14 cohorts. The detailed information of each cohort is presented in Table S1 and S2.

Raw series matrix files generated by Affymetrix were downloaded from GEO database. The R package GEOquery (19) was used to process raw matrix data. Duplicated genes detected by multiple probes were retained by taking the maximum expression value of the probe sets. Gene expression value was normalized by log_2_ transformation. Each GEO and RNA-seq dataset were processed independently.

### Construction of immune cell infiltration groups

A set of biomarkers is derived from Charoentong et al (20), comprising 45 immune signatures related to immune cell types, immunogenomic pathways and functions. The concrete gene signatures for each immune cell type were obtained from (21), and the immune-related pathways and functions were downloaded from database ‘ImmPort’ (22). Single sample gene set enrichment analysis (ssGSEA) implemented in R package GSVA was used to quantify the infiltration level of different immune cells, immunogenomic pathways and the activity of immune-related functions via expression data of breast cancer (23). Based on the results of ssGSEA, patients in TCGA breast cancer cohort (TCGA-BRCA) were divided into high and low immune cell infiltration groups using hierarchical clustering analysis (Figure S1) (24).

### Identification of immune and metastatic candidate genes

Using Wilcoxon rank-sum (Wilcoxon) test, the differentially expressed genes (DEGs) related to tumor immune infiltration were detected from high and low immune infiltration conditions according to the filtering criteria |log2FC| > 0.5 and adjusted p < 0.05 using Benjamini and Hochberg (BH) method (25). Meanwhile, utilizing the Wilcoxon test with the same criteria in the comparison between metastasis and primary breast cancer groups from the union of GSE10893 and GSE3521, the DEGs involved in metastatic mechanism were then identified. For these two DE analyses, Venn analysis found 52 metastatic and immunogenomic candidate genes. The heatmap of these DEGs are visualized in Supplementary Figures S3-S4.

### Establishment of prognostic risk score

Univariate Cox proportional hazard regression analysis was designed to screen features related to overall survival (OS) from 52 candidate genes in TCGA BRCA cohort. The filtered gene list is provided in Table S2. Subsequently, only the genes with absolute Hazard ratio (HR) larger than 1 and p-value less than 0.05 were retained. To eliminate collinearity, the eligible candidate genes were further filtered depending on the criteria that the square root of Variance Inflation Factor (VIF) was less than 2 and the Pearson Correlation Coefficient was smaller than 0.5. Ultimately, 12 prognostic genes that were significantly correlated with patients’ OS were identified.

These 12 prognostic signatures were classified into binary status. One was defined as the protective status in which HR was less than 1 whereas another was the dangerous status in which the corresponding HR was greater than 1. The expression status of each protective mRNA was assigned as 1 if the expression level of this mRNA was above the median of the expression values of all samples, otherwise it would be assigned as In contrast, the expression of dangerous mRNA was assigned as 1 if it had expression value below median, otherwise assigned as 0. This approach not only allows the risk score, which is based on protective and dangerous genes, to simultaneously contribute to consistent survival outcome, but also avoids the influence of inconsistent sequencing platforms. To date, several machine learning methods were found to be successful in various data mining problems, including those with transcriptomic data (26, 27). Therefore, a multilayer perceptron neuron network was built to estimate the weights of the 12 prognostic genes. In the Figure S4, the 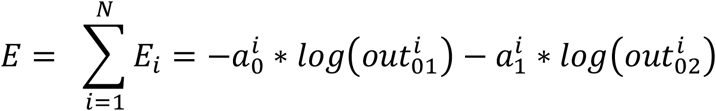 was defined, where *W* is the weight of each input node and *i*_*j*_ (*j* = 1,2,.., 12) is the ‘0-1’ status of gene. Then we exploited rectified linear unit (ReLU):

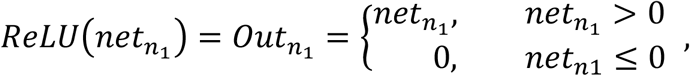

and 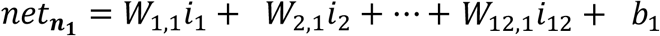 as an activation function in the hidden layer. In the output layer, we applied the Softmax function to each node and designated probability of death:

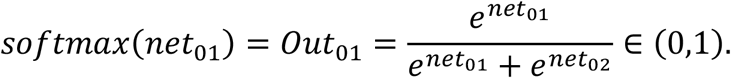

We then created two nodes *a*_0_ = 0 and *a*_1_ = 1 for alive and dead, respectively. Cross entropy error is computed as:

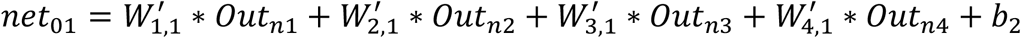, *where i is ith sample*.

Finally, the value of each weight was optimized by minimizing *E* using gradient descent. The R packages Tensorflow and Keras were employed to construct neuron network. After training, the coefficient of each prognostic gene was then determined as the maximum weight in the hidden layer (26).

Lastly, the risk score that consists of 12 metastatic and immunogenomic prognostic genes (MIRS) for each patient is defined as the following:

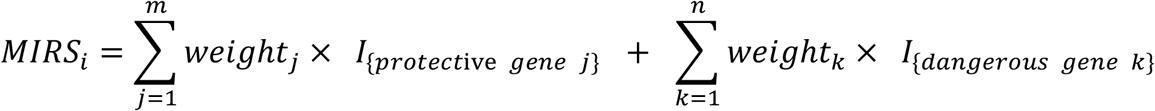

where *m* and *n* denote the number of protective and dangerous genes, respectively, *weight* is the maximum weight from the hidden layer. Additionally, *I*_{*protective gene j*}_ and *I*_{*dangerous gene k*}_ denote the following indicator functions:

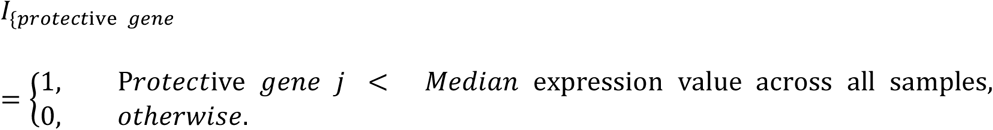

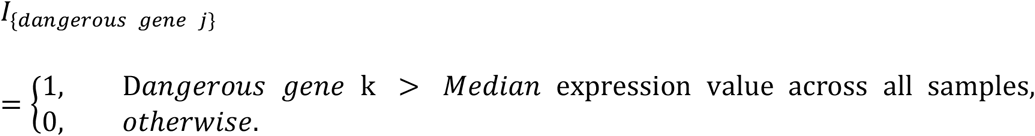

### Statistical analysis

All statistical analyses were performed using R software. The R packages pheatmap and ggplot2 were used to plot heatmap and other graphs. The R package forestplot was used to draw forest plot. The pROC package was employed to generate the Receiver Operating Characteristic (ROC) curve and calculate the Area Under Curve (AUC), which was an indicator to evaluate the predictive performance of risk score.

Based on the risk score, breast cancer patients in the investigated cohort were stratified into subtypes of high risk or low risk depending on whether the value of (*MIRS in each patient*)/(*median of MIRS in all patients*) was greater or less than 1. This stratification method allows reasonable comparisons between different data platforms. OS curves were established by Kaplan–Meier survival (KM) curve function ggsurvplot, as implemented in R package survminer, and the difference in survival distributions between risk subgroups was estimated by two-side log-rank test. Based on univariate Cox proportional hazard regression analysis, the targeted prognostic genes which were significantly correlated with OS were disclosed and the Hazard ratio (HR), 95% confident interval of HR and p-value were also evaluated. Multivariate Cox proportional hazard regression model was implemented to assess whether the risk score is an independent prognosis factor when compared with other important clinical features. All statistical tests were considered significant with p-value < 0.05.

Full details about data and methods descriptions, including data information, gene set enrichment analysis and mutation landscape analysis.

## Results

### Screening of candidate genes from three public datasets

To obtain significant prognostic biomarkers in breast cancer, we proposed a systematic scheme of bioinformatic analysis (Figure 1). Given that the processes of metastasis and immune infiltration in tumor play various important roles in cancer development, we hypothesize that the expression of genes which were associated with metastasis and immune infiltration in tumor should be correlated to the OS of cancer patients. We thereby identified prognostic signatures based on these two characteristics. Concretely, using ssGSEA method, the expression profile of 1,100 patients from TCGA cohort were used to construct groups of high and low immune cell infiltration. Then the patients were classified into the high immune infiltration group and low immune infiltration group (Figure 2A and Figure S1). Furthermore, to validate the reliability of the above grouping tactic, we investigated the expression level of two immune-related gene families between these two groups: *CD1* and *IL1*. As expected, the expression of both immune-related gene families in the high immune infiltration group is significantly higher than that in the low immune infiltration group (Figure 1B and Figure S5). Additionally, compared with low immune cell infiltration group, high immune cell infiltration group exhibits a higher fraction of immune cell, stromal cell but lower tumor purity using ESTIMATE (28) algorithm (Figure 2C). Furthermore, we found that high immune cell infiltration group had significantly higher proportions in most immune cell types than low immune infiltration group (Figure 2D) using CIBERSORT algorithm under the permutation test with 1000 times. These findings support that our immune cell infiltration grouping is highly confident to be used in downstream analyses. Next, 1,222 differentially expressed genes were identified via differential expression (DE) analysis between these two groups, which represents a high-confidence dataset of genes related to immune infiltration (Table S4).

**Figure 1.**
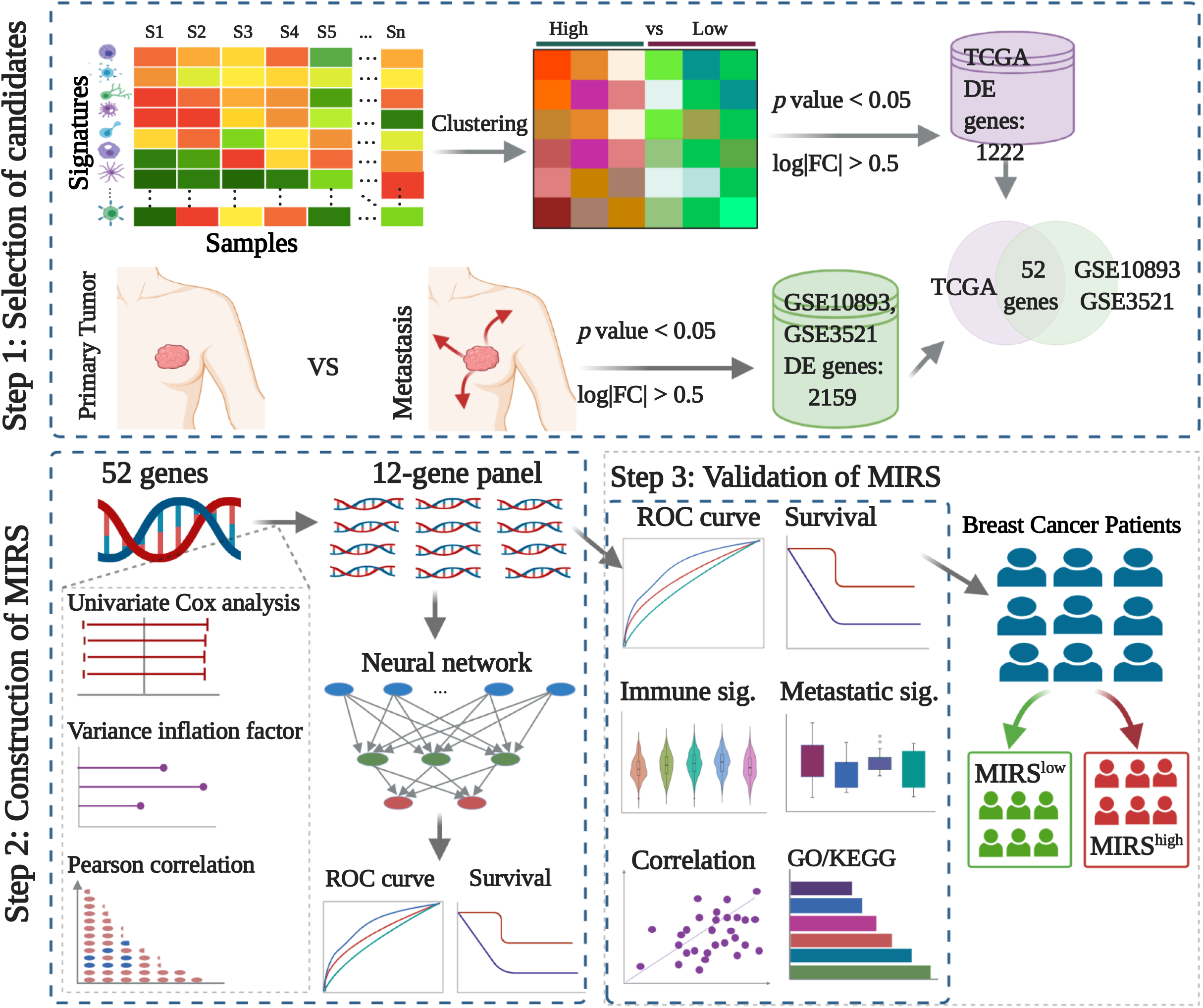
Systematic bioinformatic analysis pipeline.

**Figure 2.**
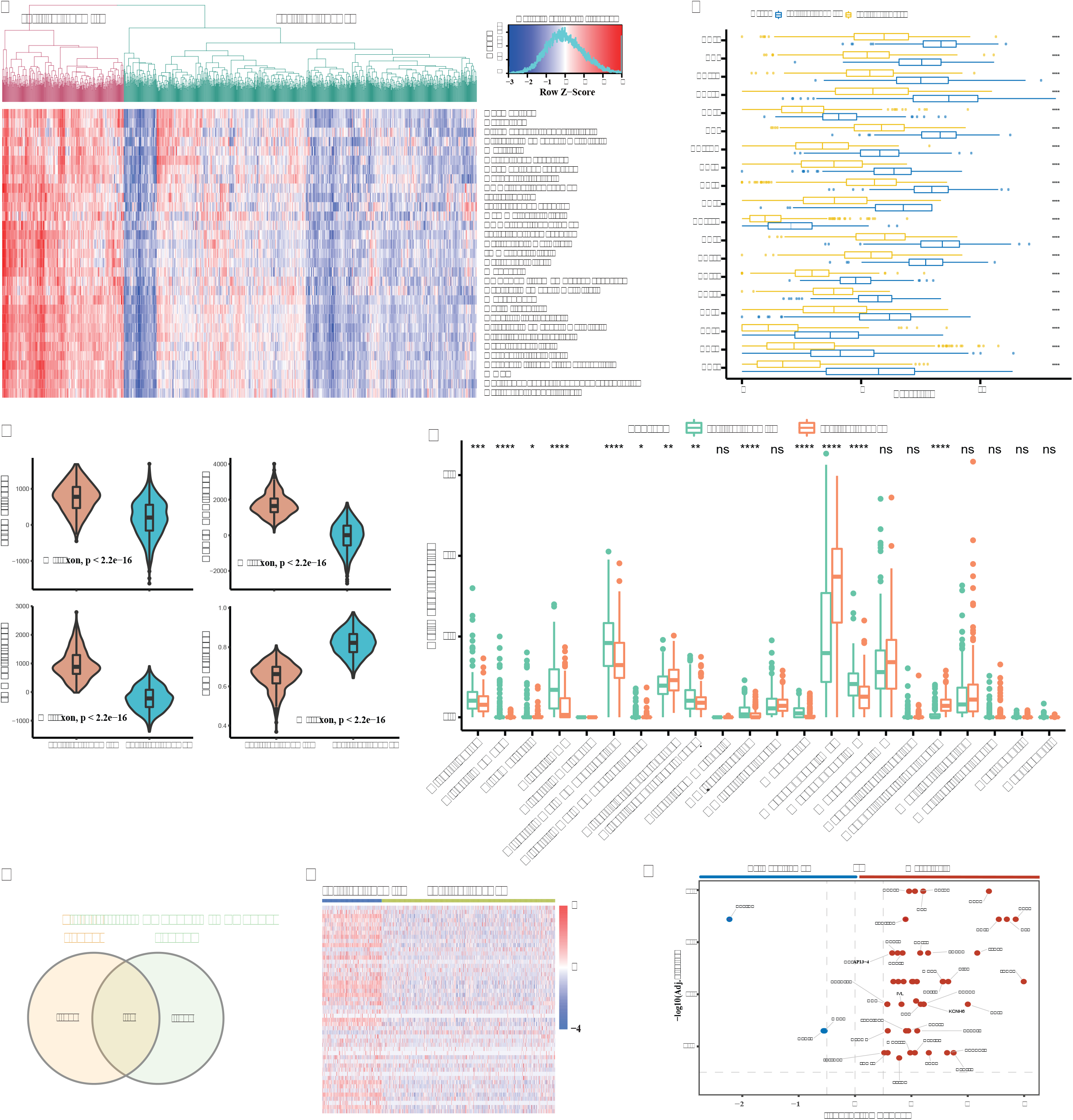
Exploration of the immune cell infiltration grouping, and 52 candidate genes were expressed in BRCA samples from the TCGA, GSE10893, and GSE3521 datasets. **(A)** Heatmap for the high and low immune-cell infiltration grouping from the TCGA cohort. **(B)** Boxplots for the expression levels of the CD family gene between high and low infiltration groups. **(C)**Comparison of Stromal score, Immunity score, ESTIMATE score and Tumor purity between the high and low immune infiltration groups. **(D)** Boxplots illustrate the 22 immune cell proportion s between high and low immune infiltration groups. **(E)** Venn plot of the differentially expressed genes from the TCGA data and GEO datasets. **(F)** Heatmap of the 52 candidate gene expression values between high and low immune infiltration groups from the TCGA dataset. Cluste1 represents the low immune infiltration level group, cluster 2 represents the high immune infiltration level group. **(G)** Volcano plot of the 52 candidate genes between the primary and metastasis tumor groups both from the GSE3521 and GSE10893. The blue dots show the DE genes are down regulated in the metastasis group. The red dots display the DE genes are up regulated in the metastasis group. The p-values were calculated using Wilcox rank sum test.

On the other part, aimed at identification of metastasis-related candidates, DE analysis between metastasis and primary patients with breast cancer were performed using two GEO cohorts (GSE10893 and GSE3521). The reason why we only chose these two GEO datasets is that they have relatively balanced sample sizes between the metastasis and primary groups when compared with other cohorts (Table S1). For instance, TCGA breast cancer cohort contains 1,165 primary individuals but only 23 metastatic individuals. There is no doubt that such an extremely imbalanced data would lead to biased result in DE analysis. This step yielded a union of 2,159 DE genes from the results of these two GEO datasets (Table S4). Finally, a total of 52 genes was obtained by intersecting 1,222 immune-infiltration-related genes and 2,159 metastasis-related genes (Figure 2E), which represents prognostic candidates associated with both tumor-immune infiltration (Figure 2F) and metastasis (Figure 2G).

### Construction and validation of MIRS in breast cancer cohorts

Univariate Cox regression analyses were performed to estimate the prognostic relationship between candidate genes and overall survival in TCGA cohort. Among these 52 candidate genes, 15 genes with p-value less than 0.05 were selected for follow-up study (Table S2). Given that too many redundant variables would result in overfitting in the linear model, we employed the analyses of Variance Inflation Factor and Pearson Correlation Coefficient to eliminate the redundant genes (Figure 3A and 3B). As a result, a panel of 12 genes is reserved to establish the predictive model.

**Figure 3.**
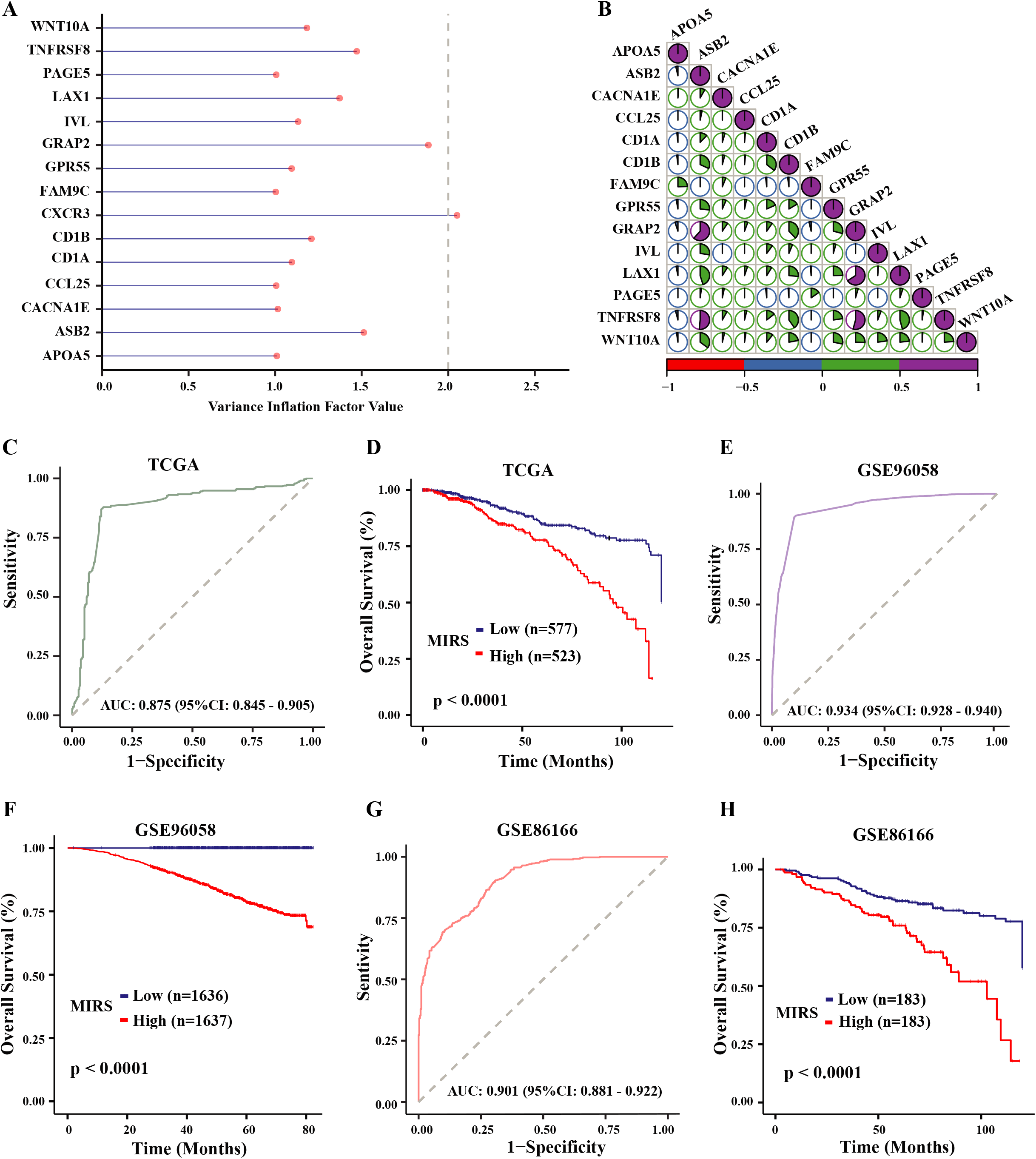
Construction and validation of the MIRS in the training and testing cohorts. **A**. The square root of the variance inflation factor value for each candidate gene in the training data. **B**. Correlations between the candidate genes in the training TCGA data. Different correlations between two genes are represented by different colors. **C**. ROC curve for the patient’s overall survival prediction in the training TCGA data. **D**. Kaplan-Meier curves of overall survival according to the MIRS subtypes in the training TCGA data. **E**. ROC curve for the patient’s overall survival prediction in GSE96058. **F**. Kaplan-Meier curves of overall survival according to the MIRS subtypes in GSE96058. **G**. ROC curve for the patient’s overall survival prediction in GSE86166. **H**. Kaplan-Meier curves of overall survival according to the MIRS subtypes in GSE86166.

The TCGA-BRCA data (N = 1100 patients) were randomly classified into training data (N = 770 patients) and testing data (N = 330 patients) at a ratio of 7:3. We then optimized the weights for each gene with Neuron network in the training TCGA data. The MIRS for each patient was built by summation of *Weight* × *I*_{*protective or dangerous gene*}_ of all 12 genes (Table 1). MIRS was initially used to predict patient’s survival status, which yielded great predictive performance with AUC accuracy of 0.875 in the training TCGA cohort (Figure 3C). In addition, all the patients were classified into MIRS^high^ group and MIRS^low^ group using the median value of MIRS as risk cut-off. As shown in Figure 3D, patients in MIRS^low^ group had significantly longer OS or disease-free survival (DFS) time than those in MIRS^high^ group (log-rank p<0.001) (Figure 3D and Figure S6-A).

**Table 1.** The 12 prognostic genes for calculating the risk score in TCGA data

| Gene ID | Category | Gene expression<br>(high) | Gene expression<br>(low) | Weight |
| --- | --- | --- | --- | --- |
| APOA5 | Dangerous | 1 | 0 | 0.4703 |
| FAM9C | Dangerous | 1 | 0 | 0.5585 |
| IVL | Dangerous | 1 | 0 | 0.4467 |
| PAGE5 | Dangerous | 1 | 0 | 0.5637 |
| CACNA1E | Protective | 0 | 1 | 0.3596 |
| CCL25 | Protective | 0 | 1 | 0.5013 |
| CD1A | Protective | 0 | 1 | 0.1782 |
| CD1B | Protective | 0 | 1 | 0.7733 |
| GPR55 | Protective | 0 | 1 | 0.6999 |
| **LAX1** | **Protective** | **0** | **1** | **0.6383** |
| **TNFRSF8** | **Protective** | **0** | **1** | **0.6234** |
| **WNT10A** | **Protective** | **0** | **1** | **0.4189** |

To further examine the robustness and feasibility of this MIRS model, a comprehensive survival analysis with KM method was performed in three independent testing cohorts. Notably, MIRS exhibited robust predictive capacity with AUC of 0.934, 0.901, and 0.904 in GSE96058, GSE86166 and GSE20685, respectively (Figure 3E and 3G, Supplementary Figure S6-C). Regarding the survival analyses, consistent with the result of the training data, the patients who are divided into MIRS^high^ group have significantly worse OS than those in MIRS^low^ group (Figure 3F, Figure 3H and Supplementary Figure S6-B). These analyses indicated that MIRS had precisely prognostic ability in breast cancer. The higher score of MIRS corresponds to poor outcome, and the lower score of MIRS refers to favorable outcome.

### Correlation of MIRS with the metastatic and immunogenomic landscape between the high and low subtypes

We want to further scrutinize the correlation of metastatic and immunogenomic landscape with MIRS in breast cancer patients. Initially, we investigated the correlation between MIRS and the fraction of immune cell, stromal cell, as well as tumor purity via ESTMATE in the GSE86166 cohort. The results showed that MIRS^low^ group had a higher fraction of immune cell and stromal cell cell but a lower tumor purity (Figure 4A). Similar situations were observed in GSE96058 (Figure S7). Reasonably, a higher fraction of immune cell and lower tumor purity reflects a high level of infiltrating T-lymphocytes in the patients of MIRS^low^ group, which is consistent with previous survival analysis.

**Figure 4.**
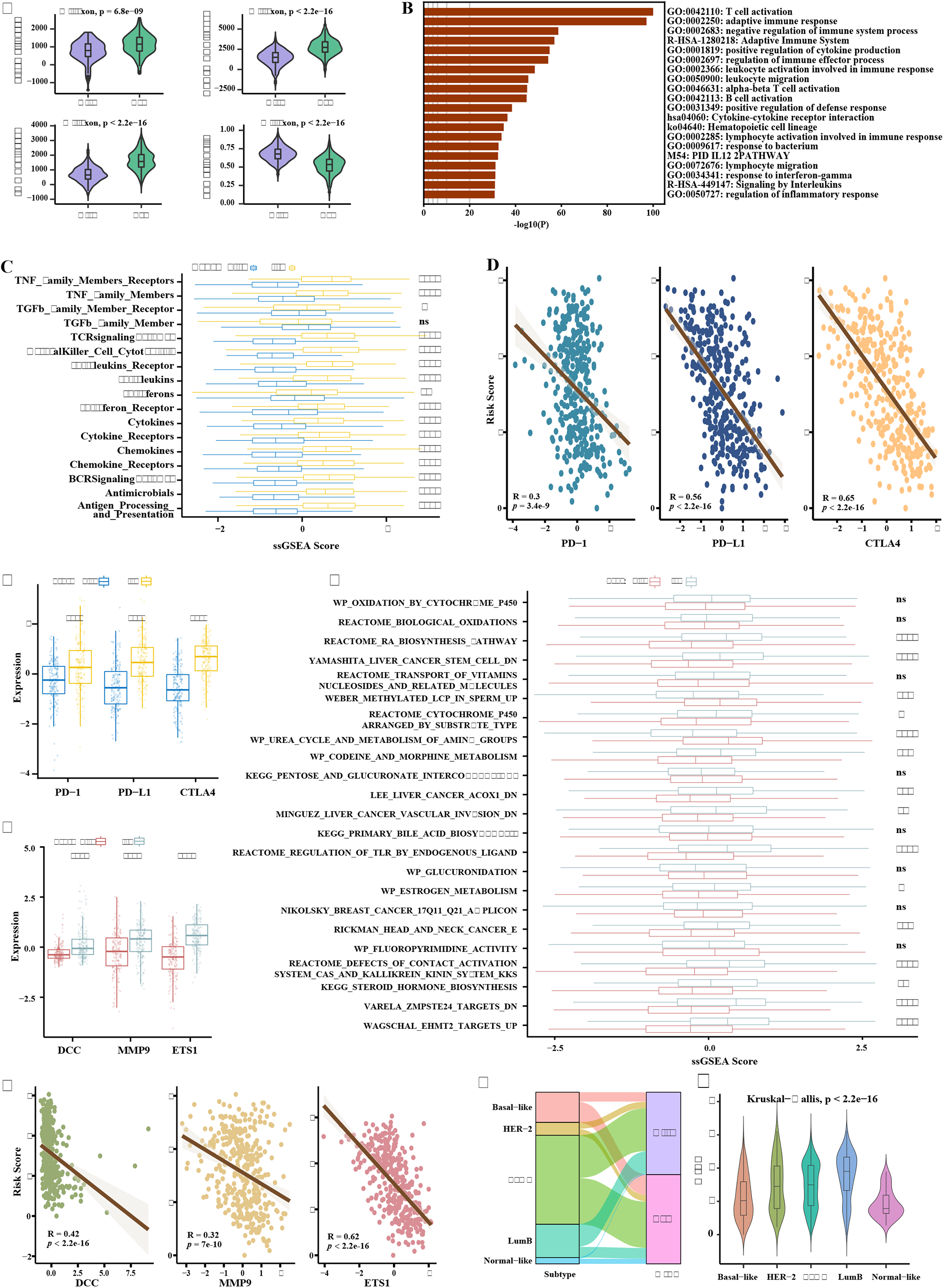
Correlation of MIRS with the metastatic and immunogenomic landscape between the high and low MIRS subtypes. **A**. Comparison of the Stromal score, ESTIMATE score, Immune score, and Tumor purity between high and low MIRS subtypes in GSE86166. The p-values were calculated using Wilcoxon rank sum test. *p<0.05; **p<0.01; ***p<0.0001. **B**. Function enrichment bar plot for the genes in GSE86166 which were highly correlated (Spearman correlation coefficient≥ 0.04) with 12 prognostic genes in GSE86166. **C**. Boxplots of the ssGSEA score for 17 immune-related biological functions and pathways between two MIRS subtypes in the GSE86166. The p-values were calculated using Wilcoxon rank sum test. *p<0.05; **p<0.01; ***p<0.0001. **D**. The spearman correlation between the gene expression levels of PD-1, PD-L1 and CTLA4 and MIRS score in the GSE86166 data, respectively. **E**. The boxplots of PD-1, PD-L1 and CTLA4 for two MIRS subtypes in the GSE86166 data. The p-values were calculated using Wilcoxon rank sum test. *p<0.05; **p<0.01; ***p<0.0001. **F**. The boxplots of DCC, MMP9 and ETS1 for two MIRS subtypes in GSE86166 dataset. The p-values were calculated using Wilcoxon rank sum test. *p<0.05; **p<0.01; ***p<0.0001. **G**. Boxplots of the ssGSEA score for 23 metastatic biological functions and pathways between two MIRS subtypes in the GSE86166. The p-values were calculated using Wilcoxon rank sum test. *p<0.05; **p<0.01; ***p<0.0001. **H**. The spearman correlation between the gene expression levels of DCC, MMP9 and ETS1 and MIRS score in the GSE86166, respectively. **I**. Sankey diagram for the MIRS values with different intrinsic molecular subtypes in TCGA patients. **J**. Violin plots for the distribution of MIRS values in different intrinsic molecular subtypes at TCGA BRCA cohort. The p-values were calculated using Kruskal-Wallis test. *p<0.05; **p<0.01; ***p<0.0001.

Moreover, 730 genes were identified to be correlated to the 12 genes of MIRS (Spearman Correlation Coefficient ≥ 0.4) using GSE86166, subsequently, functional enrichment analysis achieved via METASCAPE, indicating various immune-related processes and pathways were significantly enriched, including T cell activation, Cytokine-cytokine receptor interaction and B cell activation (Figure 4B). This observation discloses a strong correlation of MIRS with immune activity. Alternatively, we applied ssGSEA analysis to evaluate the immune infiltration level in GSE86166 using 17 immune-related biological functions and pathways derived from the immune-related database ‘ImmPort’ (22). The result illustrates that most of the 17 items show significant difference between MIRS^high^ and MIRS^low^ group (Figure 4C). Notably, all immune-related biological processes and pathways exhibit significantly higher level of immune infiltration in MIRS^low^ group (Figure 4C), which is consistent with our previous analysis. Moreover, we estimated the correlation of MIRS with three important immune checkpoint molecules: PD-1, PD-L1 and CTLA4. As illustrated in Figure 4E, compared with MIRS^high^ group, MIRS^low^ group shows significantly higher expression (Wilcoxon test P < 0.0001). MIRS scores are moderately correlated to the expression levels of PD-1, PD-L1 and CTLA4 (Figure 4D). Overall, the differences in tumor immunogenicity between the MIRS groups are significant, MIRS^high^ group has relatively low immune infiltration level while MIRS^low^ group has relatively high immune infiltration level. Similar results were also observed in TCGA and GSE96058 cohort (Supplementary Figure S8). This finding further suggested MIRS^low^ group maight have better response in therapy of immune checkpoint blockade.

To investigate the correlation between MIRS score and metastatic mechanism, we firstly downloaded the metastasis breast cancer (METABRIC) cohort from human cancer metastasis database https://hcmdb.i-sanger.com/, which contains primary tumor and metastatic tumor. Then the functional analysis achieved by GSEA detects 23 qualified metastasis-related gene sets (NES| > 1, NOM p-value < 0.05 and FDR q-value < 0.25). After that, ssGSEA analysis was used to evaluate the above significant metastatic pathways. We observe that the metastatic pathways exhibit significant difference between two MIRS groups, and the majority of MIRS^high^ group had higher ssGSEA score (Figure 4G). A higher ssGSEA score suggests high activity of metastatic processes. Similar results are found in TCGA and GSE96058 cohort (Figure S9-S10). Furthermore, the expression discrepancy of three well-known genes (DCC, MMP9 and ETS) were found to be correlated to the invasion and metastasis in breast cancer (29) (Figure 4F), and MIRS exhibits moderately negative correlation with the expression of these genes (Figure 4H).

We also examined the relationship between intrinsic molecular subtypes and MIRS. In breast cancer, major subtypes based on the ER, PR and HER2 exist on tumor cells. As shown in Figure S15, although the expression levels of ER, PR and HER2 were moderately correlated with MIRS, the differences in the expression levels of ER, PR and HER2 between high and low MIRS subtypes were statistically significant in TCGA and GSE86166. Additionally, for TCGA cohort, we noticed the imbalanced proportions of intrinsic molecular subtypes between MIRS^high^ and MIRS^low^ groups (Figure 4I). 48.09% of LumA tumor and 22.5% of Normal-like tumor are present in MIRS^high^ group whereas 32.33% of LumB tumor in MIRS^low^ subtype. However, we found that higher proportion of Basal-like tumor was present in MIRS^low^ subtype. In Muenst et al.’ study (30), they pointed out that the number of tumor-infiltrating lymphocytes was the highest in the basal-like subtype, which may support a high enrichment of basal-like tumor in MIRS^low^ group. We also found that the normal-like had significantly lower MIRS than other molecular subtypes, in contrast to the LumB subtype had the highest MIRS (Figure 4J). In addition, a statistically significant difference was detected among these five intrinsic molecular subtypes by using Kruskal-Wallis method (Figure 4J). Similar results were found in METABRIC cohort (Supplementary Figure S11). These analyses indicate that MIRS group exhibited chaotic correlation with classic molecular subtypes, which could be attributed to the high tumorous heterogeneity in breast cancer.

### Identification of MIRS related biological characteristics in prognosis of breast cancer

The above analyses implied high correlations between MIRS and tumor-infiltration microenvironment as well as tumor metastasis. We further explore the molecular mechanism of 12-gene panel underlying the prognosis of breast cancer. Initially, through literature, we found that the majority of those prognosis-related genes, except for *APOA5*, has been reported to be involved in the processes of tumorigenesis (Table S6). It is worth mentioning that APOA5, encoding an apolipoprotein, is associated with cardiovascular diseases (31, 32), but little work studies its roles in tumorigenesis and prognosis. To delineate its potential prognostic role in breast cancer, we divided APOA5 expression into four quartiles, then GSEA analysis between the highest and lowers quartiles in TCGA-BRCA was conducted. Interestingly, many metastatic and immune-related pathways were observed to be enriched in the highest quartile, including EMT, TNFα signaling and Immune response regulating signaling pathways (Figure 5A and S12A). Subsequent survival analysis of pan-cancer based on TCGA cohorts was performed via Kaplan-Meier Plotter (https://kmplot.com/analysis/) (33), indicating that APOA5 may serve as prognostic indicator in many cancers (Figure S12B). The breast cancer patients with the highest APOA5 expression have a worse survival outcome (Figure S12B). Overall, our analysis hinted that APOA5 may exert its prognostic function to affect the immune activity in breast cancer, and it is likely to be a potential target for the future research of breast cancer therapy.

**Figure 5.**
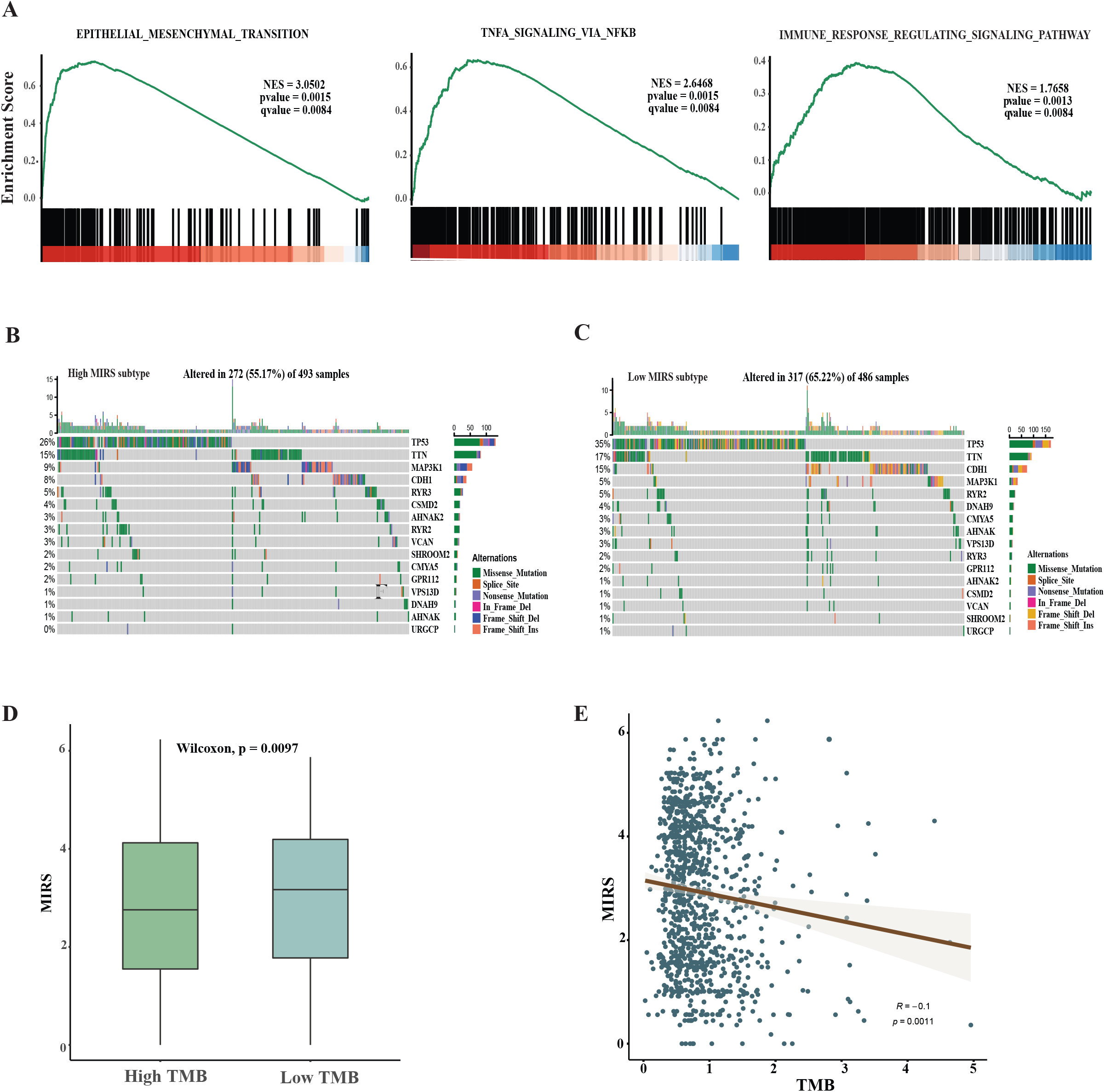
Identification of MIRS-related biological characteristics in prognosis of breast cancer. **A**. GSEA enrichment plots in TCGA. **B**. The Oncoplot of top 10 genes with the highest mutation frequency in high MIRS group (TCGA data). **C**. The Oncoplot of top 10 genes with the highest mutation frequency in low MIRS group (TCGA data). **D**. Boxplots of the MIRS score between the high and low TMB subtypes in TCGA data. The p-values were calculated using Wilcoxon rank sum test. **E**. The spearman correlation between the MIRS score and TMB values in TCGA data.

Genomic mutations are mostly involved in the survival prognosis of various cancers (34). Thus, we tested the associations between somatic mutations and MIRS in TCGA BRCA data. According to the analysis in the study of Chen et al (35), only the genes with somatic mutation frequencies more than 2.5% were included. By analyzing the mutation annotation of TCGA BRCA cohort, we selected the top 10 genes by mutation frequency. As provided in Figure 5B and C, MIRS^low^ group has increased frequency of mutation events than MIRS^high^ group. Rizvi et al (36) and Capalbo et al’ studies (37) demonstrated that the patients with more mutations might have an increased number of neoantigens that enhance response to immunotherapy. This result might explain, in the present study, the reason that MIRS^low^ group has better prognostic outcomes than MIRS^high^ group.

Recently, tumor mutation burden (TMB) is the paramount prognostic measure in cancer survival (38). We further investigated the associations between MIRS and TMB. As illustrated in Figure 5D, the patients in MIRS^low^ group exhibited markedly increased TMB when compared with those with MIRS^high^ group. Lee et al (39) and Karn et al’s studies (40) showed that high TMB was associated with improved survival. Additionally, Chen et al (35) reported that the increased TMB was correlated to improved response to PD-1 blockades therapy. Correlation analysis between MIRS and TMB demonstrated that MIRS score was negligibly correlated with TMB (Spearman coefficient: R = -0.1, p = 0.0011; Figure 5E). These findings indicate that MIRS may be related to immunotherapy response, and the patients with lower MIRS may have probably response in immunotherapy.

### The role of MIRS in the prediction of therapeutic benefits

To explore predictive ability of MIRS in immunotherapy for each patient, T cell inflamed score (TIS), IFN -gamma signature, antigen presenting machinery genes (APM) and Immunotherapyscore (IPS) (20, 41, 42), which are prevailing predictors of clinical response to immunotherapy across different tumor types were compared. Notably, the higher of TIS, IFN-gamma score, APM and IPS mean that patients receiving immunotherapy are more likely to response All patients in GSE20711 and GSE58812 with MIRS^low^ showed significantly increased predictor scores than those with MIRS^high^ (Figure 6A and S13A), which hints that MIRS^low^ group is more likely to have immunotherapy response. To further appraise the prognostic capability of MIRS^low^ group in immunotherapy, the differences in overall survival between MIRS^high^ and MIRS^low^ groups were compared using KM survival analysis in breast cancer testing cohort. Unfortunately, there are hitherto few public datasets of breast cancer patients receiving immunotherapy. Instead, the data of melanoma from Liu et al (43) and TCGA-SKCM dataset with patient receiving immunotherapy were used in present analysis. As a result, compared with PD-1 and TMB biomarkers upon receiving anti-PD-1 treatment, MIRS showed robust AUCs (Figure 6B-D). Furthermore, the patients with MIRS^high^ have significantly shorter overall survival than their counterparts (Figure 6E and Figure S13B). MIRS significantly increases in patients with stable disease (SD) or progressive disease (PD) when compared with those with complete response (CR) or partial response (PR) (Figure 6F and Figure S13CB). Besides, the distributions of CR/PR and SD/PD across MIRS^high^ and MIRS^low^ groups were also validated. We found that patients in MIRS^low^ group had better response to immunotherapy than those in MIRS^high^ group (Figure 6G and Figure S13DC).

**Figure 6.**
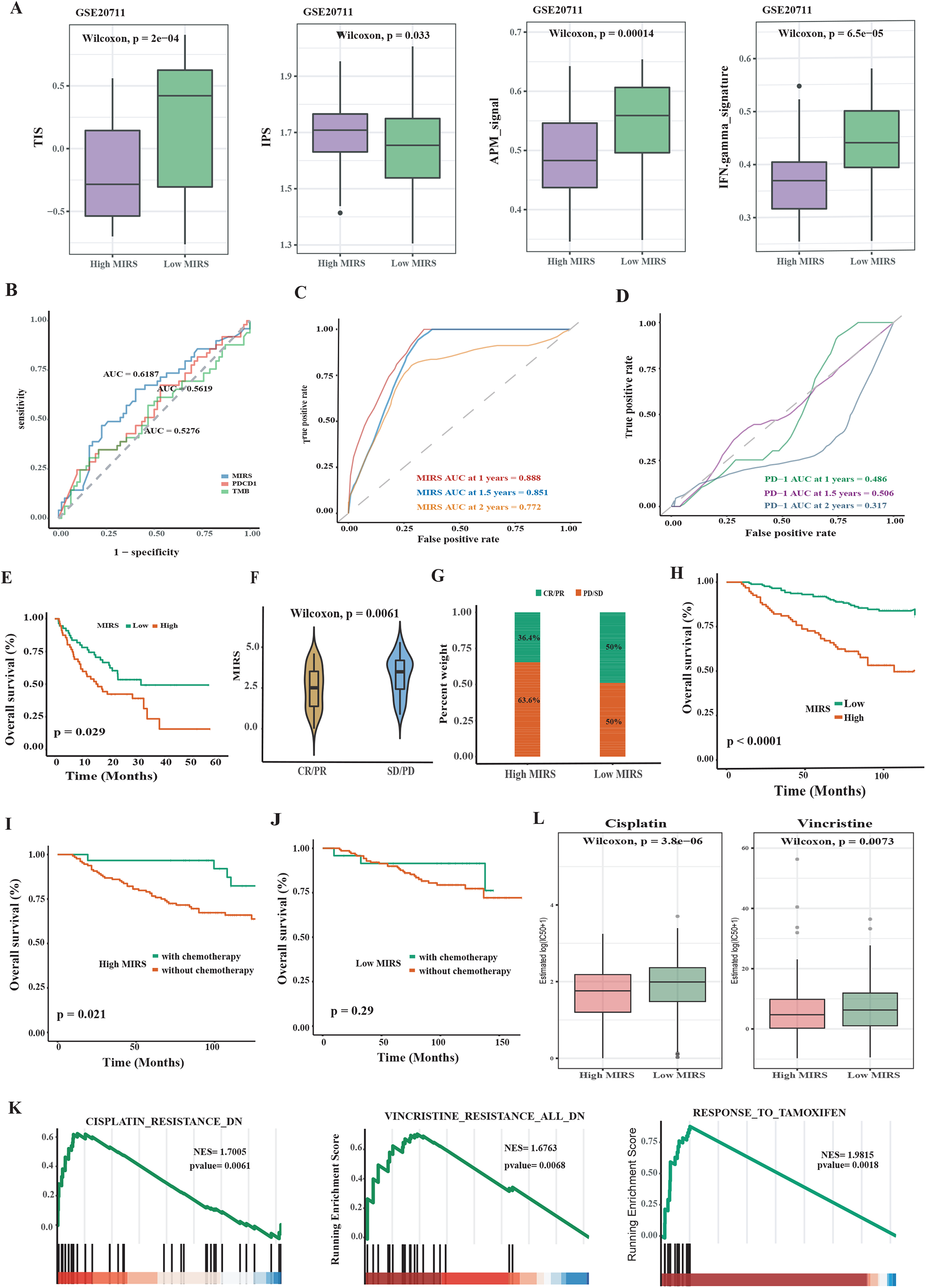
The therapeutic benefit of the MIRS value. **A**. The boxplot of TIS, IPS, APM score and IFN gamma score between the high and low MIRS in GSE20711. **B**. ROC curves between the expression level of PD-1, TMB and MIRS of anti-PD1 immunotherapy response prediction in Liu et al data. **C**. Time-dependent ROC curves of MIRS for anti-PD1 immunotherapy response prediction in the Liu et al data. **D**. Time-dependent ROC curves of the expression level for anti-PD1 immunotherapy response prediction in Liu et al data. **E**. Kaplan-Meier curves of overall survival according to MIRS subtypes in the Liu et al data. **F**. Violin plot illustrating the distribution of MIRS for patients with different immunotherapy response in Liu et al data. **G**. Bar graph showing the number of clinical responses to anti-PD-1 immunotherapy in the high and low MIRS subtypes in Liu et al data. **H**. Kaplan-Meier curves of overall survival according to MIRS subtypes with chemotherapy in GSE20685. **I**. Kaplan-Meier curves of overall survival according to the high MIRS subtype with or without chemotherapy in GSE20685. **J**. Kaplan-Meier curves of overall survival according to the low MIRS subtype with or without chemotherapy in GSE20685. **K**. GSEA predict that high MIRS group is negatively correlated with drug resistance in TCGA cohort. **L**. Chemotherapeutic sensitivity of two drugs (Cisplatin, Vincristine) were estimated and compared in TCGA cohort.

Moreover, to assess therapeutic value of MIRS in chemotherapy, we examined its predictive potential in GSE20685 with the breast cancer patients who receive adjuvant chemotherapy. The optimal cutoffs of MIRS were determined by the median cutoff, then the patients were stratified into MIRS^high^ and MIRS^low^ group. Survival analysis displays that the breast cancer patients with MIRS^low^ had much better survival than those with MIRS^high^ in adjuvant chemotherapy cases (Figure 6H). We also investigated the prognosis of different MIRS subtypes with or without adjuvant chemotherapy. As illustrated in Figure 6I, we found that MIRS^high^ group had statistically significant differences between the patients who were treated with adjuvant chemotherapy and those without adjuvant chemotherapy. However, a consistent result was not observed in those patients with MIRS^low^ (Figure 6J). These results indicated that adjuvant chemotherapy might be more beneficial to MIRS^high^ group. Based on the gene sets of different drug treatments retrieved from MSigDB database, GSEA predicted that MIRS^high^ was significantly correlated with drug sensitivity in TCGA cohort (Figure 6K). Moreover, the R package pRRophetic was used to estimate the sensitivity of three chemotherapeutic drugs, including cisplatin, tamoxifen and vincristine, which have been commonly used in breast cancer treatment. The results showed that estimated IC50 values of cisplatin and vincristine significantly decrease in MIRS^high^ subtype (Figure 6L). We did not display IC50 boxplot of tamoxifen due to the R package ‘pRRophetic’ does not contain resistant information regarding tamoxifen.

These results suggest that MIRS holds massive potential for predicting the response to chemotherapy and immunotherapy in breast cancer patients. In brief, the patients with MIRS^high^ may benefit from the chemotherapy, and patients with MIRS^low^ are likely to be more sensitive to the immunotherapy.

### Comparison of MIRS with the previously prognostic models

Before the creation of MIRS, Shimizu et al (26) demonstrated that 23-gene panel (mPS) helps predict OS in breast cancer patients based on analogous neuron network model; Cui’s score (44) constructed 8-gene signature based on traditional Lasso Cox model. We then comprehensively evaluate the prognostic power of our MIRS, mPS and Cui’s score by 0prognostic Cox analyses based on a variety of public datasets. Our MIRS performed very well in different cohorts (Figure 7A). Although mPS showed to be more robust than MIRS in many datasets, some of the HRs in mPS panel were not significant (P value > 0.05) (Figure 7B). Cui’s score performed the worst among these models (Figure 7C).

**Figure 7.**
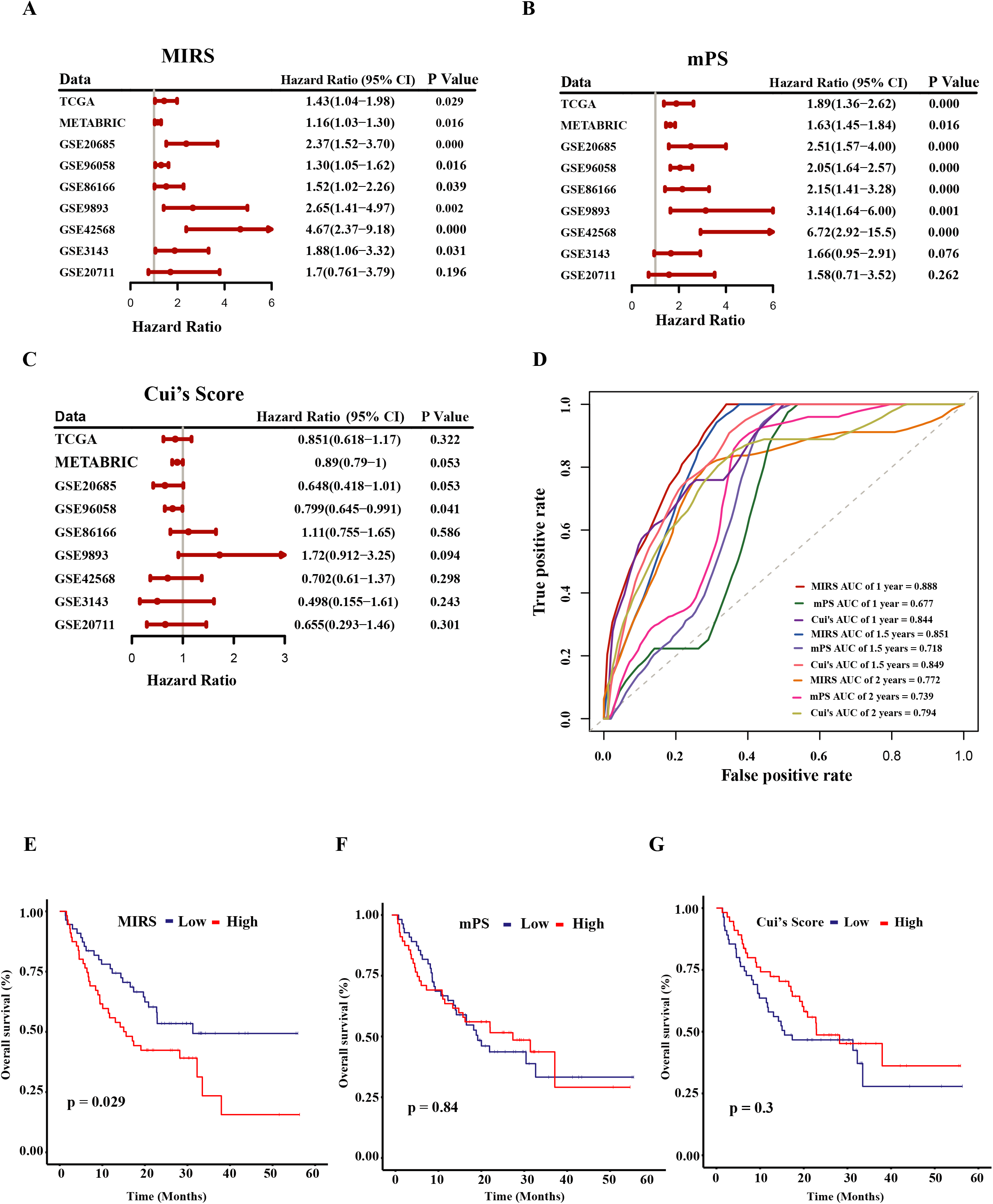
Compare MIRS with previous prognosis signatures. **A**. A meta-analysis was performed using the prognosis results of MIRS in nine public datasets. **B**. A meta-analysis was performed using the prognosis results of mPS in nine public datasets. **C**. A meta-analysis was performed using the prognosis results of Cui’s score in nine public datasets. **D**. Time-dependent ROC curves of anti-PD-1 immunotherapy on the 1-,1.5-,2-year survival rates for Liu et al data. **E**. Kaplan-Meier curves of overall survival according to MIRS subtype with immunotherapy in Liu et al data. **F**. Kaplan-Meier curves of overall survival according to mPS subtype with immunotherapy in Liu et al data. **G**. Kaplan-Meier curves of overall survival according to Cui’s score subtype with immunotherapy in Liu et al data.

Furthermore, we scrutinized the predictive potential of these three models in the response to immunotherapy. The malignant melanoma cohort data (43) that receives anti-PD-1 therapy was used. The optimal cutoffs of Cui’s score and mPS value were determined by the median. KM survival curves of MIRS show a significant difference in OS between MIRS^high^ and MIRS^low^ group (Figure 7E). On the contrary, the survival analysis of mPS and Cui’s score revealed that patients with low mPS or Cui’s score showed no statistically significant difference when compared with those with high mPS or Cui’s score (Figure 7F-G). MIRS, mPS and Cui’s score were also examined with time-dependent ROC analysis in the testing cohort for prediction in immunotherapeutic benefits. Notably, our MIRS exhibited much better predictive ability than mPS and Cui’s score for OS at 1 year, 1.5 years, and 2 years, respectively (Figure 7D).

## Discussion

With the development of transformative technologies, analyses of high throughput sequencing data have significantly deepened the understanding of modern biology, enabling the scientists to thoroughly explore key characteristics in a variety of cancers. Metastasis and tumor-immune infiltration are two of the major characteristics, and have been extensively proven to be associated with tumorigenesis, drug resistance and prognosis in breast cancer (43). Quite a few studies have disclosed the roles of metastasis and tumor-immune infiltration as prognostic factors in predicting the survival outcomes for breast cancer (45). Unfortunately, breast tumors are highly heterogeneous among individuals, and much current work has only considered organ-specific metastasis or immune infiltration level and thus insufficient to achieve robust predictive power on prognosis. To address this issue, in this study we developed a comprehensive and efficient prognosis model, considering metastasis and immune infiltration levels together, to aid clinicians in providing precise treatment strategies.

Given the promising predictive value of MIRS, we systematically investigated the relationships between MIRS and clinical pathological characteristics. In different sequencing platform data, MIRS demonstrated as an independent prognosis factor compared with other conventional clinical features (Figure S14). As illustrated in Figure S17A, we observed differences between MIRS and Age, Gender and Metastasis variables. Subsequently, we used decision curve analysis (DCA) to decipher the effect in combining MIRS with clinical indicators. In the DCA analysis, the net benefit of clinical indicators combined with MIRS were better than of sole clinical indicator (Figure S17B). Additionally, we employed TCGA and GSE96058 datasets to investigate whether MIRS is suitable for all BRCA subtypes due to its complete subtype information. However, we have not observed consistently predictive ability in both datasets (Figure S18). This unsatisfied performance may come from the fact that we built our MIRS model without tumor subtype information. Together, these results demonstrate the validity and reliability of MIRS in clinical applications, but it no suitable to all subtypes in breast cancer.

Next, we compared MIRS with the representative prognostic models, mPS and Cui’s score. Univariate cox regression analysis using nine public cohorts indicated that MIRS and mPS performd well in most cohorts. These results indicated that, constructing a prognostic system considering only metastatic features may be insufficient. Compared with AI methods, traditional survival model showed weak power. Nonetheless, mPS scoring system, based on an analogous AI approach, does not work well in predicting immunotherapeutic. It might explain that the establishment of mPS does not consider immunogenomic features, thus failing to achieve satisfactory immunotherapeutic prediction.

Apart from being informative regarding prognosis, MIRS can also act as an independent predictor to guide therapeutic strategies. Our analyses indicated that MIRS^high^ group had lower TIS, IPS, IFN-gamma score and APM score, implying MIRS^high^ group is more likely to escape from immunity in breast cancer. For further validation, we tested if the OS between MIRS^high^ and MIRS^low^ groups was associated with immunotherapy. We used two malignant melanoma cohorts with immunotherapeutic information by conducting KM analysis. This survival analysis showed that MIRS^low^ group exhibited improved survival and better response to immunotherapy than MIRS^high^. We speculate that the immunotherapy may achieve beneficial treatment for MIRS^low^ patients.

Currently, chemotherapy is one of the main treatments for breast cancer. Hence, it is necessary to identify patients who may potentially benefit from chemotherapy. Through the analysis of breast cancer patients with chemotherapy clinical information, we found that patients with MIRS^high^ respond better to chemotherapy than patients with MIRS^low^. Chemotherapy has been reported to be related to immune infiltration (46). In the Ahn et al’s study (47), they demonstrated that the high level of the CD8+ TILs filtration is associated with chemotherapy resistance. This may be the reason that high filtration MIRS^low^ subtype shows favorable chemotherapy. These results emphasize the significance of MIRS^high^ patients who could benefit from chemotherapy.

As a gene prognostic signature particularly designed for breast cancer patients, MIRS is a novel and robust approach in risk stratification and personalized treatment. However, there are still flaws in the current study. First, due to the remarkable intra-tumor heterogeneity in breast cancer, we cannot cover all metastatic signatures despite a large numbers of breast cancer patients used in this study. Second, only the median cutoff of MIRS is used to classify the patients into high and low subtypes, the optimal cutoff of MIRS would be needed to provide rational strategies. Lastly, all the conclusions in this research are obtained from *in silico* studies, clinical experiments are required to confirm our findings.

MIRS has the potential to assist oncologists to screen patients who are more likely to benefit from immunotherapy or chemotherapy. It would be of great significance to validate the value of MIRS in prospective clinical trials.

## Contributors

CH and XDZ conceived the presented idea. CH, DLL and MD collected the public data, MD and CH developed the methodology, MD and DLL analyzed the data under the supervision of CH. CH and MD took the lead in drafting the manuscript with input from all authors. CH, ELHL and XDZ revised the manuscript, PYZ and BQS interpreted results from a clinical point of view. All authors read and approved the final manuscript.

## Data Availability

All data produced in the present work are contained in the manuscript

## Acknowledgements

This work was supported by Dr. Neher’s Biophysics Laboratory for Innovative Drug Discovery (File no. 001/2020/ALC), by the Science and Technology Development Fund, Macau Government (File no. 0020/2021/A), by the University of Macau (grant numbers: FHS-CRDA-029-002-2017 and MYRG2018-00071-FHS), Zhongnanshan Medical Foundation of Guangdong Province (grant number: ZNSA-2021016) and the Science and Technology Development Fund, Macau SAR (File no. 0004/2019/AFJ and 0011/2019/AKP).

## Data sharing statement

Data are available in a public, open access repository. All used data in the current study are downloaded from Gene Expression Omnibus (GEO, https://www.ncbi.nlm.nih.gov/geo/) and The Cancer Genome Atlas (TCGA) database (https://cancergenome.nih.gov/).

