## Supplemental methods, tables and figures for "MIRS: an AI scoring system for predicting the prognosis and therapy of breast cancer"

Supplementary Information

A Metastatic and Immunogenomic Risk Score precisely predicts prognosis and therapy response of patients with Breast cancer

**Chen Huang^1,3^**^†^**, Min Deng^2^**^†^**, Dongliang Leng^2†^, Elaine Lai-Han Leung^1,3^, Baoqing Sun^4^, Peiyan Zheng^4^, Xiaohua Douglas Zhang^2*^**

^1^ Dr. Neher's Biophysics Laboratory for Innovative Drug Discovery, Macau University of Science and Technology, Macau, SAR, China

^2^ CRDA, Faculty of Health Sciences, University of Macau, Taipa, Macau

^3^ Stat Key laboratory of Quality Research in Chinese Medicine, Macau Institute For Applied Research in Medicine and Health, Macau University of Science and Technology, Macau, SAR, China

^4^ Department of Allergy and Clinical Immunology, State Key Laboratory of Respiratory Disease, National Clinical Research Center of Respiratory Disease, Guangzhou Institute of Respiratory Health, First Affiliated Hospital of Guangzhou Medical University, Guangzhou, Guangdong, China

^†^These authors contributed equally to this work

### Supplementary methods

#### Data information

Two public datasets comprising 1,243 breast cancer patients were analyzed in the training phase to generate risk score. The cohorts GSE86166, GSE96058, GSE20685, GSE20711, GSE58812, GSE9893, GSE3143, GSE425678 and METABRIC which in total comprised 6598 breast cancer patients were used to test the robustness of the established risk score in the validation phase. Expression profiles and clinical data from the skin cutaneous melanoma dataset (TCGA-SCKM) that received various immunotherapies, such as immune checkpoint inhibitors, vaccines, and cytokines, were downloaded from TCGA database. Liu et al data [1] that received anti-PD-1 therapy was also downloaded. These two malignant melanoma cohorts were used to gauge the predictive power of MIRS in the response to immunotherapy. All these 14 analyzed datasets were considered as complete cohort.

#### Grouping validation

For validating the rationality of the grouping of immune cell infiltration , Stromal Score, Immune Score, ESTIMATE Score, and Tumor Purity for each breast cancer patient were calculated by ESTIMATE package in R with default parameters [2]. CIBERSORT algorithm [3] was applied to assess the fractions of immune cell types from different immune infiltration groups. The immune-related family genes CD1 [4] and IL1 [5] were conducted to check the differences in expression level between these two immune infiltration groups.

**Functional enrichment analysis and** **mutation landscape analysis**

GO, KEGG and REACTOME pathway enrichment analyses were performed and visualized using METASCAPE (https://metascape.org/gp/index.html#/main/step1), and the p-value in enrichment analysis was adjusted by BH method [6]. Moreover, to predict the ability of MIRS response to immunotherapy, each breast cancer patient was scored with GSVA method using T-cell inflammatory signature (TIS) [7].

Somatic mutation information was extracted from TCGA database and visualized using the maftools. Tumor Mutation Burden (TMB) of each patient was estimated by R package TCGAmutations [8]. All parameters were set to default setting.

### Supplementary Figures and Tables

**Figures**

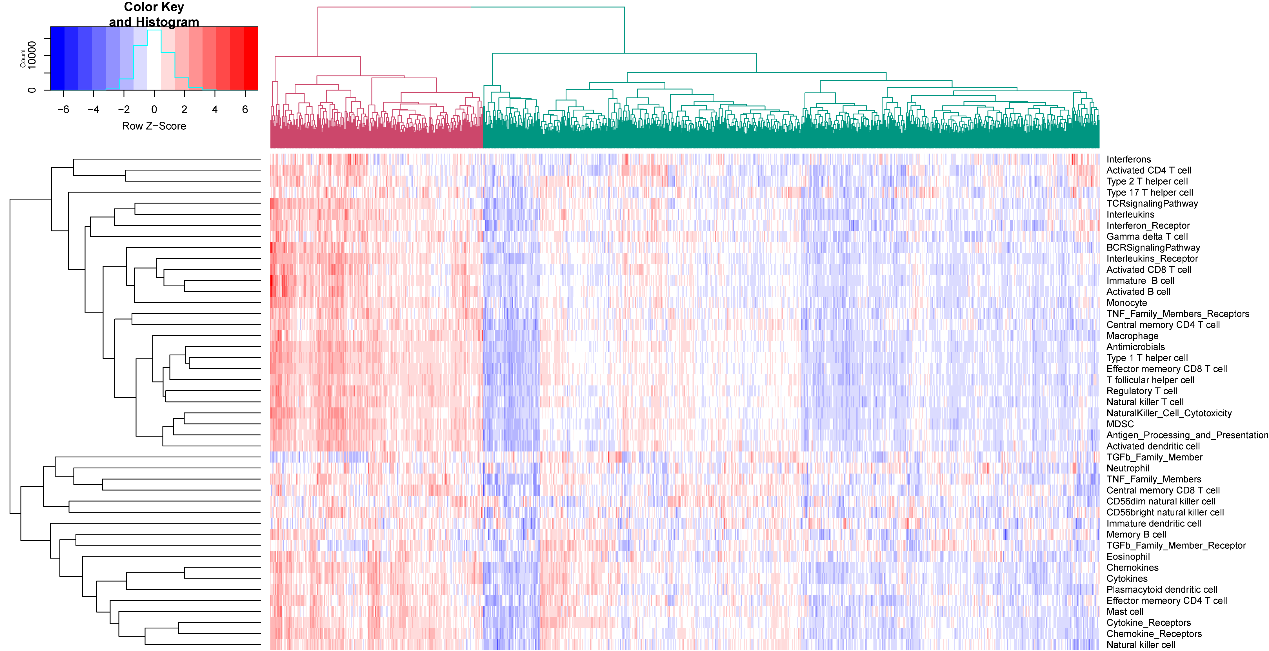

**Figure S1. Unsupervised clustering result.**

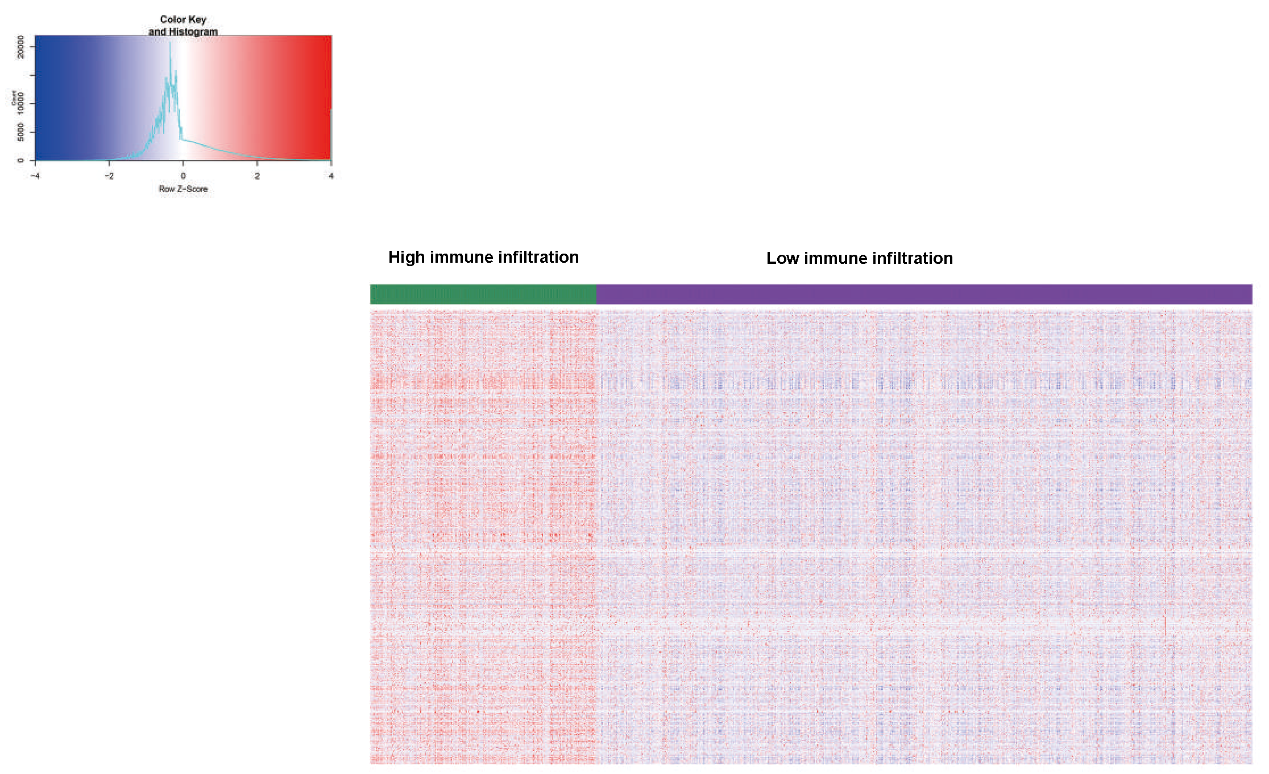

Figure S2. Heatmap of differentially expressed genes between the high and low immune infiltration groups in TCGA data.

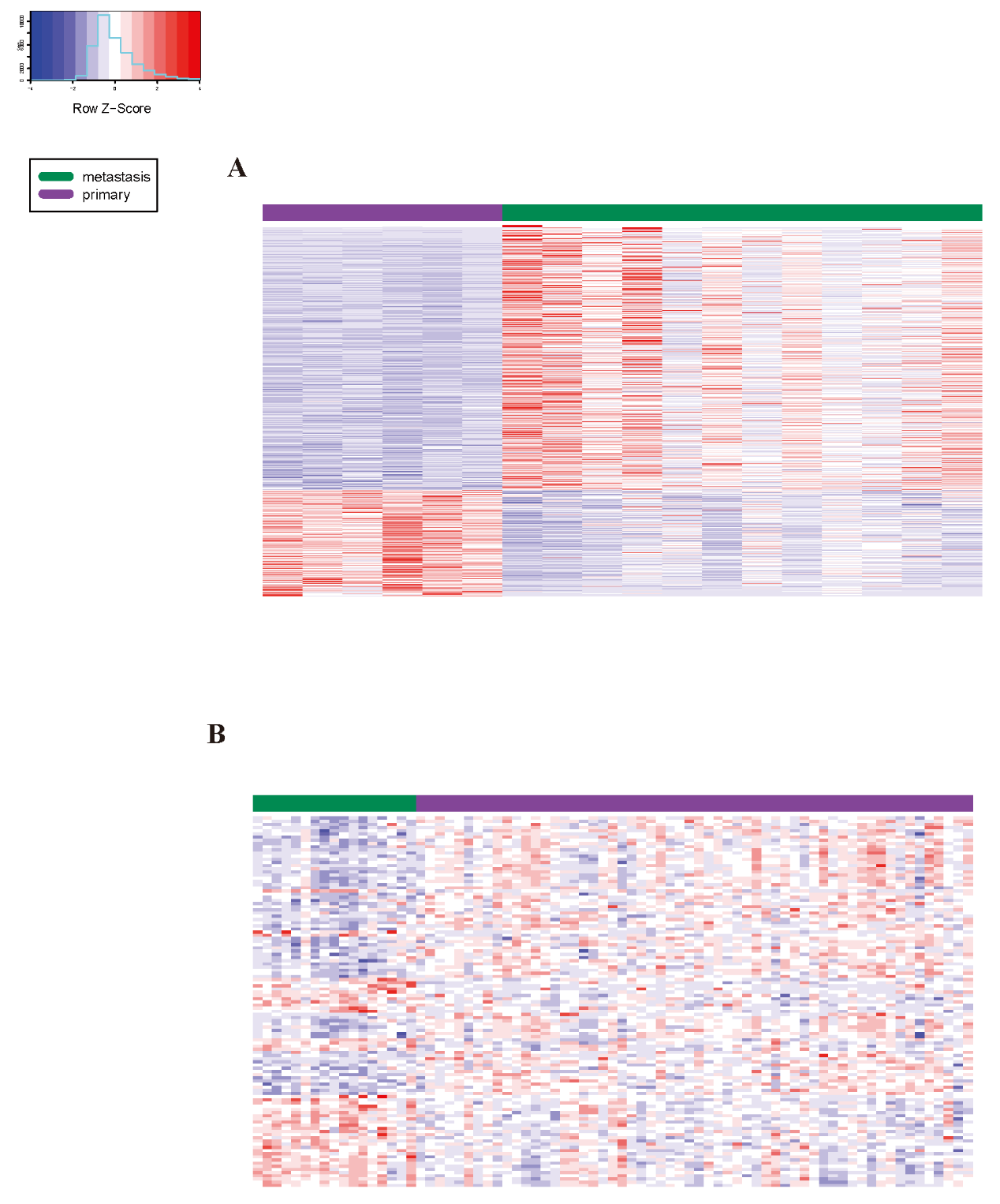

Figure S3. (A) Heatmap of differentially expressed genes between the primary and metastasis groups in GSE10893. (B) Heatmap of differentially expressed genes between the primary and metastasis groups in GSE3521.

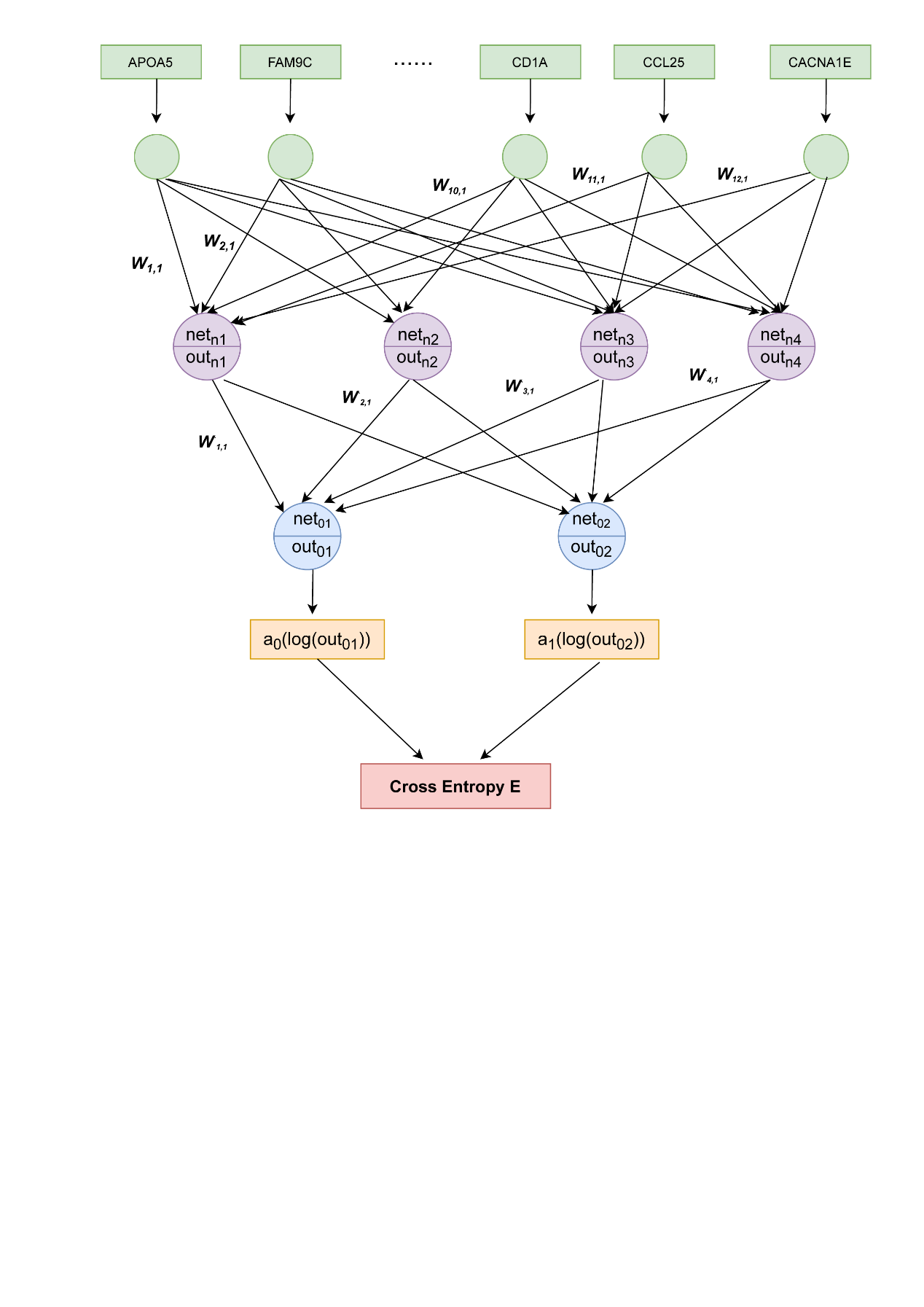

Figure S4. The workflow of Neuron network training model.

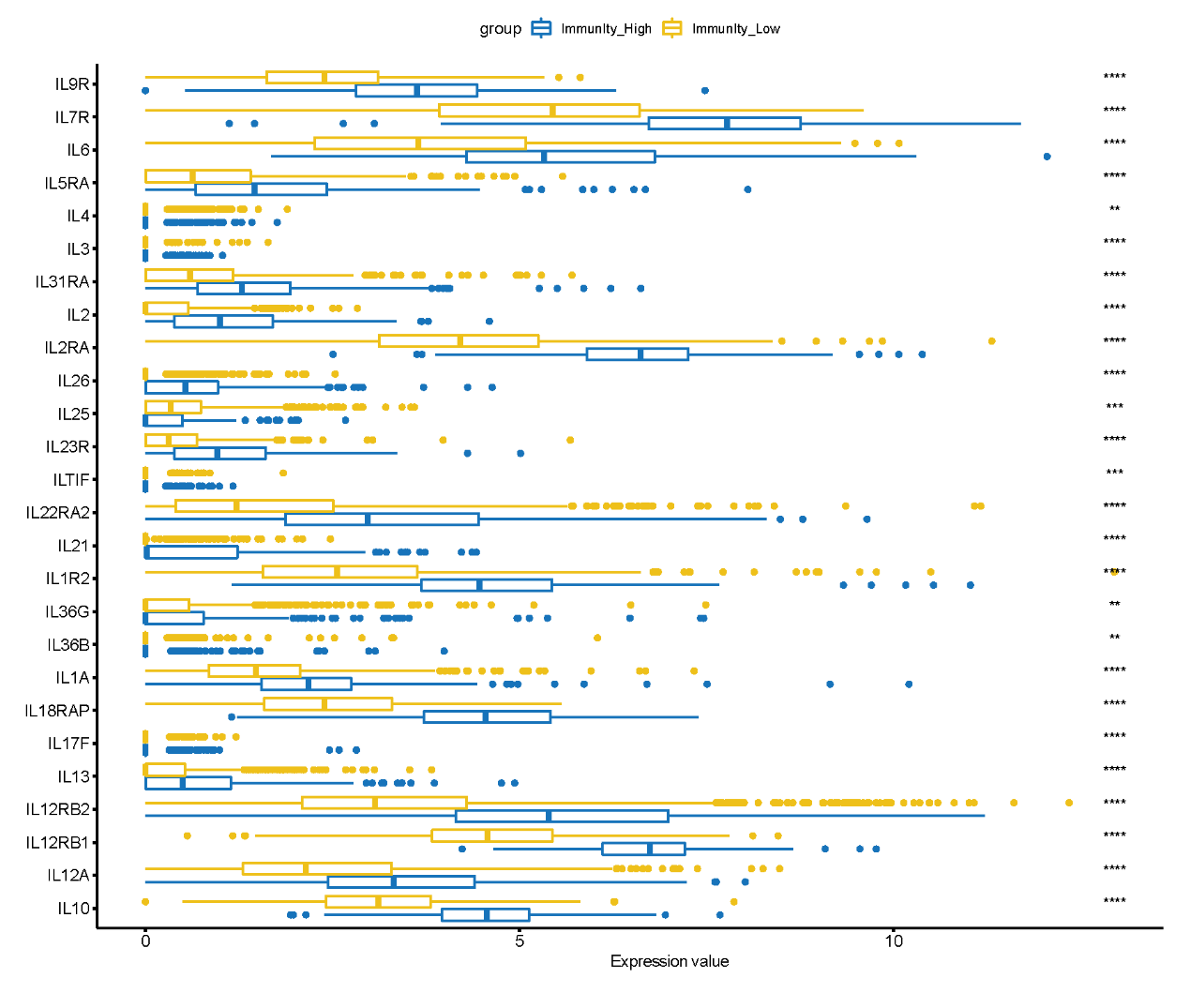

Figure S5. The expression levels of IL family gene between the high and low immune infiltration groups in TCGA data.

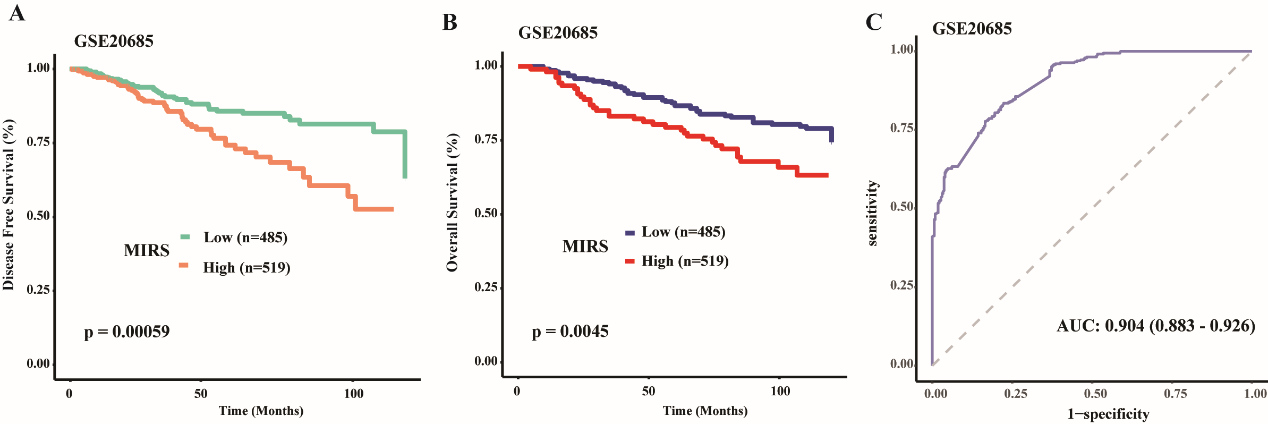

Figure S6. (A) Kaplan-Meier curves of disease-free survival according to MIRS subtypes in the TCGA data. (B) ROC curves for the BRCA patient's overall survival prediction in the GSE20685. (C) Kaplan-Meier curves of overall survival according to the MIRS subtypes in GSE20685.

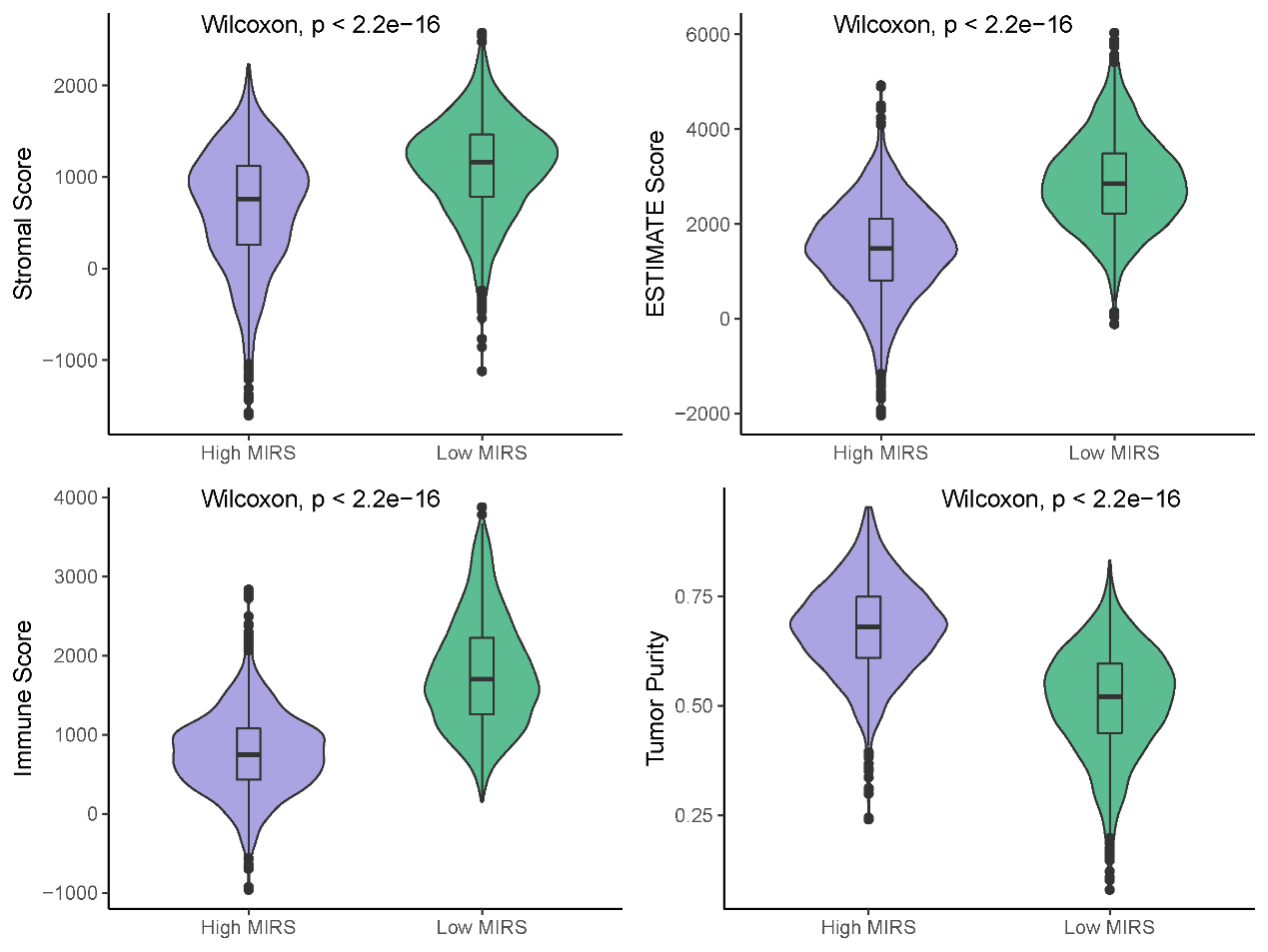

Figure S7. Comparison of the Stromal score, ESTIMATE score, Immune score, and Tumor purity between the high and low MIRS subtypes in GSE96058.

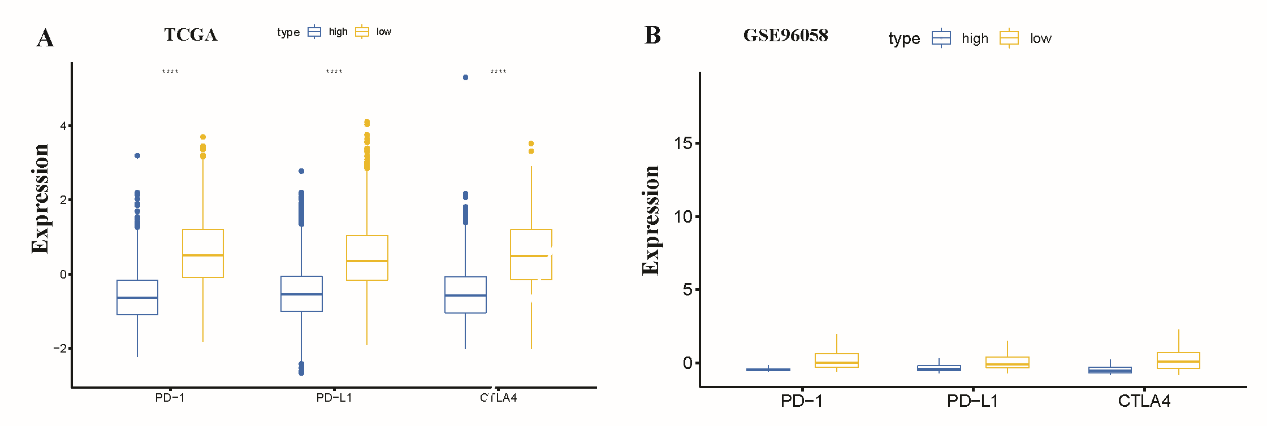

Figure S8. (A) The boxplots of PD-1, PD-L1 and CTLA4 for two MIRS subtypes in the TCGA cohort. (B) The boxplots of PD-1, PD-L1 and CTLA4 for two MIRS subtypes in GSE96058.

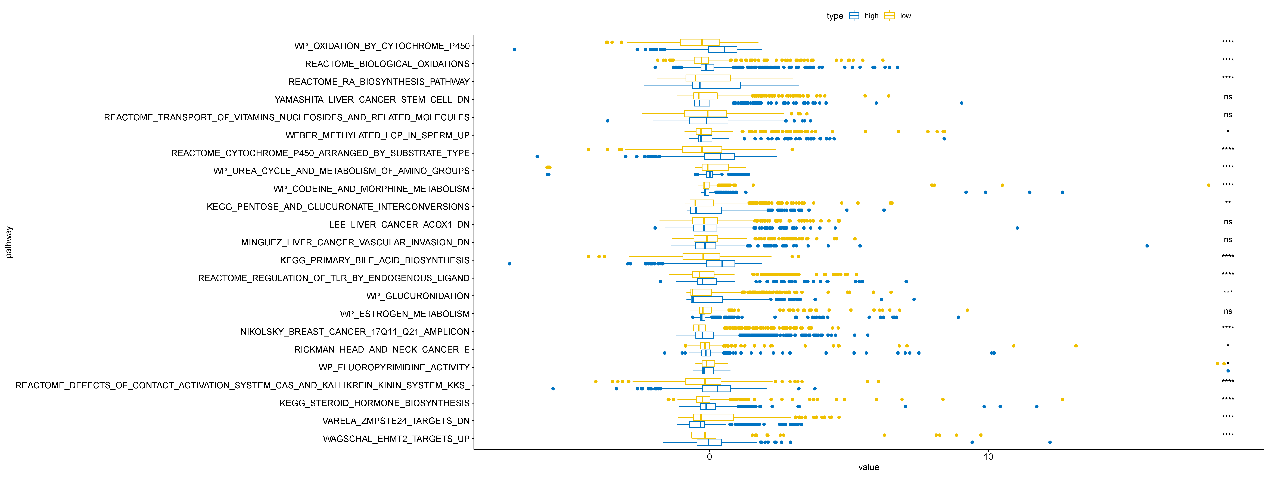

Figure S9. Boxplot of the ssGSEA score for 17 immune-related biological functions and pathways between two MIRS subtype in TCGA data.

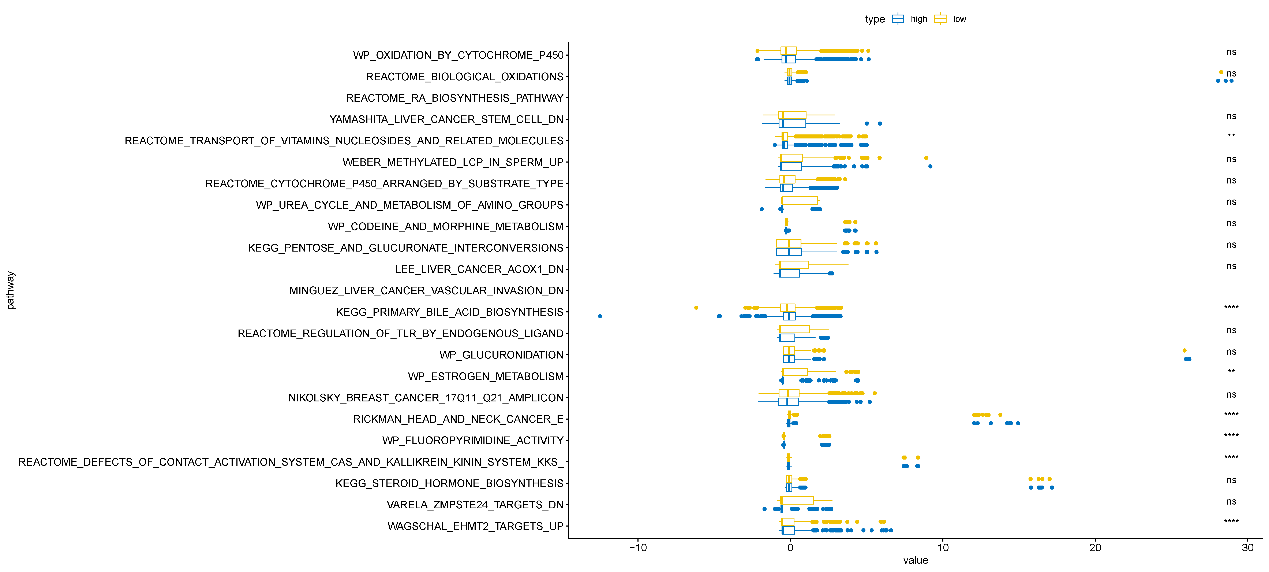

**Figure S10. Boxplots of ssGSEA score for 17 immune-related biological functions and pathways between two MIRS subtypes in the GSE96058.**

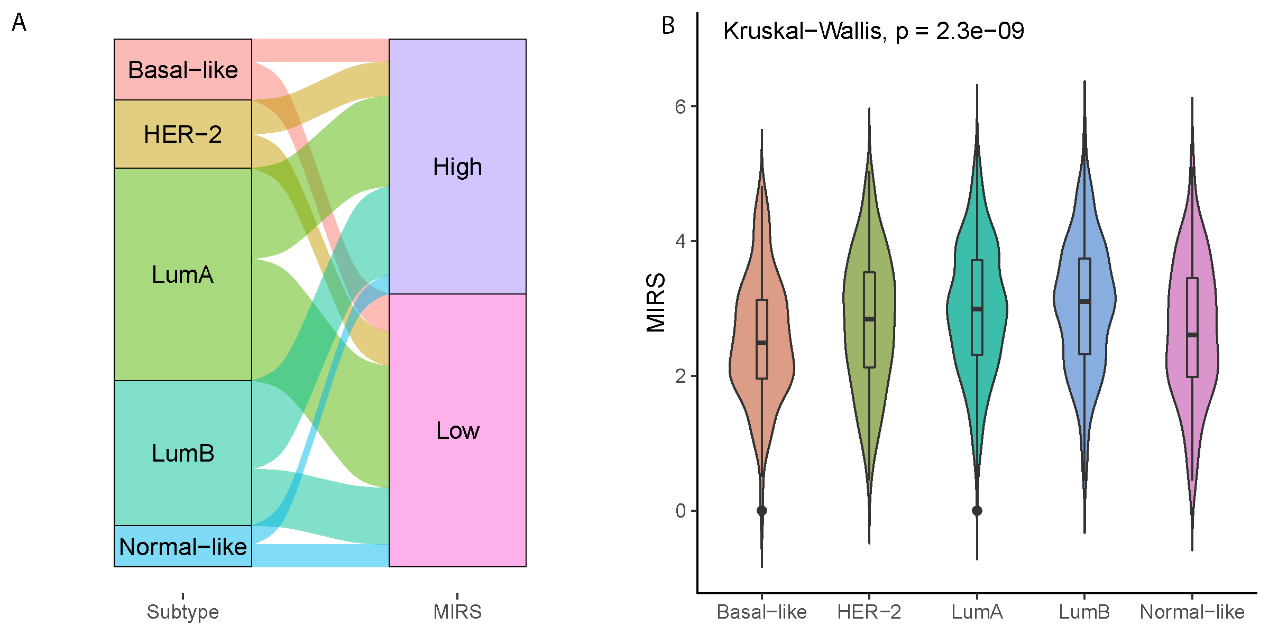

**Figure S11. (A) Sankey diagram for the MIRS values with different intrinsic molecular subset in METABRIC data. (B) Violin plots for the distribution of MIRS values in different intrinsic molecular subtypes in MTABRIC data.**

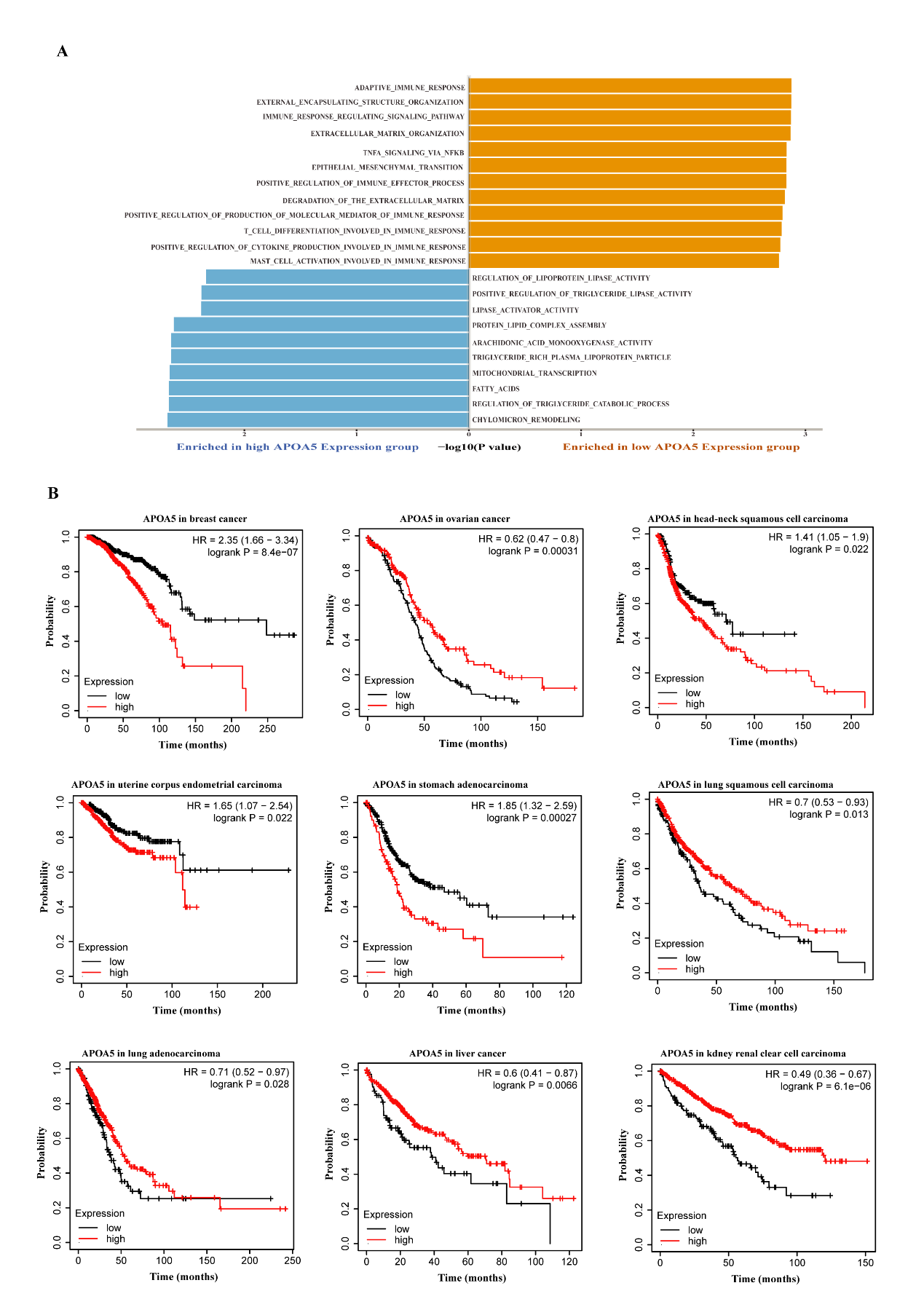

**Figure S12 Investigation the role of APOA5 (A) GSEA analysis between the highest and lowest quartile of APOA5 expression. (B) KM survival curves of APOA5 expression across different cancer types**

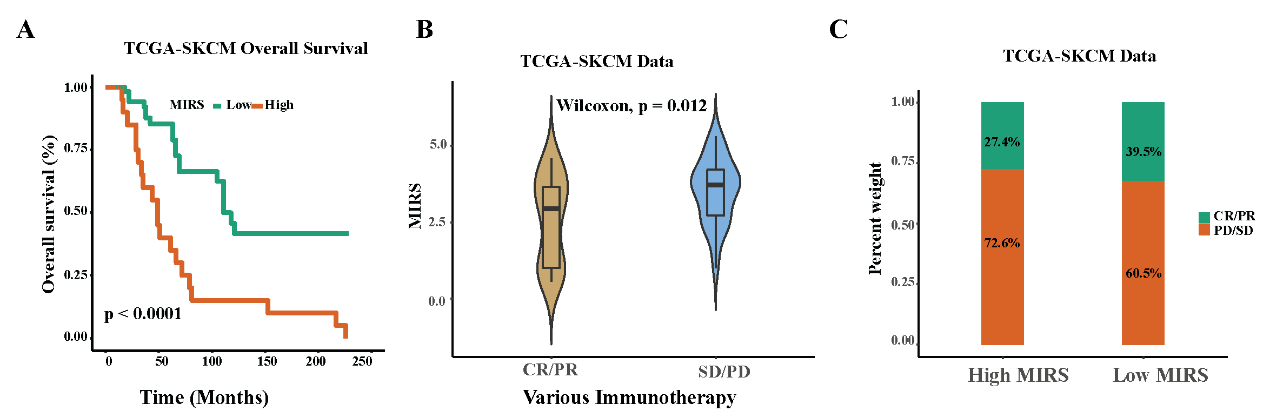

**Figure S13. (A) Kaplan-Meier curves of overall survival according to MIRS subtypes in the TCGA-SKCM data. (B) Violin plot illustrating the number of clinical responses to anti-PD-L1 immunotherapy in the high and low MIRS subtypes for the TCGA-SCKM data. (C) Bar graph illustrating the number of clinical response to anti-PD-L1 immunotherapy in the high and low MIRS subtypesfor the TCGA-SCKM data.**

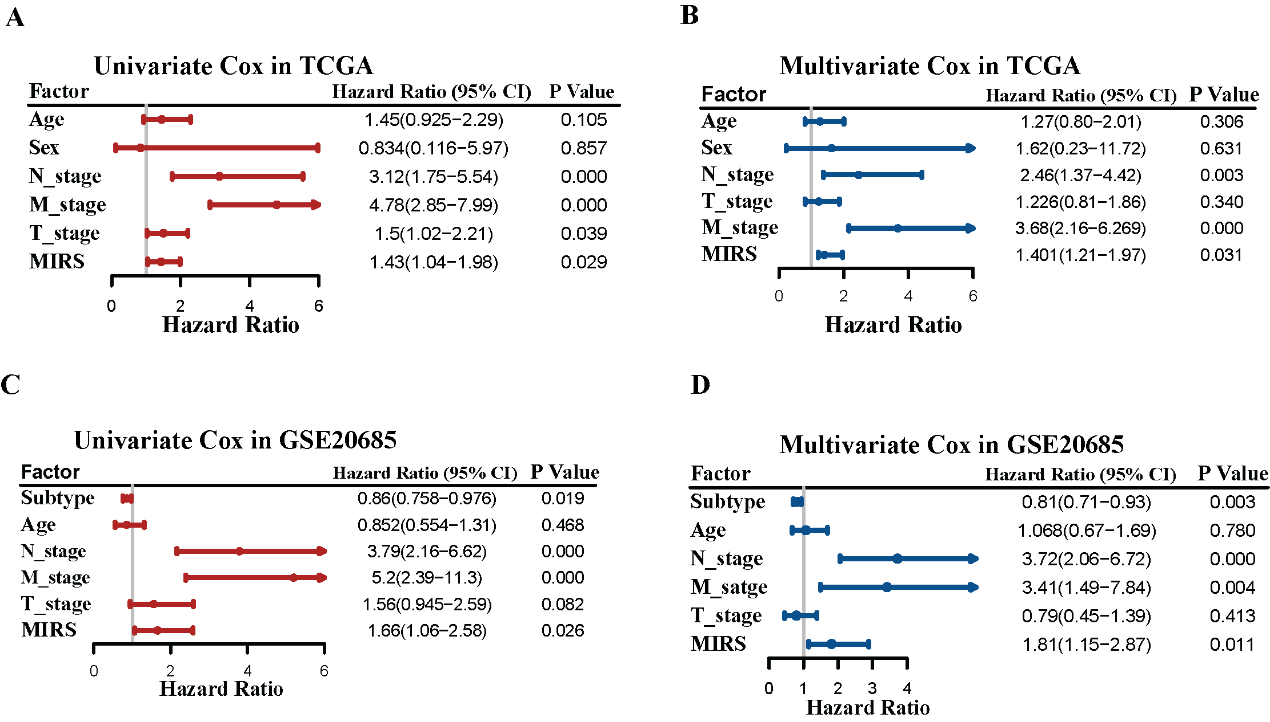

**Figure S14 Univariate and multivariate cox proportional hazard analyses in different cohorts. (A) Univariate cox analysis of overall survival in the TCGA cohort. The HR value, its 95% CI and P-value for the cox hazard model are shown. (B) Multivariate cox analysis of overall survival in the TCGA cohort. (C) Univariate cox analysis of overall survival in the GSE20685. (D) Multivariate cox analysis of overall survival in the GSE20685.**

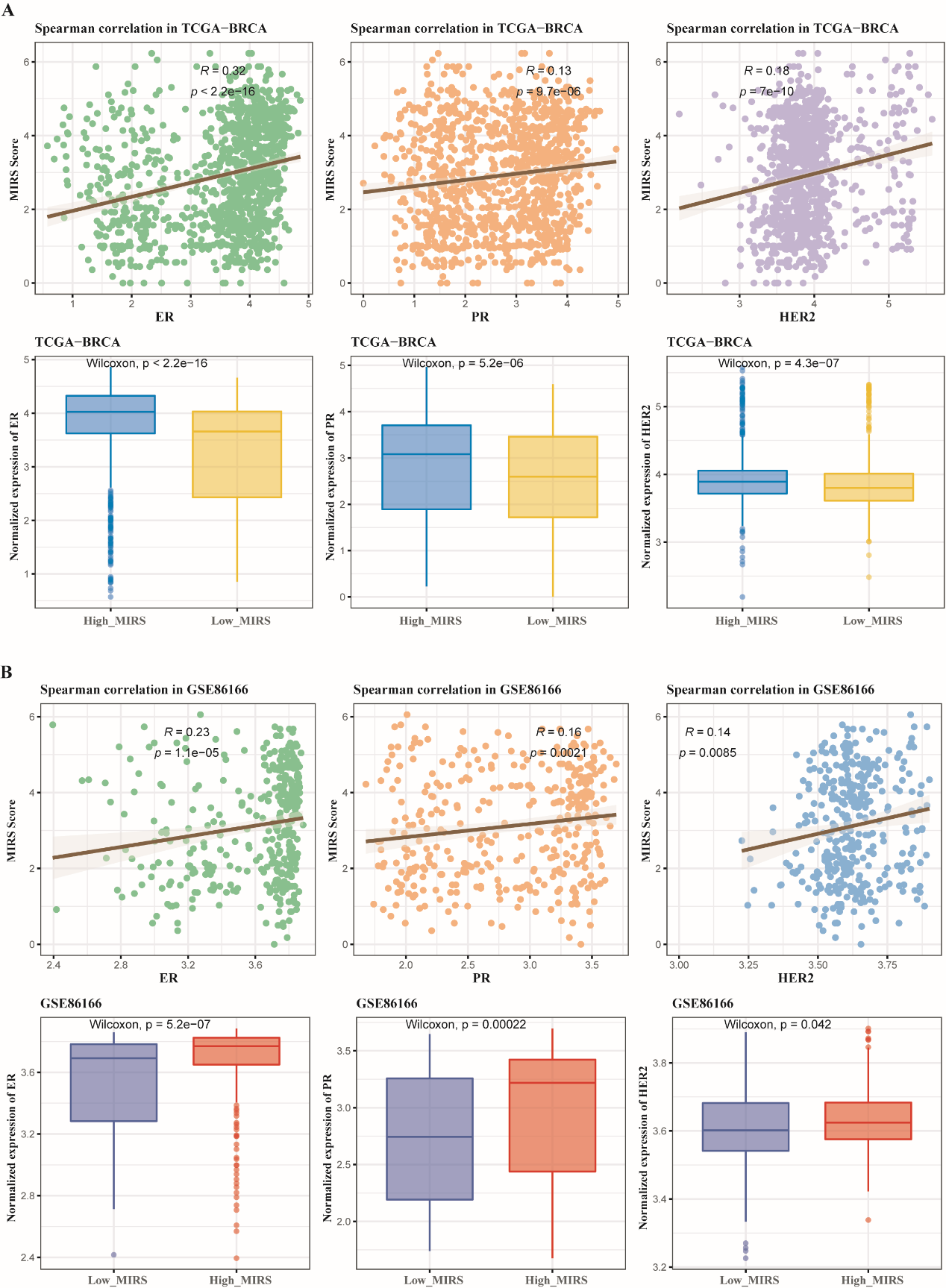

**Figure S15 Associations between MIRS and EP, PR, HER2. (A) Spearman correlation and Wilcox rank-sum test analyses in TCGA data. (B) Spearman correlation and Wilcox rank-sum test analyses in GSE86166 data.**

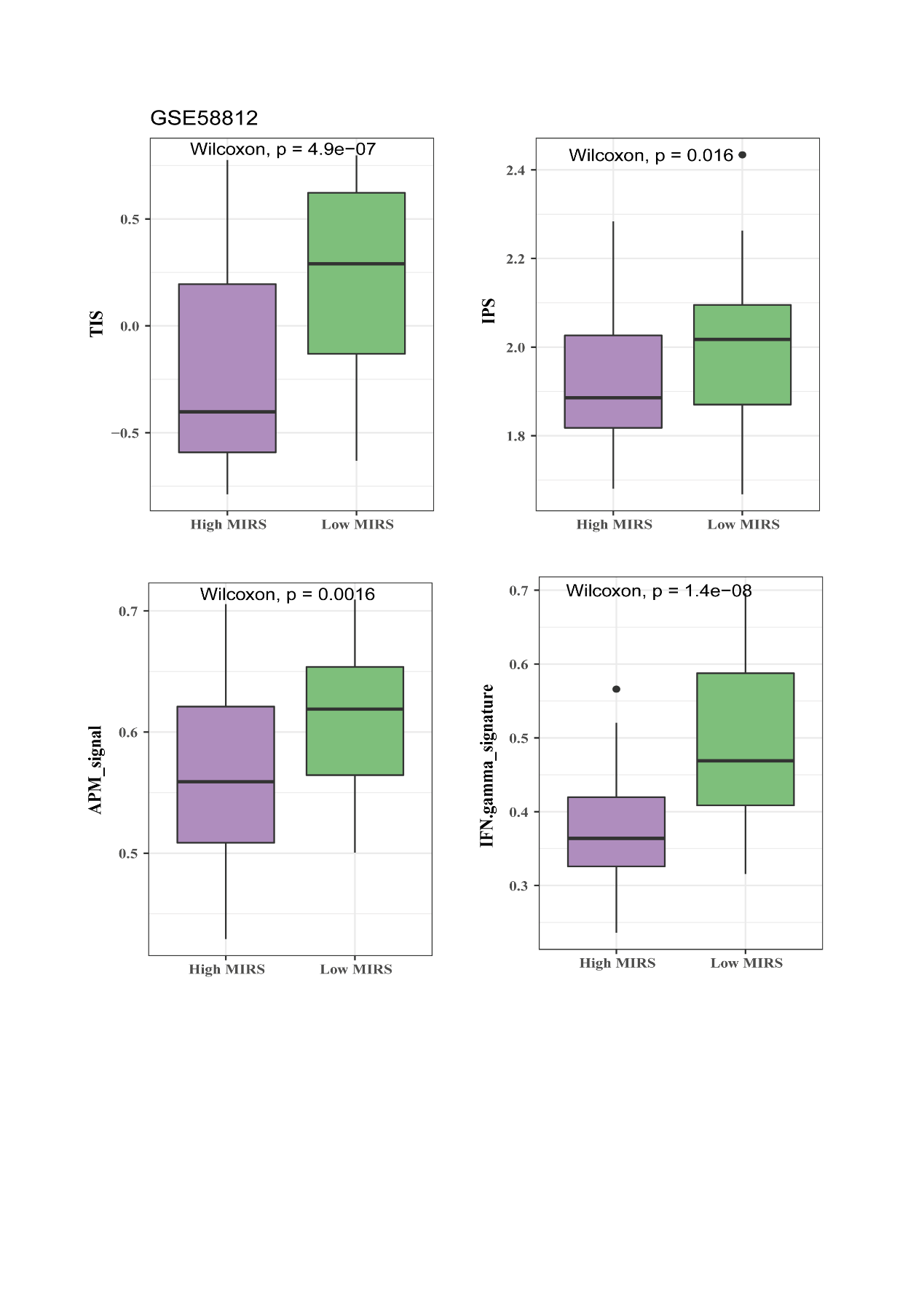

**Figure S16 Associations between MIRS and immunotherapy response in GSE58812.**

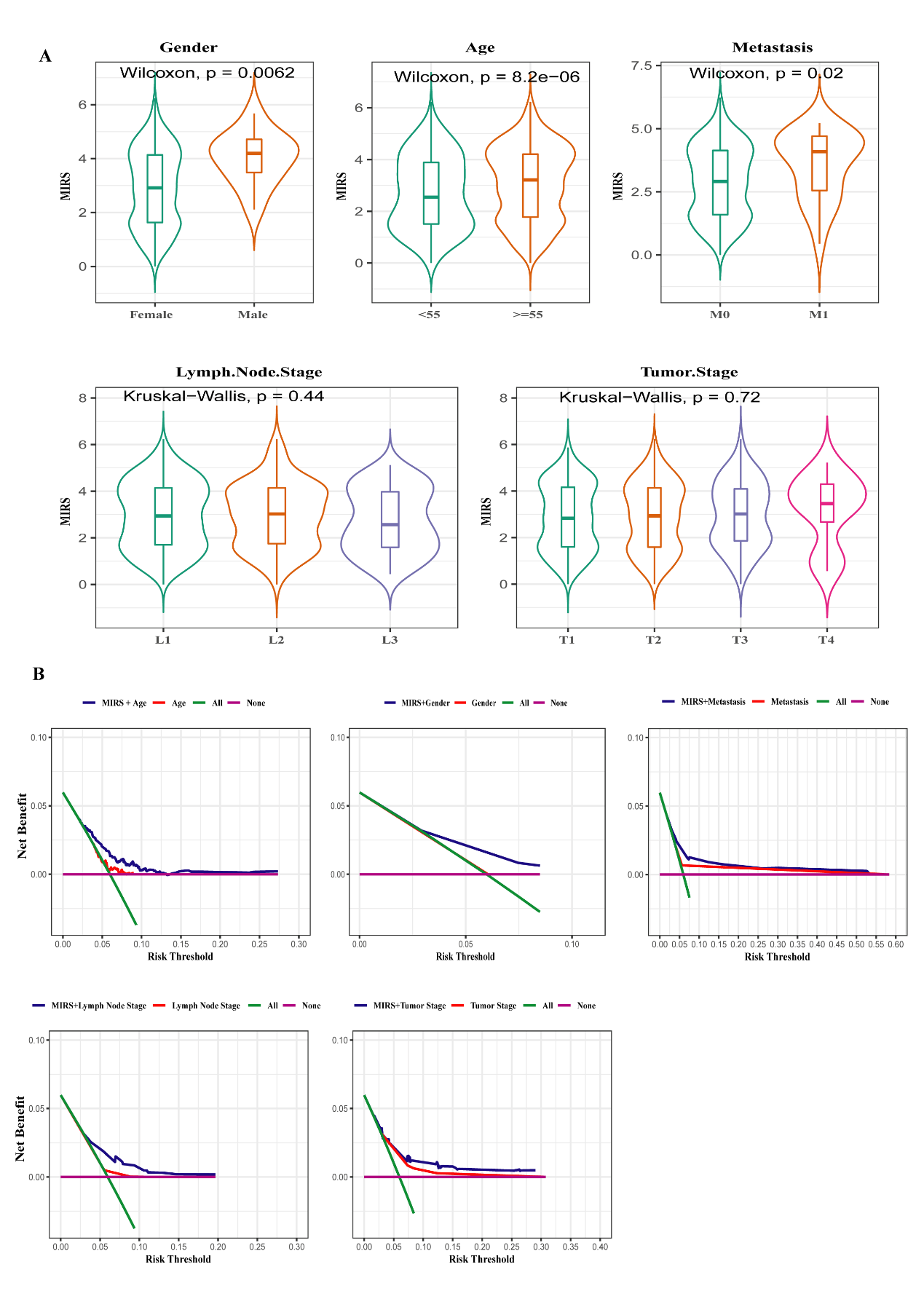

**Figure S17 Associations between MIRS and clinical index (A) Multiple comparison between MIRS and clinical features. (B) DCA analysis among MIRS, Age, Gender, Metastasis, Lymph node stage and Tumor stage.**

**Figure S18 Associations between MIRS and specific subtypes (A) KM survival analysis and ROC curves in TCGA data. (B) KM survival analysis and ROC curves in TCGA data.**
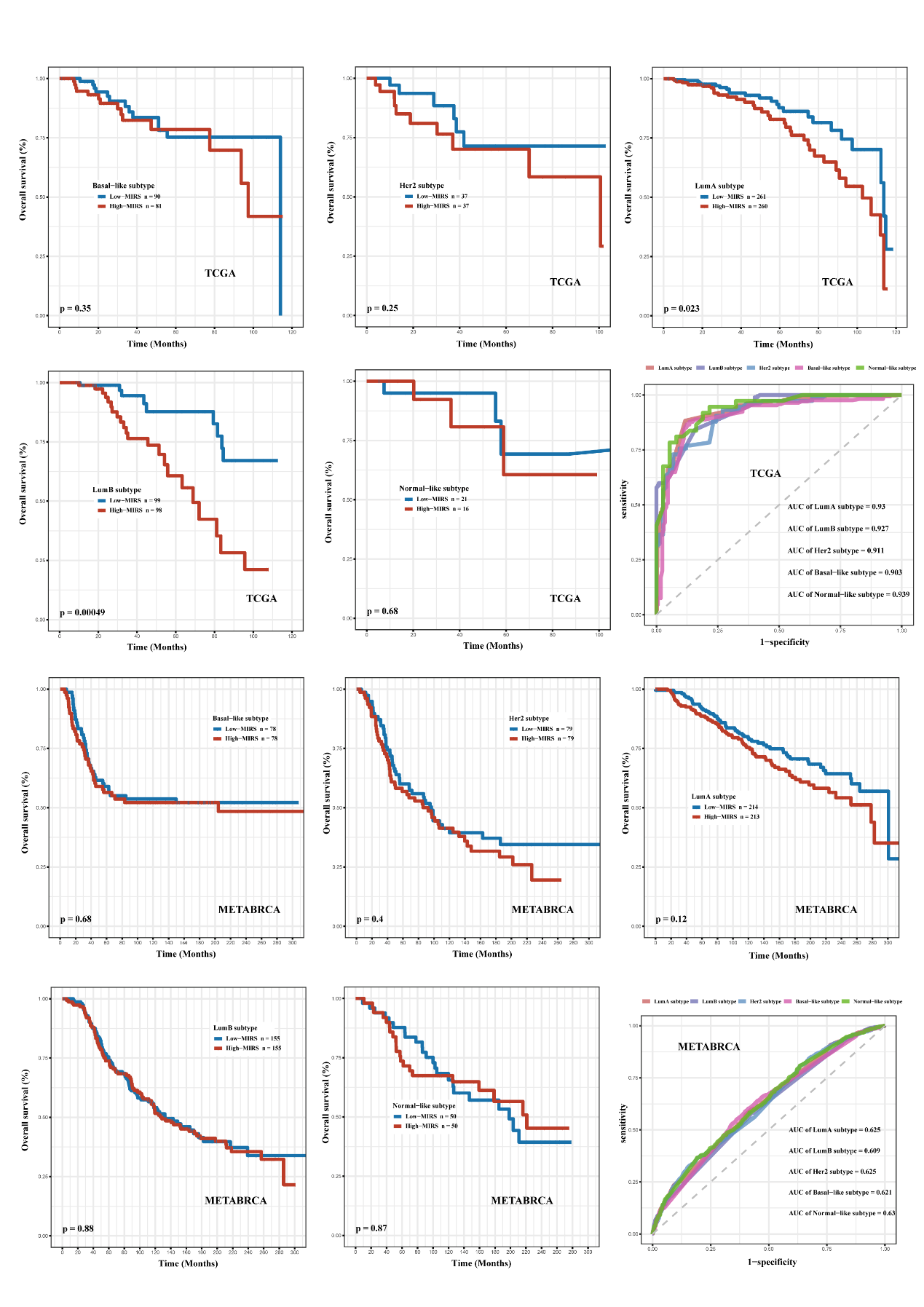

**Tables**

**Table S1. Data Information.**

| **Dataset** | **Metastasis** |  | **Primary** | **Total samples** |
| --- | --- | --- | --- | --- |
| GSE10893-GPL5325 | 12 |  | 6 | 27 |
| GSE3521-GPL887 | 17 |  | 58 | 75 |
| GSE86166-GPL15048 | 0 |  | 366 | 366 |
| GSE96058-GPL18573 | 0 |  | 3273 | 3273 |
| GSE20685-GPL570 | 0 |  | 327 | 327 |
| GSE20711-GPL570 | 0 |  | 88 | 88 |
| GSE58812-GPL570 | 0 |  | 107 | 107 |
| GSE9893-GPL5049 | 0 |  | 155 | 155 |
| GSE3143-GPL8300 | 0 |  | 158 | 158 |
| GSE42568-GPL570 | 0 |  | 104 | 104 |
| Liu et al data | 0 |  | 111 | 111 |
| TCGA-SKCM | 0 |  | 470 | 470 |
| METABRIC | 0 |  | 1980 | 1980 |
| TCGA-BRCA | 23 |  | 1165 | 1188 |

**Table S2. Data Utility**

| **Dataset** | **Clinical application** | **Statistical analysis** | **Metastatic**  **landscape** | **Immune**  **landscape** | **Therapy prediction** |
| --- | --- | --- | --- | --- | --- |
| **GSE10893-GPL5325** | 🗶 | 🗶 | ✓ | 🗶 | 🗶 |
| **GSE3521-GPL887** | 🗶 | 🗶 | ✓ | 🗶 | 🗶 |
| **GSE86166-GPL15048** | ✓ | ✓ | 🗶 | ✓ | 🗶 |
| **GSE96058-GPL18573** | ✓ | ✓ | 🗶 | ✓ | 🗶 |
| **GSE20685-GPL570** | ✓ | ✓ | 🗶 | 🗶 | ✓ |
| **GSE20711-GPL570** | ✓ | ✓ | 🗶 | ✓ | 🗶 |
| **GSE58812-GPL570** | ✓ | ✓ | 🗶 | ✓ | 🗶 |
| **GSE9893-GPL5049** | ✓ | 🗶 | 🗶 | 🗶 | 🗶 |
| **GSE3143-GPL8300** | ✓ | 🗶 | 🗶 | 🗶 | 🗶 |
| **GSE42568-GPL570** | ✓ | 🗶 | 🗶 | 🗶 | 🗶 |
| **Liu et al data** | ✓ | ✓ | 🗶 | 🗶 | ✓ |
| **TCGA-SKCM** | ✓ | ✓ | 🗶 | 🗶 | ✓ |
| **METABRIC** | ✓ | ✓ | ✓ | 🗶 | 🗶 |
| **TCGA-BRCA** | ✓ | ✓ | ✓ | ✓ | ✓ |

*Note:* Sign ✓ means that the data is being used, whereas sign 🗶 means that the data is not being used.

**Table S3 Genes were selected by Univariate cox proportional hazard analysis.**

| **Gene ID** | **Beta** | **HR (95% CI for HR)** | **Wald test** | **P value** |
| --- | --- | --- | --- | --- |
| APOA5 | 0.171 | 1.19(1.05-1.34) | 7.79 | 0.00526 |
| ASB2 | -0.157 | 0.855(0.761-0.961) | 6.93 | 0.00849 |
| CACNA1E | -0.15 | 0.861(0.755-0.981) | 5.06 | 0.0245 |
| CCL25 | -0.195 | 0.823(0.693-0.977) | 4.98 | 0.0257 |
| CD1A | -0.0763 | 0.927(0.862-0.995) | 4.34 | 0.0372 |
| CD1B | -0.145 | 0.865(0.785-0.952) | 8.71 | 0.00316 |
| CXCR3 | -0.107 | 0.899(0.824-0.98) | 5.79 | 0.0161 |
| FAM9C | 0.136 | 1.15(1.01-1.3) | 4.67 | 0.0307 |
| GPR55 | -0.137 | 0.872(0.791-0.96) | 7.8 | 0.00523 |
| GRAP2 | -0.112 | 0.894(0.805-0.993) | 4.38 | 0.0364 |
| IVL | 0.0778 | 1.08(1.02-1.15) | 6.21 | 0.0127 |
| LAX1 | -0.0857 | 0.918(0.845-0.997) | 4.09 | 0.0431 |
| PAGE5 | 0.113 | 1.12(1.01-1.24) | 5.07 | 0.0243 |
| TNFRSF8 | -0.136 | 0.873(0.776-0.982) | 5.15 | 0.0232 |
| WNT10A | -0.14 | 0.869(0.786-0.961) | 7.54 | 0.00605 |

**Table S4 Differentially expressed genes between the high immunity and low immunity groups in TCGA data.**

| **gene ID** | **Adj.p.value** | **logFC** | **gene ID** | **Adj.p.value** | **logFC** |
| --- | --- | --- | --- | --- | --- |
| 1-Dec | 0.002628 | -0.89349 | LOC116249 | 0.001267722 | -0.56127 |
| A2ML1 | 7.20E-15 | -0.68284 | LOC116403 | 1.89E-75 | -1.11871 |
| AADAC | 3.86E-12 | -0.76568 | LOC120712 | 3.32E-12 | -1.80888 |
| AADACL4 | 2.52E-06 | -1.57195 | LOC124977 | 1.27E-48 | -0.66378 |
| AADACP1 | 2.43E-11 | -0.62553 | LOC126277 | 6.15E-05 | 0.507753 |
| AANAT | 4.30E-06 | -0.54257 | LOC129287 | 0.000928726 | -0.56055 |
| AATK-AS1 | 0.003942 | 0.723072 | LOC129564 | 9.30E-06 | -0.85322 |
| ABCD2 | 3.25E-66 | -0.67427 | LOC135196 | 0.025350612 | 1.01358 |
| AC006129.2 | 3.22E-70 | -0.52419 | LOC138778 | 0.004256926 | 0.569927 |
| ACAP1 | 4.81E-92 | -0.52987 | LOC147864 | 3.75E-35 | -0.5627 |
| ACMSD | 0.000282 | 0.523999 | LOC148864 | 0.001598363 | -0.51097 |
| ACTL6B | 3.33E-05 | 3.634381 | LOC149803 | 1.49E-29 | -1.35255 |
| ACTL9 | 0.003962 | -1.15259 | LOC149915 | 8.81E-05 | 0.997256 |
| ACTR3BP2 | 0.035491 | -1.41458 | LOC154397 | 1.47E-79 | -0.58359 |
| ACY3 | 2.11E-33 | -0.6938 | LOC154544 | 9.42E-09 | -0.72845 |
| ADAM18 | 0.006864 | 1.521438 | LOC154576 | 1.29E-87 | -1.13808 |
| ADAM2 | 0.03957 | 0.61628 | LOC154866 | 0.000254622 | 0.738246 |
| ADAMDEC1 | 6.62E-68 | -0.60611 | LOC155456 | 0.003310666 | -1.15524 |
| ADGRE3 | 8.04E-23 | -1.04419 | LOC157462 | 6.85E-13 | -0.75259 |
| ADGRG3 | 4.63E-33 | -0.61387 | LOC158826 | 2.02E-12 | -0.76903 |
| ADGRG4 | 1.01E-08 | -1.11041 | LOC161464 | 0.003160194 | -0.71612 |
| ADGRG5 | 1.45E-69 | -0.60592 | LOC164378 | 0.000433753 | 1.455641 |
| ADH1A | 3.61E-08 | -0.55332 | LOC196745 | 1.96E-10 | -0.94667 |
| ADH4 | 3.19E-14 | -0.935 | LOC197414 | 4.33E-42 | -0.76636 |
| AGRP | 1.84E-06 | -0.54375 | LOC199676 | 1.30E-11 | -0.98944 |
| AGXT2 | 1.02E-06 | 1.259175 | LOC202365 | 8.41E-16 | -0.93588 |
| AHSP | 1.33E-11 | -1.0493 | LOC204855 | 1.05E-50 | -1.37592 |
| AICDA | 2.43E-51 | -1.6974 | LOC220000 | 1.85E-05 | -0.89382 |
| AIM2 | 8.21E-82 | -0.77985 | LOC221440 | 5.84E-79 | -1.07975 |
| AIPL1 | 1.39E-09 | -0.97182 | LOC221461 | 2.76E-33 | -0.90578 |
| AIRE | 2.35E-12 | -1.25597 | LOC221559 | 3.85E-07 | -0.98166 |
| ALX1 | 6.33E-05 | -0.50024 | LOC253271 | 3.53E-45 | -0.51724 |
| AMER3 | 0.003326 | 1.072313 | LOC256159 | 0.001198677 | 0.604101 |
| AMN | 1.11E-09 | -0.50134 | LOC283156 | 0.008205268 | -1.12011 |
| AMPD1 | 3.83E-35 | -0.91825 | LOC284798 | 0.000505935 | -0.55877 |
| AMTN | 2.17E-08 | -0.9163 | LOC285629 | 1.13E-09 | -0.65586 |
| AMY1A | 4.21E-10 | -0.5295 | LOC285768 | 0.007679675 | -0.61661 |
| ANGPTL6 | 4.85E-34 | -0.5465 | LOC286031 | 1.08E-23 | -0.57835 |
| ANKRD18DP | 2.14E-06 | -0.69268 | LOC387699 | 5.34E-09 | -0.70838 |
| ANKRD26P3 | 0.000129 | -0.73013 | LOC387860 | 0.000317994 | -0.57244 |
| ANKRD31 | 1.05E-11 | 0.540703 | LOC388117 | 0.014733286 | -0.93349 |
| ANKRD55 | 1.27E-40 | -0.76498 | LOC400760 | 1.30E-58 | -0.70351 |
| ANKUB1 | 2.58E-05 | -1.14423 | LOC402707 | 0.004275787 | 0.988556 |
| ANX14 | 0.000581 | -0.71063 | LOC4687 | 3.38E-86 | -0.59986 |
| AOX2P | 0.000107 | 0.616235 | LOC51180 | 0.030548812 | 0.665641 |
| APOA2 | 0.000848 | -0.97362 | LOC51269 | 0.001658904 | -0.58825 |
| APOA5 | 0.001545 | 0.676949 | LOC63189 | 1.02E-19 | -2.46307 |
| APOBEC3H | 2.29E-73 | -0.75401 | LOC63472 | 4.87E-76 | -0.9695 |
| APOC1P1 | 3.62E-11 | -0.92106 | LOC641864 | 1.46E-05 | -0.93332 |
| APOC4 | 3.41E-17 | -0.70959 | LOC644308 | 0.005991221 | -1.71976 |
| AQP7P3 | 0.000633 | -0.63737 | LOC644392 | 5.48E-05 | -0.64123 |
| ARL14 | 3.10E-12 | -0.87574 | LOC644781 | 2.54E-20 | -1.0441 |
| ARRDC5 | 6.23E-66 | -0.97266 | LOC647582 | 8.62E-05 | -0.56097 |
| ARSF | 0.000231 | -0.50745 | LOC648109 | 0.000187389 | -0.65914 |
| ART3 | 4.09E-28 | -0.86695 | LOC648271 | 1.86E-12 | -1.03147 |
| AS1 | 1.17E-62 | -0.50321 | LOC648473 | 2.23E-07 | -0.73331 |
| ASB15 | 8.72E-06 | 1.074495 | LOC648861 | 7.28E-62 | -1.12149 |
| ASB2 | 2.17E-75 | -0.53895 | LOC648953 | 7.66E-63 | -1.6653 |
| ASGR2 | 5.65E-67 | -0.69673 | LOC649015 | 0.001016215 | 0.770394 |
| ASIP | 1.91E-16 | -0.92527 | LOC649285 | 4.26E-06 | -1.07292 |
| ATP4A | 1.56E-07 | -0.67316 | LOC649464 | 2.38E-05 | -0.57465 |
| ATXN8OS | 0.000397 | 2.57203 | LOC650293 | 0.000179152 | Inf |
| AVPR1B | 4.55E-06 | -0.77309 | LOC653409 | 0.003915444 | -0.72028 |
| B3GALT2 | 5.09E-40 | -0.54126 | LOC654123 | 1.15E-82 | -0.52324 |
| BANK1 | 1.43E-71 | -0.74227 | LOC727730 | 5.94E-08 | -0.57237 |
| BARHL1 | 0.001576 | 1.255985 | LOC728943 | 0.013193305 | -0.60467 |
| BDK | 6.34E-08 | -0.64977 | LOC729609 | 0.011416922 | 0.66484 |
| BEND4 | 8.92E-55 | -1.28979 | LOC732373 | 1.95E-11 | -1.13083 |
| BET1P1 | 2.27E-16 | -0.85583 | LOC82212 | 0.000137465 | 0.522835 |
| BFSP2 | 7.02E-20 | -0.58443 | LOC89959 | 1.38E-63 | -1.04118 |
| BHR1 | 4.25E-64 | -1.16689 | LOC90262 | 4.29E-90 | -0.52701 |
| BLK | 2.10E-60 | -1.19738 | LOC90330 | 2.16E-05 | 0.971854 |
| BMP10 | 0.023675 | -0.5373 | LOC90374 | 7.70E-05 | -0.62651 |
| BMP3 | 2.70E-10 | -0.54787 | LOC90526 | 0.004771539 | -0.95125 |
| BPI | 4.51E-13 | -0.64579 | LOC90888 | 0.006470441 | 0.585646 |
| BPIFC | 0.003449 | -1.11226 | LOC91681 | 1.44E-05 | -0.61442 |
| BRD7P3 | 8.34E-06 | -0.63484 | LOR | 4.92E-09 | -0.89795 |
| BSPH1 | 0.027513 | #NAME? | LPAL2 | 7.64E-25 | -0.52464 |
| BTF3P11 | 0.001078 | 0.748764 | LPPR3 | 3.41E-16 | 0.518128 |
| BTLA | 3.31E-87 | -0.88719 | LRIT2 | 0.000314686 | -0.63572 |
| BTN1A1 | 4.20E-37 | -1.14551 | LRRC10 | 0.046948467 | 1.536567 |
| BTNL2 | 0.000284 | -0.59708 | LRRC18 | 1.80E-09 | -0.55829 |
| BTNL3 | 8.15E-08 | -0.83461 | LRRC7 | 2.72E-06 | -0.5154 |
| BTNL8 | 1.17E-31 | -1.07764 | LRRC74A | 0.000868228 | -0.53429 |
| C10ORF128 | 2.18E-59 | -0.55686 | LRRTM1 | 7.30E-05 | -0.63926 |
| C10orf50 | 1.81E-15 | -1.0142 | LTA | 2.07E-92 | -0.84917 |
| C10ORF62 | 0.002616 | 0.709101 | LTB | 2.44E-70 | -0.55948 |
| C10orf73 | 1.22E-06 | -0.94888 | LTK | 1.18E-35 | -0.62499 |
| C10ORF99 | 0.007001 | -0.72056 | LUZP4 | 0.005628149 | -0.61583 |
| C11ORF16 | 6.94E-06 | -0.55032 | LW | 1.28E-60 | -0.71928 |
| C11ORF21 | 3.11E-79 | -0.84089 | LY6D | 2.04E-11 | -0.66736 |
| C12ORF42 | 1.82E-23 | -0.98171 | LY6G6D | 0.015080481 | 0.896321 |
| C12ORF74 | 6.60E-15 | -0.87793 | LY86-AS1 | 1.57E-08 | -0.66967 |
| C12ORF77 | 1.33E-10 | -1.10321 | LY9 | 4.13E-94 | -0.71918 |
| C14ORF180 | 4.40E-06 | -0.50271 | LYPD2 | 0.004006022 | -0.89774 |
| C15ORF53 | 1.75E-51 | -1.8043 | LYPD8 | 1.01E-09 | -1.35351 |
| C15orf6 | 2.50E-11 | -0.5445 | LYPN | 3.96E-72 | -0.53589 |
| C16ORF90 | 0.001155 | -0.83919 | MAGEA4 | 0.000460136 | -0.74331 |
| C16ORF92 | 0.000268 | 1.192387 | MAGEB4 | 0.000634542 | -0.51065 |
| C17orf33 | 4.36E-07 | -0.76744 | MAL | 4.28E-70 | -0.83285 |
| C17ORF99 | 3.02E-14 | -0.88883 | MAP1LC3C | 1.81E-27 | -0.62469 |
| C1ORF140 | 9.54E-09 | -0.51473 | MAPT-AS1 | 4.40E-08 | 0.6018 |
| C1orf68 | 0.027011 | -1.48847 | MAPT-IT1 | 2.75E-10 | 0.572915 |
| C1QL2 | 3.15E-13 | -0.74435 | MARCO | 3.20E-34 | -0.73975 |
| C20orf138 | 0.011747 | 0.703387 | MAS1 | 7.90E-07 | -1.52064 |
| C20ORF141 | 5.56E-10 | -0.71055 | MASP1 | 3.59E-15 | -0.50325 |
| C20ORF166-AS1 | 7.42E-09 | -0.64881 | MC3R | 0.049531601 | -0.86274 |
| C20orf65 | 4.63E-60 | -1.8014 | MCCD1 | 4.94E-05 | 0.616043 |
| C22ORF34 | 2.32E-26 | -0.75426 | MCEMP1 | 5.38E-16 | -0.58813 |
| C2ORF83 | 0.000806 | -0.87606 | MDGA2 | 4.22E-06 | -0.52943 |
| C3ORF79 | 9.42E-05 | -2.03552 | MDS2 | 8.47E-45 | -1.52921 |
| C3ORF84 | 0.001203 | -1.36294 | MEP1A | 6.24E-36 | -1.186 |
| C4ORF26 | 9.67E-06 | -0.51808 | MGAT4C | 8.34E-05 | -0.50248 |
| C5ORF52 | 0.048382 | -0.91448 | MGC12916 | 2.97E-50 | -0.55588 |
| C5ORF58 | 5.04E-48 | -0.73254 | MGC27382 | 1.21E-05 | -0.99248 |
| C6orf95 | 6.66E-82 | -0.68668 | MGC35434 | 1.61E-13 | -0.5168 |
| C7ORF33 | 8.75E-10 | -1.01374 | MIR155HG | 1.27E-88 | -0.59414 |
| C7ORF62 | 0.000613 | 0.939782 | MIR7-3HG | 7.81E-05 | 2.62196 |
| C7ORF71 | 0.003836 | -0.57591 | MIXL1 | 1.01E-27 | -1.27345 |
| C7ORF72 | 0.00093 | -0.92034 | MMP12 | 1.01E-21 | -0.56844 |
| C8ORF86 | 2.79E-17 | 0.984546 | MMP20 | 1.68E-09 | -0.93193 |
| C9ORF139 | 2.19E-39 | -0.61676 | MMP27 | 1.66E-09 | -0.52664 |
| C9ORF170 | 1.76E-12 | -0.53754 | MMP8 | 4.77E-10 | -0.6543 |
| CA1 | 0.001747 | -0.90772 | MOGAT1 | 2.12E-05 | -0.50872 |
| CA4 | 7.03E-07 | -0.65834 | MPD1 | 1.90E-26 | -1.08848 |
| CA6 | 6.33E-05 | -0.59918 | MPO | 8.44E-31 | -0.57075 |
| CACNA1E | 2.24E-30 | -0.52925 | MPPED1 | 0.0008672 | 0.624589 |
| CACNG3 | 1.12E-23 | -2.29905 | MRAP | 4.61E-05 | -0.58303 |
| CAMK4 | 3.58E-55 | -0.50369 | MRGPRX2 | 7.21E-07 | -0.62388 |
| CARD17 | 1.69E-38 | -1.83302 | MROH2B | 0.016376361 | -0.53352 |
| CARD18 | 0.009576 | -0.84585 | MS4A1 | 3.94E-64 | -0.87111 |
| CARTPT | 1.18E-05 | 1.439823 | MS4A3 | 3.69E-09 | -0.97517 |
| CASP14 | 0.004451 | -0.52704 | MS4A8 | 1.15E-08 | 0.585016 |
| CASP5 | 2.47E-46 | -1.28908 | MUSK | 6.48E-22 | -1.49146 |
| CATSPER1 | 2.32E-28 | -0.53948 | MYBPC2 | 2.04E-42 | -1.19572 |
| CCDC141 | 1.29E-57 | -0.66287 | MYBPC3 | 1.37E-18 | -0.84016 |
| CCDC42 | 1.79E-25 | -1.20328 | MYF5 | 0.023746016 | 0.812087 |
| CCDC63 | 0.005543 | -0.63621 | MYH5~withdrawn | 0.012241832 | -0.55004 |
| CCDC83 | 5.00E-06 | 0.549403 | MYH6 | 7.32E-36 | -1.5671 |
| CCL1 | 2.03E-21 | -1.37441 | MYL2 | 4.27E-11 | -0.66527 |
| CCL13 | 8.00E-65 | -0.86952 | MYO7B | 8.57E-34 | -0.53262 |
| CCL16 | 1.82E-06 | -0.58045 | MYOC | 4.83E-06 | -0.55586 |
| CCL17 | 7.88E-54 | -0.80988 | MYOG | 7.17E-07 | 1.087675 |
| CCL18 | 9.71E-43 | -0.58128 | MYT1 | 4.61E-15 | 0.664551 |
| CCL19 | 2.39E-57 | -0.60122 | NAT16 | 0.003920177 | 0.641755 |
| CCL20 | 2.15E-23 | -0.51896 | NBPF22P | 3.02E-06 | 0.558924 |
| CCL23 | 1.54E-47 | -1.02451 | NBPF6 | 8.98E-07 | 0.549342 |
| CCL24 | 4.32E-26 | -1.92954 | NCF1B | 1.84E-79 | -0.56412 |
| CCL25 | 2.31E-26 | -1.05165 | NCR1 | 5.44E-47 | -1.71625 |
| CCL3L3 | 1.90E-13 | -0.53331 | NCR2 | 6.94E-13 | -1.71095 |
| CCL7 | 2.54E-19 | -0.86391 | NDST4 | 0.001062677 | 0.929312 |
| CCR3 | 4.14E-30 | -0.60811 | NEUROD1 | 0.039049905 | 2.272902 |
| CCR4 | 2.55E-68 | -0.60609 | NEUROD4 | 0.002162159 | 2.53518 |
| CCR7 | 3.16E-77 | -0.51634 | NKG7 | 9.84E-85 | -0.52808 |
| CCR8 | 1.63E-56 | -0.75373 | NKX1-2 | 2.45E-05 | -0.8681 |
| CD19 | 1.09E-64 | -1.32116 | NLC1-C | 8.01E-07 | -0.60106 |
| CD1A | 2.27E-28 | -0.68415 | NLRP10 | 1.30E-06 | -1.0244 |
| CD1B | 9.41E-58 | -1.31672 | NLRP11 | 4.39E-06 | -0.58048 |
| CD1C | 1.95E-49 | -0.62045 | NLRP13 | 0.04963756 | 0.550869 |
| CD200R1 | 2.99E-88 | -0.60073 | NLRP4 | 1.07E-10 | -0.9241 |
| CD200R1L | 1.00E-20 | -1.62786 | NLRP8 | 2.07E-05 | 0.699111 |
| CD226 | 3.98E-76 | -0.55589 | NMBR | 0.000495419 | 0.562108 |
| CD244 | 1.17E-86 | -0.70399 | NME8 | 7.06E-41 | -0.64185 |
| CD247 | 3.11E-94 | -0.54372 | NMRK2 | 0.000756063 | -0.88751 |
| CD27 | 3.23E-85 | -0.51691 | NOS1 | 4.00E-06 | -0.51164 |
| CD300E | 1.56E-32 | -0.96284 | NPFFR1 | 2.33E-23 | -1.51188 |
| CD300LD | 0.015452 | -0.703 | NPHS1 | 2.13E-06 | -0.6186 |
| CD38 | 3.56E-70 | -0.77685 | NPY | 0.01314234 | -0.71419 |
| CD3D | 9.59E-95 | -0.57689 | NR0B2 | 0.005581347 | 0.718921 |
| CD3G | 5.23E-87 | -0.76266 | NR2E1 | 0.000230448 | -0.63625 |
| CD40LG | 1.85E-85 | -0.7947 | NUGGC | 1.61E-76 | -0.94177 |
| CD5 | 5.98E-95 | -0.53666 | NUTM1 | 8.20E-06 | -0.56321 |
| CD7 | 3.17E-80 | -0.52811 | NXPE1 | 0.000221345 | -0.68896 |
| CD70 | 2.29E-52 | -0.66263 | NXPE4 | 2.51E-13 | -0.77874 |
| CD79A | 6.02E-64 | -0.63252 | OCSTAMP | 8.88E-06 | -0.74428 |
| CD79B | 1.31E-68 | -0.50683 | ODAM | 0.011167593 | -0.6845 |
| CD96 | 2.41E-92 | -0.51966 | OLAH | 7.64E-12 | -0.84817 |
| CDH17 | 1.37E-12 | -0.51129 | OPN1LW | 0.00119481 | -1.39834 |
| CDH19 | 0.000108 | -0.57295 | OPN5 | 0.00264755 | -0.72877 |
| CDHR1 | 3.11E-41 | -0.64023 | OPRK1 | 8.42E-06 | -0.51475 |
| CDIPT-AS1 | 1.81E-07 | 0.521144 | OPRM1 | 4.72E-05 | -1.57493 |
| CDX2 | 2.15E-06 | -0.70663 | OR10A1 | 0.009345103 | -1.20089 |
| CEACAM16 | 0.000522 | 1.013267 | OR10G2 | 0.000247235 | -0.65898 |
| CEACAM18 | 0.027513 | #NAME? | OR10H1 | 0.011191987 | 0.672728 |
| CEACAM20 | 1.28E-06 | -0.79759 | OR10J3 | 0.038141444 | 1.972128 |
| CEACAM22P | 3.75E-05 | -0.52578 | OR10Q1 | 7.81E-05 | -0.72165 |
| CEACAM3 | 9.52E-39 | -1.11176 | OR10V1 | 0.00031781 | -1.06929 |
| CEACAM4 | 2.37E-62 | -1.13663 | OR11L1 | 0.026587091 | 1.153463 |
| CEBPE | 5.14E-26 | -0.64678 | OR1D4 | 0.001750138 | -1.32749 |
| CELA1 | 4.22E-05 | -0.56411 | OR1E9P | 0.002096428 | -0.73369 |
| CELF3 | 5.37E-07 | 0.830156 | OR2B11 | 7.55E-08 | -1.0842 |
| CFC1B | 0.022582 | 0.802709 | OR2C5P | 0.031860446 | -0.5381 |
| CFHR1 | 7.00E-14 | -0.96312 | OR2S2 | 8.24E-05 | -0.99131 |
| CFHR5 | 0.005581 | 0.657413 | OR2T10 | 1.05E-07 | -2.43301 |
| CGA | 0.000613 | 0.725009 | OR2T3 | 0.013609611 | -0.80817 |
| CGB1 | 0.039085 | -1.05341 | OR2T5 | 0.009833228 | -0.99403 |
| CHIT1 | 5.23E-32 | -0.57067 | OR3A1 | 1.97E-06 | -0.74353 |
| CHRNB3 | 0.004253 | -0.62567 | OR3A2 | 1.22E-16 | -0.832 |
| CLC | 3.50E-16 | -1.5549 | OR3A8P | 7.23E-12 | -1.71071 |
| CLCA1 | 0.049774 | -0.61085 | OR4C6 | 2.75E-29 | -2.08379 |
| CLCA3P | 1.21E-05 | -1.09494 | OR4F12 | 0.033435002 | -1.34617 |
| CLDN16 | 2.18E-14 | -0.98831 | OR4K15 | 0.023175292 | 0.617891 |
| CLEC10A | 8.13E-70 | -0.66304 | OR51Q1 | 0.003275292 | 0.829759 |
| CLEC12A | 5.17E-77 | -0.70558 | OR52I1 | 0.006587925 | -0.90893 |
| CLEC12B | 6.14E-38 | -1.15494 | OR52K2 | 0.016829371 | -0.57614 |
| CLEC17A | 3.38E-52 | -1.17622 | OR52N1 | 0.028456849 | -0.5634 |
| CLEC4C | 4.30E-60 | -1.95388 | OR52N2 | 1.35E-05 | -0.59448 |
| CLEC4E | 2.54E-67 | -0.72899 | OR52N4 | 5.31E-37 | -1.14235 |
| CLEC4G | 7.26E-31 | -1.18211 | OR52N5 | 0.003545039 | -0.67588 |
| CLEC4M | 1.12E-07 | -0.71713 | OR56B1 | 5.52E-09 | -0.97868 |
| CLEC-6 | 1.19E-51 | -1.40004 | OR5B12 | 0.000150433 | -1.05183 |
| CLEC6A | 1.64E-56 | -1.80756 | OR5B2 | 5.83E-05 | -0.95284 |
| CLEC9A | 4.56E-42 | -0.8209 | OR5B21 | 6.16E-08 | -1.39249 |
| CLECL1 | 8.71E-77 | -0.68316 | OR6B2P | 3.06E-06 | 0.773814 |
| CLLU1OS | 1.15E-25 | -1.23695 | OR6B3 | 3.41E-05 | 0.880452 |
| CLNK | 8.06E-59 | -1.64706 | OR6C2 | 0.016702741 | -1.5165 |
| CLRN1 | 0.00241 | 0.668916 | OR6K3 | 0.001528072 | -1.06538 |
| CLRN1-AS1 | 7.74E-07 | 0.629146 | OR6N1 | 0.004227619 | -2.36114 |
| CMA1 | 3.40E-14 | -0.67035 | OR6S1 | 2.91E-09 | -2.18101 |
| CMYA4 | 8.05E-20 | -0.52513 | OR6W1P | 0.010502241 | 0.617884 |
| CNGB1 | 5.79E-12 | -0.77568 | OR7E156P | 0.013583882 | -0.64487 |
| CNR2 | 8.64E-59 | -1.42922 | OR7E24 | 0.01042303 | -0.67503 |
| CNTNAP4 | 1.55E-25 | -0.88893 | OR9G9 | 0.045335972 | -1.33848 |
| COL19A1 | 8.46E-34 | -1.37717 | OSTN | 0.007459386 | -0.89647 |
| COL4A3 | 8.46E-39 | -0.7305 | OTC | 0.002317446 | -0.77613 |
| COL6A5 | 1.18E-41 | -1.45205 | OTX2 | 0.007430729 | -0.68678 |
| CPA1 | 1.16E-05 | -0.58303 | OXT | 2.20E-05 | -0.53771 |
| CPA5 | 2.98E-14 | -0.70596 | P2RX1 | 3.96E-66 | -0.68842 |
| CPNE6 | 7.55E-15 | -1.43253 | P2RY10 | 4.44E-73 | -0.60634 |
| CPXCR1 | 0.004459 | -1.2968 | PABPC1L2A | 0.047925511 | 0.736988 |
| CR1 | 7.35E-76 | -0.73909 | PADI4 | 2.75E-21 | -0.82395 |
| CR2 | 1.96E-41 | -1.1369 | PADI6 | 0.001013833 | -0.66454 |
| CRCT1 | 0.004369 | -1.05371 | PAGE5 | 1.18E-08 | -0.59842 |
| CREB3L3 | 2.40E-38 | -1.22272 | PARP15 | 1.83E-82 | -0.75449 |
| CRLF2 | 5.00E-12 | -1.08566 | PATL2 | 2.14E-69 | -0.81945 |
| CRP | 8.50E-08 | -0.63286 | PAX5 | 1.57E-50 | -1.33172 |
| CRTAM | 8.33E-98 | -0.69764 | PBOV1 | 9.38E-08 | 0.679959 |
| CSAG2 | 2.27E-08 | -0.83576 | PCED1B-AS1 | 3.14E-89 | -0.64598 |
| CSAG3 | 3.09E-14 | -0.70808 | PDE6G | 1.43E-75 | -0.5646 |
| CSF2 | 4.91E-34 | -1.65495 | PF4 | 1.83E-07 | -1.29591 |
| CSMD3 | 0.001592 | 1.106014 | PF4V1 | 8.59E-06 | -0.78551 |
| CSN1S1 | 0.000306 | -0.75677 | PFIC2 | 2.28E-08 | -0.53211 |
| CSN3 | 7.92E-07 | -0.9297 | PGA4 | 0.001280257 | -0.50062 |
| CST13P | 0.005649 | 0.855505 | PGLYRP3 | 1.57E-05 | -0.82232 |
| CST9 | 9.49E-12 | 0.769569 | PGLYRP4 | 2.77E-12 | -0.72962 |
| CST9L | 2.80E-08 | 1.003307 | PGM5-AS1 | 4.53E-08 | -0.66087 |
| CT47B1 | 0.049382 | -2.06626 | PHF21B | 2.92E-06 | 0.611038 |
| CT83 | 2.94E-06 | -0.80076 | PHF2P1 | 0.010404715 | 1.2502 |
| CTRC | 5.64E-07 | -0.79341 | PHGR1 | 5.27E-08 | 0.701233 |
| CTSE | 2.06E-18 | -0.9051 | PHOX2B | 0.030027339 | -0.60017 |
| CXCL3 | 1.47E-20 | -0.67092 | PHP | 0.035582166 | 0.595313 |
| CXCL5 | 1.37E-17 | -0.68137 | PI3 | 1.63E-14 | -0.71745 |
| CXCL6 | 1.02E-20 | -0.76727 | PKHD1L1 | 4.44E-22 | -0.62916 |
| CXCR2P1 | 2.01E-46 | -0.77875 | PLAC8 | 4.77E-72 | -0.71539 |
| CXCR3 | 5.62E-92 | -0.58125 | PLEKHG7 | 1.05E-28 | -0.87459 |
| CXCR6 | 1.15E-90 | -0.52858 | PLET1 | 0.000829959 | -0.61485 |
| CXORF49B | 8.77E-09 | -0.91088 | PM20D1 | 7.87E-24 | -0.74762 |
| CXORF65 | 6.22E-63 | -1.79092 | PMCH | 1.66E-33 | -0.8289 |
| CXORF67 | 9.77E-11 | -0.78585 | PMP2 | 4.47E-05 | -0.70946 |
| CYP11B1 | 1.10E-05 | -1.12725 | PNLIPRP1 | 0.032931868 | 1.699467 |
| CYP2A7 | 9.87E-06 | 0.724698 | PNLIPRP2 | 6.40E-06 | 0.560225 |
| CYP2B | 7.31E-10 | 0.689138 | PNLIPRP3 | 0.001732959 | -0.74537 |
| CYP2C9 | 7.36E-07 | -0.55575 | PNMA5 | 1.41E-12 | -0.62706 |
| CYP7A1 | 3.82E-09 | -0.54446 | PNOC | 8.24E-75 | -1.03511 |
| D2S69E | 9.11E-68 | -0.57435 | POTEC | 0.01060682 | 0.533639 |
| DAD1P1 | 0.043698 | -0.53988 | POTED | 0.002814582 | 0.577835 |
| DAO | 2.54E-05 | -0.6558 | POTEG | 0.026824109 | 0.695675 |
| DAW1 | 4.76E-05 | -0.6723 | POTEH | 8.75E-07 | 0.730541 |
| DAZL | 2.45E-53 | -1.87523 | POU2AF1 | 1.46E-61 | -0.58539 |
| DCANP1 | 8.05E-91 | -0.93531 | POU3F1 | 5.84E-40 | -0.51773 |
| DCC | 1.85E-13 | -0.63091 | POU4F1 | 4.25E-15 | -0.6213 |
| DCSTAMP | 3.56E-33 | -0.53709 | PP12613 | 7.29E-18 | -1.82675 |
| DDX43 | 9.11E-13 | -0.50554 | PPP1R17 | 3.17E-06 | -0.83559 |
| DEFA1B | 3.86E-05 | -1.2299 | PPP1R2P9 | 4.68E-19 | -1.60342 |
| DEFA4 | 1.39E-10 | -2.4162 | PPP3R2 | 0.00063029 | -0.50577 |
| DEFB103A | 0.003063 | -2.00067 | PPY | 0.000345604 | -1.01004 |
| DEFB124 | 0.003371 | -1.10805 | PRDM13 | 0.014593214 | -0.53444 |
| DFNA48 | 4.07E-20 | -0.60462 | PRF1 | 8.54E-92 | -0.50299 |
| DFNB11 | 1.50E-09 | -0.58316 | PRG1 | 0.033266305 | -0.50854 |
| DFNB23 | 0.007819 | -0.51469 | PRKAG3 | 6.07E-08 | -0.71679 |
| DHRS9 | 5.64E-59 | -0.54418 | PRKCB | 4.34E-100 | -0.50459 |
| DIO3OS | 7.46E-13 | -0.63564 | PRKCQ | 3.74E-82 | -0.6617 |
| DIRC1 | 2.05E-20 | -0.61995 | PRL | 0.000816911 | -0.51491 |
| DKFZP434A014 | 5.13E-16 | -0.65298 | PRLH | 3.54E-05 | 1.097335 |
| DKFZp434J193 | 0.003385 | -0.6616 | PRM3 | 0.005632108 | -2.13355 |
| DKFZp547D155 | 0.000391 | 1.206823 | PRMT8 | 2.27E-05 | 0.739749 |
| DLX6-AS1 | 1.56E-08 | -0.68408 | PROK2 | 1.69E-24 | -1.12135 |
| DMRTA2 | 1.23E-06 | -0.65905 | PRR18 | 1.83E-07 | -0.53993 |
| DMRTC1 | 0.013819 | 1.097715 | PRR23B | 0.010993104 | -2.06057 |
| DNAI2 | 6.53E-08 | -0.59807 | PRR27 | 0.000205195 | -0.68913 |
| DNAJC5B | 7.73E-49 | -0.59452 | PRSS33 | 9.51E-06 | -0.56321 |
| DNASE1L3 | 2.08E-41 | -0.70939 | PRSS41 | 0.00278563 | -0.73352 |
| DNMT3L | 0.034426 | -0.66975 | PRTN3 | 9.85E-09 | 0.895174 |
| DNTT | 1.33E-08 | -1.1765 | PSAPL1 | 4.16E-05 | -1.00585 |
| DO | 1.59E-17 | -0.64591 | PSKH2 | 0.002367336 | 0.877564 |
| DPEP3 | 1.07E-18 | -0.87572 | PSMA8 | 7.72E-34 | -1.19084 |
| DPPA3 | 1.51E-07 | -1.35029 | PSTPIP1 | 1.13E-92 | -0.50463 |
| DPPA4 | 2.17E-51 | -1.60531 | PTPN20A | 0.033908651 | 0.718636 |
| DPPA5 | 0.040296 | -0.58188 | PWAR4 | 0.017347921 | 1.375336 |
| DPYS | 1.04E-31 | -0.93164 | PWRN2 | 0.000105737 | -1.50041 |
| DRD3 | 4.92E-05 | -1.33627 | PXT1 | 2.50E-07 | -0.51291 |
| DRGX | 0.000111 | -1.89574 | PYHIN1 | 1.32E-91 | -0.78622 |
| DSCR4 | 0.008078 | -1.39499 | PYY | 6.34E-09 | 0.783413 |
| DTHD1 | 1.28E-56 | -1.13065 | RAB9BP1 | 0.039049905 | -3.32083 |
| DUX4L6 | 0.008993 | 2.043886 | RAET1L | 1.08E-09 | -0.69852 |
| ECEL1 | 3.48E-26 | -0.53083 | RAX | 0.000998429 | -0.65883 |
| ED2 | 7.37E-16 | -0.57967 | RDH8 | 3.34E-05 | -0.7043 |
| EIF4E1B | 0.006851 | 0.526045 | RETN | 8.71E-08 | -0.6447 |
| ELAVL3 | 0.000334 | 0.927922 | RFPL4 | 0.000109711 | -0.72648 |
| EMR4 | 2.44E-34 | -0.66427 | RFPL4B | 4.97E-13 | -0.72113 |
| ENAM | 7.13E-08 | -0.60664 | RGL4 | 3.55E-78 | -0.78807 |
| ENPP7 | 1.19E-05 | -0.56437 | RGR | 0.001924866 | -0.53479 |
| ENTHD1 | 1.77E-50 | -0.79239 | RGS13 | 3.65E-29 | -0.61779 |
| EOMES | 2.75E-77 | -0.74146 | RGS20 | 2.31E-11 | -0.51339 |
| EPB42 | 6.83E-05 | -0.71018 | RLBP1 | 0.000380334 | -0.65779 |
| EQTN | 2.97E-05 | -0.82193 | RLN3 | 0.000173596 | -0.79026 |
| ERC2-IT1 | 0.00509 | -0.97943 | RLTPR | 3.61E-45 | -0.51105 |
| ERN2 | 2.95E-08 | -0.72326 | RMST | 0.000669919 | 0.607655 |
| ESX1 | 0.005214 | -0.52205 | RNASE3 | 2.08E-12 | -0.86021 |
| FABP7 | 3.73E-09 | -0.59313 | RNF35 | 2.07E-35 | -0.70092 |
| FAM129C | 9.76E-48 | -1.05033 | ROS1 | 1.57E-07 | -0.70427 |
| FAM150A | 0.046998 | -0.6204 | RPTN | 0.021884551 | -0.76686 |
| FAM150B | 1.44E-16 | -0.65324 | RTP1 | 1.79E-08 | -0.92222 |
| FAM151A | 3.39E-20 | -1.31436 | RTP3 | 0.00011071 | -1.28643 |
| FAM153A | 1.56E-18 | -0.71375 | RTP5 | 4.32E-54 | -1.695 |
| FAM159A | 2.81E-52 | -0.56705 | RUFY4 | 1.46E-40 | -0.68697 |
| FAM179A | 1.57E-40 | -0.74468 | RUNX1-IT1 | 8.27E-29 | -1.20308 |
| FAM180B | 7.58E-14 | -0.62616 | RXFP1 | 1.33E-07 | -0.51768 |
| FAM187B | 1.21E-05 | -0.63072 | S100A12 | 1.15E-26 | -1.08195 |
| FAM19A1 | 8.72E-24 | -0.74109 | S100A7 | 8.55E-08 | -0.57217 |
| FAM215A | 9.52E-18 | -0.85617 | S100A7A | 2.99E-06 | -0.67453 |
| FAM216B | 8.79E-14 | -0.76202 | S100G | 0.000710415 | 0.947459 |
| FAM222A-AS1 | 5.35E-11 | 0.509855 | S100Z | 1.65E-22 | -0.64271 |
| FAM25A | 0.002409 | 0.555065 | S1PR4 | 3.81E-88 | -0.6377 |
| FAM30A | 2.49E-59 | -0.9369 | SAA3P | 0.001790079 | -1.29484 |
| FAM47A | 0.003529 | 0.68139 | SAA4 | 3.88E-16 | -0.61752 |
| FAM71A | 0.000192 | -0.78232 | SAMD3 | 4.30E-89 | -0.68326 |
| FAM71B | 0.001924 | -1.52938 | SAMD7 | 1.07E-06 | -0.96711 |
| FAM71C | 1.42E-06 | 0.81526 | SCA12 | 2.09E-59 | -0.56607 |
| FAM74A3 | 0.009709 | -0.57577 | SCA23 | 0.000863452 | -0.56049 |
| FAM92B | 1.81E-19 | -0.79433 | SCARA5 | 2.02E-21 | -0.55548 |
| FAM9C | 2.25E-07 | -0.57822 | SCEL | 1.18E-13 | -0.95237 |
| FASLG | 9.05E-81 | -0.76764 | SCIMP | 2.39E-96 | -0.55402 |
| FBXL21 | 0.002362 | 1.026625 | SCML4 | 4.44E-85 | -0.7603 |
| FCA/MR | 2.41E-31 | -1.50307 | SCRT1 | 0.002173958 | 0.565049 |
| FCAR | 2.25E-35 | -1.03278 | SDC4P | 0.041983706 | -1.05081 |
| FCER2 | 5.51E-58 | -1.5869 | SEL1L2 | 0.04092887 | -0.55976 |
| FCN1 | 6.89E-77 | -0.72982 | SEMG1 | 1.11E-05 | -1.46049 |
| FCRL1 | 1.27E-59 | -1.49668 | SEMG2 | 0.019160987 | -1.08883 |
| FCRL2 | 8.68E-60 | -1.24222 | SERPINA10 | 0.012272686 | 0.585511 |
| FCRL3 | 4.92E-74 | -1.0348 | SERPINA6 | 3.52E-06 | 0.575564 |
| FCRL4 | 6.39E-40 | -1.20914 | SERPINB10 | 8.85E-05 | -0.75775 |
| FCRL5 | 1.41E-59 | -0.79749 | SERPINB4 | 1.59E-05 | -0.90831 |
| FCRL6 | 9.37E-70 | -0.68159 | SFTPB | 3.07E-12 | -0.62067 |
| FCRLA | 6.36E-71 | -0.91878 | SH2D1A | 5.52E-90 | -0.68487 |
| FDCSP | 3.13E-33 | -0.93151 | SH2D1B | 4.05E-56 | -0.73231 |
| FERD3L | 0.018314 | -0.70713 | SHD | 7.01E-42 | -1.45407 |
| FFAR3 | 1.11E-32 | -0.66745 | SHISA3 | 2.57E-22 | -0.67176 |
| FGF3 | 0.003041 | 0.902909 | SIGLEC12 | 5.07E-24 | -0.82252 |
| FGFBP1 | 3.59E-15 | -0.75985 | SIGLEC23P | 1.57E-05 | -1.25782 |
| FGFBP2 | 2.37E-29 | -0.59214 | SIT1 | 2.69E-96 | -0.69593 |
| FIGLA | 0.001521 | -0.50775 | SLA2 | 1.47E-91 | -0.60278 |
| FIH | 2.08E-12 | -0.77254 | SLAMF1 | 1.12E-97 | -0.7588 |
| FKHL4 | 6.71E-11 | -0.86692 | SLAMF6 | 1.17E-95 | -0.59198 |
| FLJ20087 | 7.09E-23 | -0.53445 | SLC10A2 | 0.004682683 | -1.02938 |
| FLJ20261 | 0.001302 | 0.595361 | SLC12A3 | 1.75E-63 | -1.03087 |
| FLJ21477 | 8.45E-13 | -0.54273 | SLC13A1 | 0.034768623 | 0.634129 |
| FLJ36131 | 0.008078 | -0.73975 | SLC15A1 | 1.24E-08 | -0.54741 |
| FLJ37300 | 7.91E-30 | -0.59619 | SLC17A3 | 8.05E-06 | -0.53879 |
| FLJ41941 | 6.76E-10 | -0.70006 | SLC18A1 | 2.42E-28 | -0.87844 |
| FLJ45079 | 5.10E-12 | -0.81631 | SLC22A10 | 0.00023573 | 0.689944 |
| FLJ90231 | 7.77E-11 | -0.66313 | SLC22A12 | 0.000165277 | -1.0326 |
| FMO6P | 2.75E-15 | -0.9368 | SLC22A2 | 4.01E-07 | -0.61822 |
| FOLR3 | 2.68E-09 | -0.98614 | SLC22A24 | 3.21E-06 | 1.110018 |
| FOXB1 | 2.32E-19 | -1.83158 | SLC24A4 | 6.78E-79 | -0.75757 |
| FOXD4L3 | 1.74E-08 | -0.88443 | SLC26A9 | 1.03E-30 | -0.88715 |
| FOXN3-AS2 | 0.012162 | -0.84967 | SLC28A2 | 0.000393421 | 0.999782 |
| FRG2B | 0.010484 | 0.93575 | SLC2A2 | 0.028094826 | -0.66935 |
| FRMPD4 | 5.70E-09 | -0.55599 | SLC2A7 | 1.90E-05 | -0.9586 |
| FSCB | 0.016257 | 0.923405 | SLC32A1 | 4.51E-40 | -2.53667 |
| FSHB | 0.038432 | 1.735723 | SLC36A2 | 9.39E-08 | -0.84712 |
| FSP | 9.45E-29 | -0.6699 | SLC36A3 | 0.032349637 | -0.53039 |
| FTMT | 5.60E-05 | -0.51917 | SLC39A12 | 1.84E-09 | -0.69715 |
| FUT7 | 9.97E-83 | -0.77064 | SLC5A7 | 0.0072494 | 0.620853 |
| FXYD7 | 6.02E-46 | -0.96685 | SLC6A2 | 1.81E-15 | -0.8334 |
| GABBR2 | 5.97E-08 | -0.61565 | SLC6A7 | 2.04E-15 | -0.56977 |
| GABRA1 | 0.034141 | 0.5299 | SLC8A3 | 4.39E-22 | -0.5419 |
| GALR1 | 0.003531 | -0.55674 | SLC9A4 | 2.35E-07 | -0.58592 |
| GATSL2 | 0.040142 | 0.547115 | SLC9C2 | 4.20E-07 | -0.71187 |
| GBP6 | 3.43E-36 | -0.60006 | SLCO5A1 | 1.76E-53 | -0.72043 |
| GBP7 | 7.80E-08 | -0.66137 | SLEB2 | 5.94E-80 | -0.72713 |
| GCSAML-AS1 | 7.25E-14 | -0.70637 | SLFN12L | 1.47E-76 | -0.76435 |
| GDA | 4.53E-11 | -0.66257 | SLFN14 | 3.61E-51 | -1.43688 |
| GDAP1L1 | 1.32E-05 | 0.519079 | SLITRK2 | 9.86E-22 | -0.54594 |
| GDF7 | 5.77E-09 | -1.22672 | SMPX | 0.000317004 | -0.70804 |
| GFI1 | 1.54E-81 | -0.57217 | SMR3A | 0.009189362 | -0.86798 |
| GFRAL | 0.013118 | -0.70916 | SNORD115-26 | 0.030461261 | 1.026443 |
| GH1 | 1.82E-22 | -1.32877 | SNX20 | 1.32E-103 | -0.53294 |
| GHRHR | 0.028719 | -0.83252 | SOX14 | 0.002482557 | -1.27474 |
| GHRL | 6.98E-41 | -0.63762 | SPACA3 | 1.63E-07 | -0.94218 |
| GKN2 | 0.003448 | -0.50365 | SPATA22 | 3.07E-11 | -0.64815 |
| GLIS3-AS1 | 6.79E-05 | -0.57702 | SPATA3 | 0.034517257 | -0.63372 |
| GLP2R | 0.000189 | -0.64429 | SPATA4 | 9.93E-10 | 0.538945 |
| GLT1D1 | 5.64E-54 | -0.77575 | SPATA8 | 2.51E-13 | -1.01832 |
| GLYAT | 2.18E-06 | -0.56571 | SPATC1 | 2.44E-37 | -0.60476 |
| GML | 7.96E-10 | -1.89013 | SPATS1 | 0.001989336 | -0.55838 |
| GNG8 | 1.34E-09 | -1.44599 | SPIB | 5.60E-68 | -0.93099 |
| GOLGA6L1 | 0.007838 | -0.8535 | SPIC | 8.30E-41 | -1.86026 |
| GOLGA8G | 0.010894 | -0.51083 | SPINK2 | 1.46E-14 | -0.64359 |
| GP1BA | 2.44E-45 | -0.59947 | SPINK4 | 5.77E-08 | 0.996621 |
| GP5 | 1.08E-20 | -0.54854 | SPINK6 | 0.000468081 | -1.16635 |
| GPR12 | 2.16E-10 | -0.58718 | SPLA2S | 1.94E-76 | -0.9305 |
| GPR139 | 2.20E-07 | 0.980012 | SPP2 | 0.039616029 | -0.68844 |
| GPR142 | 4.95E-10 | -0.80403 | SPPL2C | 0.00047043 | 0.636093 |
| GPR148 | 0.000971 | 1.717032 | SPRR1B | 0.001118246 | -0.6937 |
| GPR15 | 1.69E-28 | -1.07254 | SPRR2A | 2.79E-05 | -0.99724 |
| GPR150 | 2.86E-18 | -1.16578 | SPRR2B | 0.048177803 | -1.23402 |
| GPR171 | 1.36E-85 | -0.7484 | SPRR2C | 0.007179018 | -1.17409 |
| GPR174 | 4.56E-73 | -0.9373 | SPRR2D | 7.78E-05 | -1.09902 |
| GPR18 | 6.37E-80 | -0.73309 | SPRR2E | 0.005423221 | -1.008 |
| GPR182 | 4.23E-10 | -0.52611 | SPTA1 | 3.38E-37 | -0.65075 |
| GPR25 | 8.29E-41 | -1.57031 | SPX | 1.33E-08 | -0.7394 |
| GPR28 | 1.52E-31 | -1.01992 | SSTR3 | 3.65E-62 | -1.657 |
| GPR31 | 1.79E-41 | -1.12534 | SSTR5-AS1 | 0.001653679 | 1.041741 |
| GPR55 | 2.93E-68 | -0.75934 | ST18 | 3.89E-35 | -0.61506 |
| GRAP2 | 1.51E-86 | -0.50781 | STAB2 | 5.69E-11 | -0.55965 |
| GRIA2 | 1.10E-05 | 0.502949 | STAP1 | 4.95E-71 | -0.8327 |
| GRIN2B | 2.14E-07 | -0.52047 | STAT4 | 1.18E-88 | -0.51243 |
| GSTA5 | 0.006978 | -0.56957 | STH | 0.000435508 | 0.512007 |
| GSX2 | 0.004898 | -0.87655 | STRA8 | 1.52E-05 | -0.79426 |
| GTSF1 | 1.76E-59 | -1.02919 | SUCNR1 | 4.89E-37 | -0.54005 |
| GUCY2F | 0.00174 | -0.82287 | SULT1E1 | 0.000438764 | -0.54431 |
| GYPE | 1.47E-40 | -0.51425 | SYL | 0.000864583 | -1.13905 |
| GZMA | 3.85E-90 | -0.52673 | T | 9.93E-05 | -0.71047 |
| GZMB | 4.25E-79 | -0.78462 | TAC3RL | 0.000233635 | 0.61585 |
| GZMH | 3.67E-72 | -0.63283 | TAS2R60 | 0.000840962 | -0.69143 |
| GZMK | 3.22E-82 | -0.58902 | TBC1D10C | 3.92E-87 | -0.502 |
| GZMM | 6.23E-69 | -0.76662 | TBLYM | 2.71E-91 | -0.80675 |
| H1FNT | 0.003979 | -0.51127 | TCF23 | 0.000289235 | -0.76117 |
| HAO1 | 6.94E-06 | -0.74291 | TCHHL1 | 0.00712748 | -0.79083 |
| HAS2-AS1 | 3.04E-22 | -0.52601 | TCL1A | 4.92E-65 | -1.35076 |
| HAVCR1 | 5.99E-10 | -0.61065 | TCL6 | 1.04E-16 | -0.64938 |
| HBBP1 | 4.98E-06 | -1.32114 | TDGF1P3 | 1.12E-05 | -0.55188 |
| HBD | 6.26E-12 | -0.76163 | TENB2 | 0.003049722 | -0.5093 |
| HCL3 | 2.33E-06 | -0.58983 | TEPP | 2.04E-09 | -0.59656 |
| HEATR9 | 1.39E-36 | -1.51233 | TESPA1 | 8.73E-90 | -0.64816 |
| HEMGN | 6.11E-27 | -1.36144 | TEX101 | 0.003951645 | -0.52471 |
| HEPACAM | 9.03E-08 | -0.53652 | TEX11 | 9.74E-44 | -0.95813 |
| HGC6.3 | 1.32E-11 | -0.59308 | TEX26 | 0.001676194 | -0.92884 |
| HHLA2 | 1.61E-23 | -1.02891 | TGM7 | 1.11E-05 | -0.8353 |
| HKDC1 | 1.36E-29 | -0.87755 | THEMIS | 1.65E-81 | -0.70009 |
| HLA-DOB | 1.79E-68 | -0.531 | TIFAB | 1.85E-90 | -1.10059 |
| HLA-DPB2 | 6.58E-54 | -0.68376 | TIGIT | 1.64E-90 | -0.6285 |
| HMHB1 | 5.16E-09 | -0.89021 | TIMD4 | 1.02E-48 | -0.94366 |
| HMP19 | 4.72E-07 | 0.58178 | TKTL1 | 3.04E-34 | -1.12746 |
| HMSD | 3.53E-34 | -1.27077 | TLR10 | 1.93E-67 | -0.60766 |
| HNF1B | 3.29E-05 | -0.6046 | TLX3 | 4.84E-06 | -1.02388 |
| HOMG2 | 4.54E-54 | -1.02943 | TMCO5A | 0.00139405 | -1.07596 |
| HORMAD1 | 4.03E-11 | -0.77055 | TMEM14E | 1.29E-08 | -0.6781 |
| HPCAL4 | 7.54E-25 | -0.57779 | TMEM150B | 1.64E-72 | -0.81989 |
| HPR | 3.24E-09 | -0.8796 | TMEM156 | 1.08E-59 | -0.52632 |
| HPYR1 | 0.005761 | 0.618273 | TMEM171 | 1.43E-18 | -0.52371 |
| HSCDIEL | 4.11E-50 | -0.64143 | TMEM235 | 0.038169723 | -0.55243 |
| HSD11B1 | 4.10E-69 | -0.52651 | TMEM236 | 7.68E-29 | -0.5159 |
| HSD17B13 | 1.61E-14 | -0.53725 | TMEM257 | 0.026747619 | -0.58328 |
| HSF5 | 1.84E-49 | -1.06416 | TMEM82 | 0.037050064 | 0.68467 |
| HSPC062 | 1.98E-88 | -0.904 | TMEM95 | 0.020024224 | -0.57066 |
| HTN1 | 0.001133 | -1.00577 | TMIGD2 | 1.79E-62 | -1.14447 |
| HTR1E | 2.59E-09 | 0.96417 | TNFRSF13B | 1.53E-52 | -1.15767 |
| HTR3A | 1.11E-20 | -0.83222 | TNFRSF13C | 7.27E-40 | -0.85067 |
| HTR3C | 0.003311 | -0.55864 | TNFRSF17 | 2.74E-53 | -0.85033 |
| HTR3E | 7.25E-07 | -0.98255 | TNFRSF8 | 1.30E-88 | -0.57131 |
| HTRA4 | 2.52E-54 | -0.72238 | TNFRSF9 | 1.34E-80 | -0.80876 |
| HYMAI | 1.92E-10 | -0.50616 | TNFSF12-TNFSF13 | 2.20E-60 | -0.50504 |
| HYPM | 0.004177 | 0.637112 | TNFSF18 | 8.78E-23 | -0.82695 |
| ICOS | 1.63E-83 | -0.74903 | TOX | 1.64E-76 | -0.53236 |
| IDDM12 | 8.50E-83 | -0.69192 | TP53TG5 | 7.54E-05 | 0.707748 |
| IDO | 1.69E-71 | -0.60183 | TPTE2 | 6.13E-15 | -0.91854 |
| IDO2 | 9.30E-63 | -1.38925 | TRABD2A | 9.57E-77 | -0.63409 |
| IFNB1 | 1.09E-10 | -0.756 | TRAPPC3L | 7.16E-36 | -0.96395 |
| IFNE | 6.06E-08 | -0.50296 | TREML1 | 7.97E-51 | -0.76116 |
| IFNG | 1.17E-66 | -1.44767 | TREML4 | 0.007786562 | -0.63429 |
| IFNK | 0.022565 | -0.95914 | TRH | 4.26E-11 | 0.657003 |
| IFNL1 | 2.94E-15 | -0.75862 | TRIM10 | 7.05E-08 | -0.91551 |
| IFNL2 | 5.36E-05 | -0.71617 | TRIM42 | 0.01079855 | 1.352979 |
| IFNL3 | 0.000608 | -0.73519 | TRIML1 | 0.008844691 | -0.72838 |
| IGLL1 | 8.25E-15 | -1.37225 | TRIML2 | 1.09E-10 | -0.75288 |
| IKZF3 | 5.79E-45 | -0.60529 | TRPC2 | 1.15E-13 | -0.55874 |
| IL10 | 6.84E-72 | -0.55075 | TRPM1 | 0.000400331 | -0.72919 |
| IL12A | 3.39E-23 | -0.53479 | TRPM8 | 4.47E-18 | -0.71744 |
| IL12RB1 | ####### | -0.55483 | TRYX3 | 0.004945495 | -1.15311 |
| IL12RB2 | 7.88E-42 | -0.63177 | TSGA10IP | 1.83E-15 | -0.70971 |
| IL13 | 4.51E-13 | -1.0958 | TSHR | 1.42E-46 | -0.73261 |
| IL17F | 7.10E-09 | -2.04947 | TSKS | 2.03E-22 | -0.83585 |
| IL18RAP | 1.80E-83 | -0.89804 | TSPAN32 | 1.54E-60 | -0.67326 |
| IL1A | 7.32E-21 | -0.5414 | TUBA3C | 1.71E-06 | 0.531643 |
| IL1R2 | 2.10E-57 | -0.75661 | TXK | 1.55E-65 | -0.79447 |
| IL2 | 7.95E-45 | -1.758 | UBE2DNL | 0.027749274 | -1.09054 |
| IL21 | 7.08E-41 | -2.96491 | UBL4B | 2.54E-23 | -0.52011 |
| IL22RA2 | 2.85E-34 | -0.94225 | UGT1A10 | 0.009787583 | -0.8873 |
| IL23R | 3.92E-34 | -1.34947 | UGT1A7 | 0.017250618 | -0.84612 |
| IL25 | 0.00039 | 0.578475 | UGT1A9 | 0.003396223 | -0.54927 |
| IL26 | 2.35E-44 | -2.13987 | UGT2B7 | 5.31E-09 | -0.7141 |
| IL2RA | 1.69E-85 | -0.64483 | USP17L9P | 0.023399192 | 1.143543 |
| IL3 | 1.42E-07 | -1.64623 | UTS2 | 2.14E-36 | -1.18656 |
| IL31RA | 3.75E-24 | -0.83635 | UTS2B | 7.31E-27 | -0.66887 |
| IL36B | 0.003812 | -1.11822 | UTS2R | 8.75E-14 | -0.77081 |
| IL36G | 0.010124 | -0.54274 | VAX1 | 0.007372488 | -0.53945 |
| IL4 | 0.005852 | -0.78371 | VBCH | 4.83E-12 | -0.78689 |
| IL5RA | 1.10E-21 | -0.90395 | VGLL1 | 3.36E-11 | -0.56709 |
| IL6 | 1.92E-34 | -0.5516 | VNN2 | 1.00E-85 | -0.57211 |
| IL7R | 4.04E-64 | -0.55713 | VNN3 | 6.74E-20 | -0.79893 |
| IL9R | 4.37E-42 | -0.59173 | VPREB1 | 8.32E-12 | -2.80091 |
| ILTIF | 0.000252 | -1.42005 | VPREB3 | 2.81E-47 | -0.84928 |
| INFA4 | 0.017463 | -0.51225 | VRTN | 5.37E-06 | -0.78434 |
| INSC | 1.25E-09 | -0.57235 | VSTM1 | 0.000132507 | -0.52418 |
| INSL3 | 1.11E-54 | -0.95769 | VSTM2B | 0.008998193 | -0.57656 |
| IQCF6 | 0.0239 | 0.955573 | VSX2 | 2.45E-11 | -0.79675 |
| IRGM | 6.21E-28 | -1.08181 | VWC2L | 0.003424422 | 2.243326 |
| ISL2 | 1.18E-26 | -0.94065 | WDR64 | 1.66E-13 | -0.68951 |
| ITIH1 | 2.35E-39 | -1.2858 | WFDC10A | 0.011371824 | 0.867823 |
| ITK | 5.18E-92 | -0.59316 | WFDC12 | 0.00031256 | -0.77259 |
| ITLN1 | 1.11E-09 | -0.84509 | WFDC5 | 0.00260194 | -0.60371 |
| IVL | 1.92E-06 | -0.59916 | WNT1 | 3.89E-66 | -1.26735 |
| IZUMO1R | 8.04E-14 | -1.38787 | WNT10A | 5.47E-63 | -0.59819 |
| JAKMIP1 | 9.97E-41 | -0.56967 | WNT16 | 2.34E-20 | -0.73899 |
| JM27 | 0.030495 | 0.75954 | WNT6 | 5.60E-16 | -0.50332 |
| JSRP1 | 2.42E-28 | -0.78564 | WNT7A | 9.76E-22 | -1.21596 |
| KAL3 | 1.28E-13 | -1.21059 | XCL1 | 6.47E-67 | -0.82526 |
| KCNA10 | 0.00011 | -1.07459 | XCL2 | 1.90E-76 | -0.89996 |
| KCNA2 | 4.29E-34 | -1.0084 | XCR1 | 7.14E-49 | -1.11827 |
| KCNA3 | 5.72E-77 | -0.68369 | XIRP1 | 9.31E-45 | -0.66366 |
| KCNC2 | 0.00024 | 0.626017 | XKR4 | 2.69E-16 | -1.07535 |
| KCNE5 | 9.64E-19 | -0.52369 | XPNPEP2 | 1.18E-39 | -0.62469 |
| KCNG2 | 1.04E-14 | -0.51288 | ZAP70 | 2.08E-81 | -0.59564 |
| KCNG4 | 0.001322 | 1.259634 | ZBED2 | 1.97E-73 | -0.929 |
| KCNH4 | 1.71E-37 | -0.69048 | ZBP1 | 3.18E-64 | -0.57378 |
| KCNH6 | 0.00224 | 0.768364 | ZBTB32 | 7.80E-54 | -0.62503 |
| KCNJ10 | 2.40E-51 | -0.63734 | ZDHHC19 | 7.59E-19 | -0.93226 |
| KCNJ3 | 1.09E-12 | 0.781954 | ZDHHC22 | 4.07E-05 | 0.904577 |
| KCNK10 | 1.03E-22 | -0.72785 | ZNF683 | 1.76E-63 | -0.58775 |
| KCNQ1DN | 0.001052 | -0.72338 | ZNF80 | 1.97E-69 | -1.10898 |
| KHDRBS2 | 7.11E-14 | -0.88341 | ZNF804B | 0.004685097 | 0.803847 |
| KIAA0408 | 5.39E-08 | -0.56284 | ZNF831 | 1.39E-85 | -0.74838 |
| KIAA1045 | 5.11E-27 | -0.53006 | ZP2 | 0.001976931 | -0.74082 |
| KIR2DL1 | 8.03E-33 | -1.56472 | ZPBP2 | 1.36E-12 | -0.9404 |
| KIR2DL3 | 3.45E-31 | -1.3727 | LINC00028 | 1.43E-08 | -1.10896 |
| KIR2DL4 | 8.46E-48 | -1.33614 | LINC00032 | 0.000914624 | -0.90622 |
| KIR2DS4 | 1.75E-17 | -1.20961 | LINC00092 | 3.04E-31 | -0.81575 |
| KIR3DL2 | 7.38E-45 | -1.40803 | LINC00158 | 1.30E-42 | -1.43582 |
| KIR3DL3 | 9.47E-15 | -1.73856 | LINC00161 | 1.67E-08 | -0.7919 |
| KIR3DP1 | 0.015039 | -0.86334 | LINC00189 | 4.96E-25 | -0.94007 |
| KIR3DX1 | 1.19E-41 | -2.45922 | LINC00238 | 2.92E-05 | 0.821481 |
| KIRREL3 | 1.22E-22 | -0.58485 | LINC00239 | 6.29E-34 | -0.79365 |
| KLHL1 | 0.000642 | 1.482976 | LINC00426 | 1.12E-85 | -0.90756 |
| KLHL33 | 9.18E-10 | -0.59157 | LINC00483 | 0.017091333 | 0.530114 |
| KLHL34 | 1.16E-23 | -1.11253 | LINC00494 | 1.58E-19 | -0.56467 |
| KLK1 | 9.41E-20 | -0.51272 | LINC00521 | 0.009886639 | -0.83878 |
| KLK9 | 0.016971 | -0.84665 | LINC00599 | 0.002267507 | -0.52901 |
| KLRB1 | 9.90E-83 | -0.66738 | LINC00636 | 6.56E-05 | -0.60784 |
| KLRC1 | 1.40E-57 | -0.96545 | LINC00671 | 0.006869058 | -0.57924 |
| KLRC2 | 8.85E-31 | -0.69593 | LINC00698 | 0.00110319 | -0.74533 |
| KLRC3 | 5.94E-33 | -0.74263 | LINC00906 | 0.000677709 | -0.65011 |
| KLRC4 | 1.07E-24 | -1.11762 | LINC01142 | 2.05E-09 | -0.67992 |
| KLRD1 | 2.66E-78 | -0.61099 | LINC01366 | 6.90E-33 | -0.67427 |
| KLRF1 | 1.23E-52 | -0.8245 | LINC01550 | 2.34E-27 | -0.51493 |
| KLRK1 | 1.01E-81 | -0.5559 | LINC01551 | 0.029005698 | -0.75024 |
| KRT1 | 9.25E-08 | -0.52046 | LINC01559 | 0.000496982 | -0.76628 |
| KRT2 | 1.70E-15 | -0.8197 | LIPC | 6.07E-43 | -0.59734 |
| KRT31 | 3.89E-07 | 0.630591 | LIPL2 | 0.029790349 | -0.53575 |
| KRT33A | 0.005636 | 0.562652 | LIPL4 | 1.33E-11 | -1.5513 |
| KRT35 | 0.000123 | 1.288135 | LIVIN | 3.48E-16 | -0.62665 |
| KRT36 | 4.27E-15 | -0.65065 | LOC100124692 | 6.42E-05 | -0.51215 |
| KRT37 | 1.50E-09 | 0.615884 | LOC100129963 | 6.43E-28 | -0.65412 |
| KRT4 | 1.73E-08 | -0.60444 | LOC100130293 | 0.00727823 | -0.94992 |
| KRT72 | 2.19E-15 | -0.96944 | LOC100130829 | 0.012222656 | 0.684185 |
| KRT73 | 5.77E-08 | -0.82538 | LOC100131600 | 2.70E-90 | -0.76549 |
| KRT78 | 0.000205 | -0.64657 | LOC100133342 | 0.000216858 | -0.67991 |
| KRT84 | 0.0009 | -0.65545 | LOC100134052 | 2.44E-93 | -0.80018 |
| KRT85 | 2.03E-09 | -0.93086 | LOC100240735 | 1.77E-50 | -0.52 |
| KRT9 | 9.26E-07 | -0.57994 | LOC100293817 | 8.56E-10 | -0.57501 |
| KRTAP13-4 | 9.03E-06 | -2.24797 | LOC100508111 | 0.012263591 | -1.66304 |
| KRTAP19-1 | 0.041185 | -1.36332 | LOC100509003 | 1.89E-66 | -0.53333 |
| KRTAP2-2 | 0.048724 | -0.65216 | LOC101060231 | 1.30E-07 | -0.53314 |
| KRTAP3-3 | 0.014721 | 0.671002 | LOC101060582 | 1.63E-27 | -0.60975 |
| KRTAP6-2 | 0.027513 | #NAME? | LOC101928224 | 3.17E-40 | -0.56748 |
| KRTAP7-1 | 0.029158 | -0.68511 | LOC101929842 | 7.49E-09 | -0.6879 |
| LAIR2 | 1.90E-54 | -1.21228 | LOC101929877 | 2.31E-95 | -0.5212 |
| LANCL3 | 2.38E-24 | -0.53589 | LOC105371599 | 1.65E-49 | -0.63974 |
| LAX1 | 4.34E-76 | -0.66927 | LOC105374380 | 1.94E-20 | -0.82094 |
| LBP | 1.84E-17 | -0.59759 | LOC105378361 | 4.64E-05 | -0.87162 |
| LCE3A | 0.000807 | -1.28822 | LOC113104 | 6.28E-14 | -0.75509 |
| LCE3D | 0.027511 | -1.62608 | LOC115015 | 0.007306602 | -0.64428 |
| LCE3E | 0.002596 | -2.04419 | LOC115016 | 3.60E-05 | -0.93484 |
| LCE6A | 0.008987 | -2.21675 | LOC115252 | 0.000271399 | -2.32233 |
| LCN10 | 1.34E-26 | -0.72084 | LOC115271 | 3.15E-72 | -0.81497 |
| LCN6 | 4.13E-06 | -0.71623 | LOC115473 | 2.40E-17 | -0.52566 |
| LCN8 | 9.25E-06 | -1.2398 | LOC115545 | 3.02E-52 | -0.57375 |
| LCN9 | 0.038853 | -3.707 | LOC115632 | 0.007767201 | 0.97054 |
| LCNL1 | 3.92E-34 | -1.64984 | LOC115646 | 1.79E-13 | -0.69592 |
| LEMD1 | 8.38E-10 | -0.6399 | LOC115656 | 4.71E-35 | -1.4064 |
| LGALS17A | 3.72E-25 | -0.69607 | LOC115715 | 1.67E-11 | -0.54695 |
| LGALS2 | 2.97E-67 | -0.83525 | LOC115735 | 0.001597054 | 2.595886 |
| LILRA1 | 7.62E-36 | -0.50373 | LOC115806 | 9.23E-93 | -0.53287 |
| LILRA3 | 3.22E-41 | -0.98941 | LOC115842 | 0.007258586 | -0.55189 |
| LILRA4 | 5.24E-54 | -0.61317 | LOC115917 | 7.92E-31 | -0.59996 |
| LILRA5 | 2.02E-66 | -0.52766 | LOC115919 | 3.98E-23 | -1.38963 |
| LILRP2 | 5.99E-32 | -1.48672 | LOC115963 | 9.85E-13 | 0.666073 |
| LIM2 | 4.41E-08 | -1.90183 | LOC116109 | 9.76E-21 | -0.93366 |
| LIN28A | 0.015417 | 0.698405 | LOC116243 | 5.21E-53 | -0.93249 |
| LIN28B | 0.039221 | -0.68435 |  |  |  |

**Table S4. Differentially expressed genes between the metastasis and primary groups in GSE3521 cohort.**

| **Gene ID** | **Adj.p.value** | **logFC** | **Gene ID** | **Adj.p.value** | **logFC** |
| --- | --- | --- | --- | --- | --- |
| APOBEC3A | 0.038003 | 1.421899 | IQCE | 0.026645 | -0.80125 |
| ARHGAP1 | 0.030012 | -0.59358 | PF4V1 | 0.044066 | 0.783755 |
| COL12A1 | 0.015961 | -1.85279 | GDAP1 | 0.037102 | -0.84432 |
| CITED1 | 0.033266 | -1.57679 | FLJ38725 | 0.008722 | -0.97694 |
| MLF1IP | 0.011468 | 0.893082 | DET1 | 0.049208 | -0.52187 |
| HEXA | 0.026645 | -0.59346 | PQLC1 | 0.011468 | 0.590849 |
| GRP | 0.025598 | -1.45567 | C4orf18 | 0.033597 | -2.37639 |
| C11orf60 | 0.026645 | -0.81403 | HMMR | 0.0308 | 0.740164 |
| FCGR1A | 0.023351 | -0.77328 | CTNNAL1 | 0.006348 | 1.021828 |
| NFIL3 | 0.035057 | 0.783599 | YBX2 | 0.035057 | 0.63062 |
| CILP | 0.029241 | -1.35044 | HHAT | 0.023351 | -0.966 |
| COL5A1 | 0.026645 | -1.07221 | SFRS5 | 0.025011 | -0.61442 |
| COL1A2 | 0.011468 | -1.86863 | CACNA2D4 | 0.025011 | -0.73243 |
| RGS16 | 0.035626 | -0.90628 | LAMC3 | 0.026645 | 0.619769 |
| GJB2 | 0.03428 | -0.57864 | OLIG1 | 0.023351 | 1.741222 |
| DHODH | 0.026645 | 0.719874 | SMYD2 | 0.041152 | 0.63273 |
| GOLGB1 | 0.025598 | -0.51148 | CENPM | 0.026645 | 0.686855 |
| MXRA8 | 0.049208 | -0.85211 | BTG3 | 0.024493 | 0.977814 |
| MICAL-L2 | 0.011468 | -0.59192 | MUC16 | 0.038003 | 0.633681 |
| PLAU | 0.023351 | -1.42809 | NFIB | 0.04654 | 0.943058 |
| BIRC1 | 0.026645 | -1.04974 | CSPG2 | 0.029969 | -1.10558 |
| ZNF313 | 0.028307 | 0.589367 | ZNF596 | 0.023351 | -0.71806 |
| PTTG1 | 0.027437 | 0.758194 | C7 | 0.041986 | 0.638504 |
| IMPA2 | 0.023351 | 1.047762 | MATN3 | 0.026645 | -1.86496 |
| PEX11G | 0.025948 | -0.62048 | PHF11 | 0.030012 | -0.56187 |
| MFAP5 | 0.007536 | -1.65113 | LGALS3BP | 0.039916 | -0.78129 |
| POR | 0.034529 | 0.541255 | ZBTB5 | 0.026645 | 0.640025 |
| IER2 | 0.041986 | -0.61273 | ZNF223 | 0.034529 | -0.51943 |
| SULT1B1 | 0.048064 | 0.611996 | POSTN | 0.006348 | -1.85158 |
| LOXL1 | 0.008436 | -1.25922 | CEBPG | 0.023351 | 1.718588 |
| PIGH | 0.0429 | -0.61206 | UNC5B | 0.042541 | -1.01911 |
| GLI3 | 0.006348 | -1.70863 | SLC22A4 | 0.035626 | -0.65381 |
| RIT1 | 0.042541 | 0.622501 | PHF14 | 0.030012 | -0.53686 |
| COMP | 0.011468 | -1.74986 | SSPN | 0.025598 | -1.00601 |
| SLTM | 0.03428 | -0.55966 | GALNAC4S-6ST | 0.010296 | -1.1821 |
| YBX1 | 0.025011 | 0.602101 | CALML3 | 0.006348 | -3.25635 |
| ANGPTL4 | 0.0429 | 1.920953 | PTTG2 | 0.037102 | 0.640168 |
| DIP13B | 0.026645 | -0.5028 | SFRP2 | 0.000847 | -2.61553 |
| APH1B | 0.04654 | -0.67985 | FAM107A | 0.006348 | 1.510578 |
| BBS4 | 0.04654 | -0.57856 | SCUBE2 | 0.010296 | -3.14958 |
| BIRC5 | 0.049208 | 0.886126 | PDGFRL | 0.026645 | -1.13065 |
| PCBP4 | 0.03626 | 0.628944 | BHLHB3 | 0.011468 | -1.32123 |
| TMEM16A | 0.023351 | -1.05035 | PPAPDC3 | 0.041986 | -0.56648 |
| CTSK | 0.026645 | -1.6973 | MYBL1 | 0.026645 | 1.003718 |
| NT5DC1 | 0.043687 | -0.56211 | ZNF167 | 0.044066 | -0.63179 |
| POLR2A | 0.022184 | -0.60654 | BTBD3 | 0.04654 | 0.529766 |
| IFRD1 | 0.023351 | 1.41552 | COL1A1 | 0.023351 | -1.06809 |
| HTRA1 | 0.039162 | -1.44778 | CEP55 | 0.036458 | 0.758152 |
| TNFSF4 | 0.011609 | -1.21797 | OLFML3 | 0.029969 | -0.86832 |
| ITGA11 | 0.033266 | -1.5227 | GLT8D2 | 0.008345 | -0.8376 |
| CKS2 | 0.032636 | 0.903159 | SLC27A5 | 0.026645 | 0.877524 |
| CYB5D2 | 0.039916 | -1.00769 | CRY2 | 0.039916 | -0.55584 |
| HDGF | 0.026645 | 0.781055 | POLR1E | 0.040636 | 0.51789 |
| FLJ13639 | 0.022869 | -0.6776 | FAM62B | 0.041986 | 0.853118 |
| CCDC8 | 0.025011 | -0.97636 | C6orf128 | 0.008345 | -0.89462 |
| CNIH4 | 0.023351 | 0.680526 | SMOC2 | 0.025598 | -1.76144 |
| AEBP1 | 0.015594 | -1.03062 | BUB1 | 0.030012 | 0.929531 |
| TMOD1 | 0.045061 | 0.833221 | TAGLN2 | 0.023351 | 0.521254 |
| TYMS | 0.035626 | 0.802885 | EVC | 0.024224 | -0.71461 |
| ZBED5 | 0.04654 | -0.52116 | MARCO | 0.026645 | 1.083311 |
| ADRA2A | 0.023351 | -0.6751 | MGC24103 | 0.016717 | -1.56623 |
| PGC | 0.041669 | 0.545074 | NKX2-2 | 0.027067 | 0.770413 |
| SCRG1 | 0.008345 | 2.82025 | CENPE | 0.0308 | 0.674265 |
| CENPN | 0.035626 | 0.658702 | TRIP13 | 0.049208 | 0.682361 |
| WDR51A | 0.038003 | 0.771075 | ADAM12 | 0.039916 | -0.84588 |
| SERF1B | 0.011468 | 0.526063 | MMP13 | 0.049208 | -1.58555 |
| ANLN | 0.041761 | 0.973208 | TCF21 | 0.04654 | 0.764037 |
| C1orf37 | 0.028757 | 0.526628 | C20orf103 | 0.023351 | -1.06817 |
| MMP3 | 0.018634 | -1.08739 | FLJ90709 | 0.008361 | -0.60717 |
| INHBA | 0.023351 | -1.12671 | SULF2 | 0.008722 | -1.48854 |
| RICS | 0.0308 | -0.79295 | PER2 | 0.021408 | -0.98047 |
| FAP | 0.023351 | -1.54489 | DEPDC1 | 0.018124 | 0.854664 |
| SFTPA1 | 0.026645 | 1.08707 | NUDT10 | 0.023351 | -0.75517 |
| PDCD5 | 0.026645 | 1.569097 | ZNF45 | 0.026645 | -0.59635 |
| EGLN1 | 0.023351 | 0.763139 | MAP2 | 0.016717 | 1.080392 |
| CAMTA1 | 0.027252 | 0.53559 | CDKN2D | 0.025598 | 0.913789 |
| PSD4 | 0.016717 | -0.50845 | UCK2 | 0.034529 | 0.756652 |
| TMEM26 | 0.025927 | -0.6776 | MBD6 | 0.030012 | -0.68627 |
| IBTK | 0.026645 | -0.58887 | EGR3 | 0.037099 | -1.11782 |
| PAICS | 0.04654 | 0.533398 | TTC12 | 0.041986 | -0.67454 |
| UBE2T | 0.012913 | 1.047252 | CERK | 0.023351 | 0.767521 |
| ZBED1 | 0.034529 | -0.6505 | C20orf20 | 0.049208 | 0.58751 |
| C16orf61 | 0.011468 | 0.64605 | APOBEC3B | 0.045061 | 1.663813 |
| RP11-50D16.3 | 0.049208 | -0.72311 | ARG1 | 0.006348 | 3.928526 |
| SVH | 0.04654 | 0.589637 | DRD5 | 0.04654 | 0.514115 |
| RPL30 | 0.044657 | 0.543946 | SLC7A8 | 0.041761 | -1.8217 |
| PGR | 0.041761 | -1.91892 |  |  |  |

**Table S5 Differentially expressed genes between the metastasis and primary groups in GSE10893 cohort.**

| Gene ID | P.value | Adj.p.value | Gene ID | P.value | Adj.p.value |
| --- | --- | --- | --- | --- | --- |
| DKFZp434K1815 | 0.001293 | 0.022448 | STK11 | 0.001293 | 0.022448 |
| C9orf93 | 0.000754 | 0.017438 | DDX52 | 0.001293 | 0.022448 |
| GSTM4 | 0.000108 | 0.007806 | LANCL1 | 0.001293 | 0.022448 |
| MBD3 | 0.000431 | 0.012946 | CNTNAP3 | 0.000431 | 0.012946 |
| RFPL3S | 0.003232 | 0.034676 | IL12RB1 | 0.00474 | 0.042105 |
| UHMK1 | 0.000754 | 0.017438 | CENTB2 | 0.000754 | 0.017438 |
| SRRM1 | 0.000108 | 0.007806 | LCMT2 | 0.003232 | 0.034676 |
| KIAA1856 | 0.00474 | 0.042105 | WDFY2 | 0.001293 | 0.022448 |
| FRMD4A | 0.002047 | 0.027424 | TOMM40 | 0.003232 | 0.034676 |
| JUN | 0.00474 | 0.042105 | C14orf108 | 0.000431 | 0.012946 |
| IGFBP4 | 0.00474 | 0.042105 | ITIH4 | 0.002047 | 0.027424 |
| CCDC116 | 0.000108 | 0.007806 | RAD54L | 0.000754 | 0.017438 |
| IGSF1 | 0.000108 | 0.007806 | DPH2 | 0.003232 | 0.034676 |
| C6orf111 | 0.003232 | 0.034676 | PHTF2 | 0.002047 | 0.027424 |
| ZNRF2 | 0.000431 | 0.012946 | UCN2 | 0.002047 | 0.027424 |
| SETD2 | 0.000108 | 0.007806 | SCLT1 | 0.000108 | 0.007806 |
| C13orf3 | 0.003232 | 0.034676 | P2RY1 | 0.002047 | 0.027424 |
| HSP90AB1 | 0.000431 | 0.012946 | LOC652408 | 0.002047 | 0.027424 |
| PLEKHF1 | 0.00474 | 0.042105 | PHF21A | 0.000754 | 0.017438 |
| LOC642678 | 0.002047 | 0.027424 | C22orf15 | 0.003232 | 0.034676 |
| IRS2 | 0.003232 | 0.034676 | RYBP | 0.00474 | 0.042105 |
| VPS26A | 0.003232 | 0.034676 | ZNF467 | 0.00474 | 0.042105 |
| LOC645671 | 0.003232 | 0.034676 | UBQLN2 | 0.000754 | 0.017438 |
| RASA1 | 0.001293 | 0.022448 | CYP4Z1 | 0.003232 | 0.034676 |
| PRR13 | 0.002047 | 0.027424 | MTF2 | 0.000215 | 0.010104 |
| PTD008 | 0.000215 | 0.010104 | CACNG6 | 0.00474 | 0.042105 |
| PCDHGA2 | 0.000108 | 0.007806 | KIAA0649 | 0.001293 | 0.022448 |
| EIF4G1 | 0.003232 | 0.034676 | PAX6 | 0.000108 | 0.007806 |
| MAGEA5 | 0.000431 | 0.012946 | NACAP1 | 0.002047 | 0.027424 |
| HSPB3 | 0.000431 | 0.012946 | MYH14 | 0.000108 | 0.007806 |
| PMS2L3 | 0.002047 | 0.027424 | TYR | 0.000754 | 0.017438 |
| PSMD5 | 0.003232 | 0.034676 | USP24 | 0.002323 | 0.0309 |
| ATP7B | 0.003232 | 0.034676 | ORC2L | 0.000431 | 0.012946 |
| CCHCR1 | 0.00474 | 0.042105 | C14orf106 | 0.003232 | 0.034676 |
| ACOT4 | 0.000754 | 0.017438 | RPL10A | 0.001293 | 0.022448 |
| TBL3 | 0.000108 | 0.007806 | SMAD1 | 0.00474 | 0.042105 |
| C21orf108 | 0.00474 | 0.042105 | ZNF283 | 0.003232 | 0.034676 |
| OR2A7 | 0.003232 | 0.034676 | POLD3 | 0.000108 | 0.007806 |
| ETV5 | 0.000431 | 0.012946 | C6orf108 | 0.00474 | 0.042105 |
| STX2 | 0.002047 | 0.027424 | NUTF2 | 0.00474 | 0.042105 |
| RBMS2 | 0.00474 | 0.042105 | ZNF576 | 0.000108 | 0.007806 |
| EPS15L1 | 0.001293 | 0.022448 | WTAP | 0.000431 | 0.012946 |
| SHMT2 | 0.000431 | 0.012946 | CSNK1A1L | 0.000108 | 0.007806 |
| FAM102B | 0.001225 | 0.022448 | FZD5 | 0.00474 | 0.042105 |
| RASL11A | 0.00474 | 0.042105 | KIAA1644 | 0.000215 | 0.010104 |
| MDM4 | 0.003232 | 0.034676 | HCG4 | 0.00474 | 0.042105 |
| ALDH8A1 | 0.002047 | 0.027424 | FLJ21908 | 0.001293 | 0.022448 |
| BIRC7 | 0.000754 | 0.017438 | GABRG2 | 0.001293 | 0.022448 |
| Tenr | 0.003232 | 0.034676 | TNXB | 0.003232 | 0.034676 |
| SPCS1 | 0.002047 | 0.027424 | KIAA1344 | 0.000431 | 0.012946 |
| PSORS1C2 | 0.001293 | 0.022448 | CAPZA2 | 0.001293 | 0.022448 |
| DGKG | 0.000108 | 0.007806 | C6orf173 | 0.000215 | 0.010104 |
| PCSK9 | 0.001694 | 0.027424 | GPR68 | 0.002047 | 0.027424 |
| BTN2A3 | 0.000108 | 0.007806 | TCF12 | 0.000431 | 0.012946 |
| NR1I2 | 0.000108 | 0.007806 | FOXD4L4 | 0.002047 | 0.027424 |
| IAPP | 0.00474 | 0.042105 | MGC11332 | 0.001293 | 0.022448 |
| NUP205 | 0.002047 | 0.027424 | ENO1 | 0.000215 | 0.010104 |
| RAD54L2 | 0.000431 | 0.012946 | ZNF20 | 0.000108 | 0.007806 |
| ZNF35 | 0.003232 | 0.034676 | HCG18 | 0.000431 | 0.012946 |
| ZNF263 | 0.003232 | 0.034676 | ALKBH1 | 0.000215 | 0.010104 |
| C14orf45 | 0.001293 | 0.022448 | PAICS | 0.000754 | 0.017438 |
| SNF1LK | 0.002047 | 0.027424 | RALBP1 | 0.000108 | 0.007806 |
| LPIN1 | 0.003232 | 0.034676 | CYB5R1 | 0.003232 | 0.034676 |
| TCEAL4 | 0.003232 | 0.034676 | WWTR1 | 0.000108 | 0.007806 |
| AHSG | 0.003232 | 0.034676 | TCF1 | 0.000108 | 0.007806 |
| ESR1 | 0.000431 | 0.012946 | TUBB | 0.003232 | 0.034676 |
| GPR162 | 0.000431 | 0.012946 | EEF1A1 | 0.000754 | 0.017438 |
| SPTLC2 | 0.000431 | 0.012946 | CRISPLD2 | 0.000108 | 0.007806 |
| C5orf21 | 0.000754 | 0.017438 | DST | 0.000108 | 0.007806 |
| PDK1 | 0.000754 | 0.017438 | LOC389332 | 0.002047 | 0.027424 |
| KCTD18 | 0.000108 | 0.007806 | TMEM162 | 0.002047 | 0.027424 |
| ARMCX5 | 0.000431 | 0.012946 | HIT-40 | 0.001293 | 0.022448 |
| RANBP2 | 0.00474 | 0.042105 | HAGHL | 0.003232 | 0.034676 |
| DUSP15 | 0.004936 | 0.043629 | POPDC3 | 0.000431 | 0.012946 |
| HRH2 | 0.003232 | 0.034676 | HIGD1B | 0.00474 | 0.042105 |
| PGAM5 | 0.000431 | 0.012946 | RNF170 | 0.002047 | 0.027424 |
| SH3BP4 | 0.00474 | 0.042105 | SNRPD2 | 0.000108 | 0.007806 |
| AMOTL1 | 0.000108 | 0.007806 | GCNT2 | 0.00474 | 0.042105 |
| CMIP | 0.002047 | 0.027424 | PPM1A | 0.003232 | 0.034676 |
| PRKRIP1 | 0.00474 | 0.042105 | TOMM70A | 0.000431 | 0.012946 |
| UBB | 0.001293 | 0.022448 | DNTT | 0.000108 | 0.007806 |
| NAV1 | 0.000108 | 0.007806 | MAP1LC3B | 0.002047 | 0.027424 |
| PPP2R2A | 0.001293 | 0.022448 | STXBP5 | 0.00474 | 0.042105 |
| PTPN14 | 0.000215 | 0.010104 | FAM44A | 0.000108 | 0.007806 |
| CDC42BPA | 0.000754 | 0.017438 | TTR | 0.000108 | 0.007806 |
| FBXW4 | 0.002047 | 0.027424 | LOC283412 | 0.002047 | 0.027424 |
| SLC17A5 | 0.003232 | 0.034676 | SEC14L5 | 0.001293 | 0.022448 |
| TJAP1 | 0.00474 | 0.042105 | ARCN1 | 0.001293 | 0.022448 |
| SH2B2 | 0.003232 | 0.034676 | DEDD2 | 0.00474 | 0.042105 |
| CCDC57 | 0.00474 | 0.042105 | MT | 0.003674 | 0.039253 |
| GCS1 | 0.000754 | 0.017438 | GNL2 | 0.001293 | 0.022448 |
| WDR4 | 0.000215 | 0.010104 | LIN28 | 0.00474 | 0.042105 |
| SLC5A3 | 0.003232 | 0.034676 | LOC339745 | 0.000108 | 0.007806 |
| XRN1 | 0.000108 | 0.007806 | NAB1 | 0.000108 | 0.007806 |
| KIAA1546 | 0.000431 | 0.012946 | PARD6G | 0.00474 | 0.042105 |
| FAM120A | 0.00474 | 0.042105 | GABPB2 | 0.000108 | 0.007806 |
| LOC647971 | 0.000108 | 0.007806 | GH1 | 0.002047 | 0.027424 |
| MGC42367 | 0.000108 | 0.007806 | PHF8 | 0.000108 | 0.007806 |
| LSM14B | 0.000431 | 0.012946 | LHX1 | 0.001293 | 0.022448 |
| PTMA | 0.003232 | 0.034676 | HAMP | 0.000108 | 0.007806 |
| C19orf26 | 0.000215 | 0.010104 | LOC643328 | 0.002047 | 0.027424 |
| GART | 0.000215 | 0.010104 | LRPPRC | 0.000215 | 0.010104 |
| GLYATL1 | 0.000754 | 0.017438 | DPPA4 | 0.000108 | 0.007806 |
| GDF3 | 0.000431 | 0.012946 | C19orf24 | 0.000754 | 0.017438 |
| TCEA2 | 0.002323 | 0.0309 | KIAA0372 | 0.000879 | 0.019929 |
| PTTG1IP | 0.000108 | 0.007806 | NAG6 | 0.001293 | 0.022448 |
| C9orf82 | 0.000108 | 0.007806 | NPHP4 | 0.002047 | 0.027424 |
| ZIC3 | 0.000754 | 0.017438 | C11orf56 | 0.000431 | 0.012946 |
| LOC646643 | 0.003232 | 0.034676 | PROKR1 | 0.000754 | 0.017438 |
| IFRD1 | 0.000215 | 0.010104 | RAD23B | 0.00474 | 0.042105 |
| LOC653778 | 0.000754 | 0.017438 | LOC653820 | 0.000108 | 0.007806 |
| LOC644739 | 0.002047 | 0.027424 | TCF25 | 0.000754 | 0.017438 |
| SBSN | 0.000215 | 0.010104 | MPHOSPH9 | 0.003232 | 0.034676 |
| C17orf66 | 0.002047 | 0.027424 | EWSR1 | 0.000431 | 0.012946 |
| MLANA | 0.000108 | 0.007806 | IRX4 | 0.003232 | 0.034676 |
| TMEM158 | 0.00474 | 0.042105 | EFCAB2 | 0.000431 | 0.012946 |
| TSPY1 | 0.000754 | 0.017438 | MAGEA6 | 0.002712 | 0.034676 |
| CSS3 | 0.000754 | 0.017438 | C10orf32 | 0.003232 | 0.034676 |
| SLC39A10 | 0.002047 | 0.027424 | RPL29 | 0.00474 | 0.042105 |
| SMCHD1 | 0.002047 | 0.027424 | LOC650084 | 0.00474 | 0.042105 |
| VASP | 0.000108 | 0.007806 | CDC2L6 | 0.000754 | 0.017438 |
| PRKCSH | 0.00474 | 0.042105 | ATP10D | 0.00474 | 0.042105 |
| TMEM23 | 0.003232 | 0.034676 | NACAL | 0.001293 | 0.022448 |
| LIPC | 0.001293 | 0.022448 | CEP78 | 0.003232 | 0.034676 |
| KLF3 | 0.001293 | 0.022448 | PTK6 | 0.001293 | 0.022448 |
| TRIB2 | 0.000215 | 0.010104 | ZNF442 | 0.001293 | 0.022448 |
| ITGB1BP3 | 0.00474 | 0.042105 | CRYGS | 0.000108 | 0.007806 |
| HIST1H1D | 0.000108 | 0.007806 | FAM116B | 0.000431 | 0.012946 |
| PALM | 0.003232 | 0.034676 | ZNFN1A5 | 0.00474 | 0.042105 |
| TEKT4 | 0.000754 | 0.017438 | PKN2 | 0.00474 | 0.042105 |
| SKI | 0.003232 | 0.034676 | C4BPB | 0.002047 | 0.027424 |
| RBM23 | 0.000108 | 0.007806 | CHRNB2 | 0.001293 | 0.022448 |
| ANKRD36 | 0.003232 | 0.034676 | SERBP1 | 0.000431 | 0.012946 |
| BAGE5 | 0.000754 | 0.017438 | ODF3 | 0.001293 | 0.022448 |
| FNBP1L | 0.00474 | 0.042105 | CLDN6 | 0.000215 | 0.010104 |
| TP53I13 | 0.000754 | 0.017438 | APOM | 0.000431 | 0.012946 |
| NAT1 | 0.002047 | 0.027424 | KCNJ9 | 0.000431 | 0.012946 |
| C6orf134 | 0.000108 | 0.007806 | WDFY1 | 0.002047 | 0.027424 |
| UBE2D2 | 0.001293 | 0.022448 | ZDHHC3 | 0.000215 | 0.010104 |
| CLEC2A | 0.000108 | 0.007806 | LOC493754 | 0.000108 | 0.007806 |
| GFRA4 | 0.00474 | 0.042105 | FABP1 | 0.000215 | 0.010104 |
| LOC146429 | 0.000108 | 0.007806 | BCL2L11 | 0.001293 | 0.022448 |
| DLX4 | 0.000215 | 0.010104 | CYP1A2 | 0.003232 | 0.034676 |
| SLC2A13 | 0.000215 | 0.010104 | HIST1H1B | 0.00474 | 0.042105 |
| LRRFIP1 | 0.002047 | 0.027424 | RPS2 | 0.000215 | 0.010104 |
| OTUD6A | 0.00474 | 0.042105 | ZNF564 | 0.002047 | 0.027424 |
| PHLDA2 | 0.003232 | 0.034676 | C14orf28 | 0.001293 | 0.022448 |
| CPB2 | 0.000431 | 0.012946 | CPS1 | 0.001293 | 0.022448 |
| THOC2 | 0.000108 | 0.007806 | KIAA1279 | 0.001293 | 0.022448 |
| OTUD4 | 0.002047 | 0.027424 | ZCSL3 | 0.002047 | 0.027424 |
| PDE6A | 0.001293 | 0.022448 | CPT1A | 0.000108 | 0.007806 |
| DHX32 | 0.002047 | 0.027424 | RUFY3 | 0.001293 | 0.022448 |
| TLX2 | 0.001293 | 0.022448 | TEP1 | 0.000431 | 0.012946 |
| ASH1L | 0.00474 | 0.042105 | FOXQ1 | 0.00474 | 0.042105 |
| TCEAL3 | 0.00474 | 0.042105 | OXT | 0.002047 | 0.027424 |
| EIF2C2 | 0.001293 | 0.022448 | FAM9B | 0.000754 | 0.017438 |
| PDGFD | 0.000431 | 0.012946 | SNX26 | 0.003232 | 0.034676 |
| NANOG | 0.00474 | 0.042105 | DMC1 | 0.000431 | 0.012946 |
| TSPAN10 | 0.002047 | 0.027424 | SRP68 | 0.000754 | 0.017438 |
| SFRS2 | 0.003232 | 0.034676 | CDV3 | 0.000879 | 0.019929 |
| EIF3S5 | 0.000754 | 0.017438 | SSH1 | 0.000108 | 0.007806 |
| PLD1 | 0.002047 | 0.027424 | SLC25A13 | 0.000754 | 0.017438 |
| ABCB6 | 0.003232 | 0.034676 | CYP4Z2P | 0.003232 | 0.034676 |
| HSPB9 | 0.00474 | 0.042105 | RFWD2 | 0.002047 | 0.027424 |
| HPCA | 0.003232 | 0.034676 | DMP1 | 0.00316 | 0.034676 |
| LOC653504 | 0.000215 | 0.010104 | IGFBPL1 | 0.001293 | 0.022448 |
| KIAA1586 | 0.000431 | 0.012946 | SOX8 | 0.001293 | 0.022448 |
| MMS19L | 0.00316 | 0.034676 | DBNDD2 | 0.000108 | 0.007806 |
| RUVBL1 | 0.000431 | 0.012946 | TYW1 | 0.000215 | 0.010104 |
| FGFR3 | 0.000108 | 0.007806 | WNT6 | 0.001293 | 0.022448 |
| GALNT6 | 0.00474 | 0.042105 | SPAG4L | 0.001293 | 0.022448 |
| HSP90B1 | 0.003232 | 0.034676 | LOC285908 | 0.002323 | 0.0309 |
| ADNP | 0.000754 | 0.017438 | TCF19 | 0.000215 | 0.010104 |
| C16orf33 | 0.00474 | 0.042105 | NUDT15 | 0.000108 | 0.007806 |
| IL1F7 | 0.00474 | 0.042105 | APOA5 | 0.001293 | 0.022448 |
| YEATS4 | 0.002047 | 0.027424 | GPR172A | 0.00474 | 0.042105 |
| CG018 | 0.000754 | 0.017438 | MTAC2D1 | 0.002047 | 0.027424 |
| CHMP5 | 0.002047 | 0.027424 | CYBRD1 | 0.001293 | 0.022448 |
| FLJ20628 | 0.000754 | 0.017438 | DMAP1 | 0.002047 | 0.027424 |
| ACTRT2 | 0.001293 | 0.022448 | LOC650938 | 0.000215 | 0.010104 |
| MIA3 | 0.003232 | 0.034676 | STS-1 | 0.001293 | 0.022448 |
| HTR1A | 0.00474 | 0.042105 | LOC653622 | 0.000108 | 0.007806 |
| TTLL4 | 0.000108 | 0.007806 | LOC91431 | 0.000431 | 0.012946 |
| L3MBTL2 | 0.001986 | 0.027424 | MGC9850 | 0.000215 | 0.010104 |
| SLA | 0.000754 | 0.017438 | FLJ10159 | 0.003232 | 0.034676 |
| EMX2OS | 0.00474 | 0.042105 | KIAA0020 | 0.001293 | 0.022448 |
| CABIN1 | 0.000108 | 0.007806 | METT10D | 0.000215 | 0.010104 |
| KIAA1958 | 0.00474 | 0.042105 | NTNG1 | 0.000108 | 0.007806 |
| ENTPD5 | 0.000879 | 0.019929 | VCX-C | 0.000215 | 0.010104 |
| KIAA0701 | 0.00474 | 0.042105 | EBNA1BP2 | 0.000108 | 0.007806 |
| GLIPR1L2 | 0.001685 | 0.027424 | C21orf58 | 0.001293 | 0.022448 |
| HNF4A | 0.000754 | 0.017438 | LOC653147 | 0.003232 | 0.034676 |
| TP53I11 | 0.003232 | 0.034676 | SUGT1P | 0.00474 | 0.042105 |
| SLFN13 | 0.002047 | 0.027424 | AMHR2 | 0.000431 | 0.012946 |
| BTG2 | 0.000754 | 0.017438 | CDH10 | 0.000108 | 0.007806 |
| ACSBG1 | 0.000215 | 0.010104 | ANKRD11 | 0.000108 | 0.007806 |
| NFYB | 0.002698 | 0.034676 | GNA14 | 0.000215 | 0.010104 |
| TPP2 | 0.003232 | 0.034676 | TAF4B | 0.000215 | 0.010104 |
| SIRT1 | 0.000431 | 0.012946 | PMS2 | 0.004936 | 0.043629 |
| CACNG4 | 0.00474 | 0.042105 | CYP2A7 | 0.000431 | 0.012946 |
| BYSL | 0.000431 | 0.012946 | POP1 | 0.00474 | 0.042105 |
| C9orf140 | 0.000431 | 0.012946 | PLCB1 | 0.00474 | 0.042105 |
| ASB2 | 0.001293 | 0.022448 | RHAG | 0.000108 | 0.007806 |
| LOC644165 | 0.00474 | 0.042105 | OPN1LW | 0.00474 | 0.042105 |
| GPR150 | 0.00474 | 0.042105 | SOLH | 0.001293 | 0.022448 |
| CREB1 | 0.002047 | 0.027424 | EFHA1 | 0.003232 | 0.034676 |
| EXOC6 | 0.002047 | 0.027424 | PLA2G2D | 0.000754 | 0.017438 |
| DHRS7 | 0.000108 | 0.007806 | GBX2 | 0.003232 | 0.034676 |
| FAM58A | 0.003232 | 0.034676 | PRPF38A | 0.003232 | 0.034676 |
| KRT3 | 0.00474 | 0.042105 | PRKAB1 | 0.002047 | 0.027424 |
| AVP | 0.003232 | 0.034676 | JMJD2A | 0.000431 | 0.012946 |
| DERL1 | 0.000431 | 0.012946 | LPIN2 | 0.000215 | 0.010104 |
| FOXD2 | 0.00474 | 0.042105 | SLC6A17 | 0.004936 | 0.043629 |
| LOC654155 | 0.003232 | 0.034676 | ZNF354A | 0.003232 | 0.034676 |
| NRSN2 | 0.000754 | 0.017438 | C1orf52 | 0.000108 | 0.007806 |
| TFEB | 0.000215 | 0.010104 | AJAP1 | 0.00474 | 0.042105 |
| JARID1A | 0.001293 | 0.022448 | LOC388474 | 0.00474 | 0.042105 |
| LRRIQ2 | 0.002047 | 0.027424 | DAZAP2 | 0.003232 | 0.034676 |
| CAMK2N2 | 0.001293 | 0.022448 | U2AF1L1 | 0.000108 | 0.007806 |
| DMXL1 | 0.003232 | 0.034676 | CLSTN2 | 0.00474 | 0.042105 |
| ELA2A | 0.000108 | 0.007806 | HEY2 | 0.00474 | 0.042105 |
| B3GALNT2 | 0.001293 | 0.022448 | NCLN | 0.003232 | 0.034676 |
| RAB33B | 0.002047 | 0.027424 | SLC12A6 | 0.000215 | 0.010104 |
| GCK | 0.000108 | 0.007806 | ADAM11 | 0.003232 | 0.034676 |
| C1orf59 | 0.003232 | 0.034676 | C1orf144 | 0.001293 | 0.022448 |
| ABP1 | 0.000108 | 0.007806 | HS3ST4 | 0.002047 | 0.027424 |
| MICAL1 | 0.002047 | 0.027424 | C20orf117 | 0.000879 | 0.019929 |
| RAG2 | 0.003232 | 0.034676 | MGC29671 | 0.00474 | 0.042105 |
| CAMK2D | 0.000431 | 0.012946 | REST | 0.003232 | 0.034676 |
| DSCR8 | 0.00474 | 0.042105 | JMJD1C | 0.000108 | 0.007806 |
| SRI | 0.002047 | 0.027424 | CPA3 | 0.000108 | 0.007806 |
| TMEM131 | 0.000431 | 0.012946 | ZNF179 | 0.001293 | 0.022448 |
| DDX17 | 0.002047 | 0.027424 | PI4KII | 0.000108 | 0.007806 |
| OPA3 | 0.00474 | 0.042105 | PDE6B | 0.000431 | 0.012946 |
| LOC348180 | 0.001293 | 0.022448 | RBM15 | 0.000754 | 0.017438 |
| LOC652010 | 0.001293 | 0.022448 | CCL25 | 0.000754 | 0.017438 |
| C15orf27 | 0.000431 | 0.012946 | PORCN | 0.000754 | 0.017438 |
| ZNF423 | 0.003232 | 0.034676 | LOC643416 | 0.003232 | 0.034676 |
| SLC26A2 | 0.000108 | 0.007806 | RAI1 | 0.000431 | 0.012946 |
| C1orf187 | 0.000754 | 0.017438 | LOC653596 | 0.003232 | 0.034676 |
| UTS2R | 0.001293 | 0.022448 | UBE2S | 0.000754 | 0.017438 |
| MLX | 0.000431 | 0.012946 | MRPL52 | 0.000431 | 0.012946 |
| C1orf175 | 0.00474 | 0.042105 | RHBDD2 | 0.000108 | 0.007806 |
| ZNF205 | 0.003232 | 0.034676 | CSDA | 0.003232 | 0.034676 |
| EEF1A1 | 0.000431 | 0.012946 | CNTN4 | 0.000108 | 0.007806 |
| KIAA1679 | 0.002047 | 0.027424 | HLF | 0.002047 | 0.027424 |
| GRAP2 | 0.000215 | 0.010104 | ATF4 | 0.003232 | 0.034676 |
| MTTP | 0.00474 | 0.042105 | SMARCA2 | 0.000108 | 0.007806 |
| CRIP1 | 0.000215 | 0.010104 | C9orf52 | 0.003232 | 0.034676 |
| CCNG1 | 0.001293 | 0.022448 | KITLG | 0.003232 | 0.034676 |
| GPR120 | 0.001293 | 0.022448 | RP3-452H17.2 | 0.003232 | 0.034676 |
| TTC9 | 0.001293 | 0.022448 | SUNC1 | 0.000431 | 0.012946 |
| DDX19A | 0.001293 | 0.022448 | SLC25A3 | 0.003232 | 0.034676 |
| C13orf8 | 0.001293 | 0.022448 | MTRF1 | 0.00474 | 0.042105 |
| CHGB | 0.000754 | 0.017438 | FNBP4 | 0.000108 | 0.007806 |
| RPS15A | 0.001293 | 0.022448 | FOXK1 | 0.002047 | 0.027424 |
| RPL10 | 0.00474 | 0.042105 | MYCN | 0.000431 | 0.012946 |
| ATRX | 0.000431 | 0.012946 | CXorf6 | 0.00474 | 0.042105 |
| ZNF650 | 0.001293 | 0.022448 | KIF9 | 0.00474 | 0.042105 |
| RPL11 | 0.002047 | 0.027424 | CASC4 | 0.003232 | 0.034676 |
| CENPJ | 0.000431 | 0.012946 | ADAMTSL1 | 0.000431 | 0.012946 |
| SEC31L2 | 0.002047 | 0.027424 | NPBWR1 | 0.001293 | 0.022448 |
| ZBTB43 | 0.001293 | 0.022448 | ZNF436 | 0.00474 | 0.042105 |
| LOC374920 | 0.000754 | 0.017438 | APEH | 0.001293 | 0.022448 |
| NOX4 | 0.00474 | 0.042105 | FLJ45717 | 0.002047 | 0.027424 |
| C12orf28 | 0.003232 | 0.034676 | KRT85 | 0.000754 | 0.017438 |
| SFPQ | 0.002712 | 0.034676 | RBM39 | 0.002047 | 0.027424 |
| C3orf23 | 0.002047 | 0.027424 | PKN1 | 0.000431 | 0.012946 |
| SMAP1 | 0.002047 | 0.027424 | MGC33584 | 0.00474 | 0.042105 |
| GLI4 | 0.000754 | 0.017438 | STK4 | 0.003232 | 0.034676 |
| OSMR | 0.000215 | 0.010104 | CRYGA | 0.000108 | 0.007806 |
| RPL31 | 0.003232 | 0.034676 | B4GALT2 | 0.000431 | 0.012946 |
| ANKS1B | 0.000108 | 0.007806 | ATP5B | 0.002047 | 0.027424 |
| U2AF1 | 0.000108 | 0.007806 | Septin 7 | 0.000431 | 0.012946 |
| UCP2 | 0.00474 | 0.042105 | PRAC | 0.000431 | 0.012946 |
| SERINC1 | 0.00474 | 0.042105 | LRFN1 | 0.002047 | 0.027424 |
| MYOD1 | 0.001293 | 0.022448 | ZDHHC21 | 0.000108 | 0.007806 |
| YTHDC2 | 0.002047 | 0.027424 | IRAK1 | 0.003232 | 0.034676 |
| TMPRSS5 | 0.00474 | 0.042105 | FLCN | 0.000431 | 0.012946 |
| SR140 | 0.000108 | 0.007806 | NUDCD3 | 0.000754 | 0.017438 |
| LOC440264 | 0.002047 | 0.027424 | FLJ43093 | 0.00474 | 0.042105 |
| LOC645638 | 0.003232 | 0.034676 | FOXC2 | 0.00474 | 0.042105 |
| MGC40574 | 0.00474 | 0.042105 | LCT | 0.000108 | 0.007806 |
| MDH2 | 0.002047 | 0.027424 | FANCA | 0.003232 | 0.034676 |
| MED19 | 0.00474 | 0.042105 | PAGE2 | 0.001293 | 0.022448 |
| CEACAM6 | 0.002047 | 0.027424 | IPO8 | 0.003232 | 0.034676 |
| DPF3 | 0.000431 | 0.012946 | HMG4L | 0.002047 | 0.027424 |
| C4A | 0.00474 | 0.042105 | FZD8 | 0.001293 | 0.022448 |
| BRRN1 | 0.001293 | 0.022448 | TTC3 | 0.004936 | 0.043629 |
| UGCGL1 | 0.00474 | 0.042105 | LOC652671 | 0.00474 | 0.042105 |
| RP11-130N24.1 | 0.000108 | 0.007806 | TRIM65 | 0.000108 | 0.007806 |
| LOC346887 | 0.000754 | 0.017438 | CRELD2 | 0.001293 | 0.022448 |
| C11orf35 | 0.00474 | 0.042105 | MRPL49 | 0.002047 | 0.027424 |
| MGAT5B | 0.000754 | 0.017438 | ZNF512 | 0.000754 | 0.017438 |
| TMC5 | 0.003232 | 0.034676 | PANX2 | 0.001293 | 0.022448 |
| NRIP2 | 0.003232 | 0.034676 | FOXA2 | 0.003232 | 0.034676 |
| DDX42 | 0.000108 | 0.007806 | ZNF559 | 0.002047 | 0.027424 |
| MAGEA4 | 0.000108 | 0.007806 | CTPS | 0.001442 | 0.024998 |
| CCDC76 | 0.000215 | 0.010104 | OSBP2 | 0.001293 | 0.022448 |
| MIER1 | 0.000108 | 0.007806 | OXR1 | 0.003232 | 0.034676 |
| FOXD1 | 0.002047 | 0.027424 | MGC71993 | 0.000754 | 0.017438 |
| KATNAL1 | 0.000879 | 0.019929 | STEAP3 | 0.002047 | 0.027424 |
| CKAP4 | 0.002047 | 0.027424 | SLC35E4 | 0.003232 | 0.034676 |
| TXNDC4 | 0.00474 | 0.042105 | EPHA2 | 0.002047 | 0.027424 |
| DNAJC3 | 0.001293 | 0.022448 | RPS11 | 0.002047 | 0.027424 |
| TREM2 | 0.001293 | 0.022448 | SLC4A10 | 0.000108 | 0.007806 |
| LOC645682 | 0.000108 | 0.007806 | GYPA | 0.00474 | 0.042105 |
| GPD1L | 0.003232 | 0.034676 | CTNNA1 | 0.003232 | 0.034676 |
| IRF2BP1 | 0.00474 | 0.042105 | FRYL | 0.002047 | 0.027424 |
| TSHZ2 | 0.00474 | 0.042105 | OTUD7A | 0.001293 | 0.022448 |
| LIME1 | 0.00474 | 0.042105 | KIAA0565 | 0.000215 | 0.010104 |
| MGC34800 | 0.002047 | 0.027424 | TBC1D19 | 0.002047 | 0.027424 |
| C16orf55 | 0.002047 | 0.027424 | LASS4 | 0.00474 | 0.042105 |
| TMEM106B | 0.000108 | 0.007806 | UBA52 | 0.000431 | 0.012946 |
| EXT1 | 0.000108 | 0.007806 | PYY2 | 0.001293 | 0.022448 |
| SNX14 | 0.004262 | 0.042105 | PACAP | 0.001293 | 0.022448 |
| FLJ11021 | 0.000431 | 0.012946 | PDHA1 | 0.003232 | 0.034676 |
| C2orf3 | 0.000431 | 0.012946 | MLL | 0.000431 | 0.012946 |
| SMOC1 | 0.000431 | 0.012946 | LOC653472 | 0.000215 | 0.010104 |
| SMAD7 | 0.001293 | 0.022448 | DAP | 0.000108 | 0.007806 |
| LOC653103 | 0.003232 | 0.034676 | AGPAT1 | 0.000215 | 0.010104 |
| PHF19 | 0.000754 | 0.017438 | MTP18 | 0.002047 | 0.027424 |
| NR0B1 | 0.001293 | 0.022448 | LOC644337 | 0.001293 | 0.022448 |
| DICER1 | 0.002047 | 0.027424 | TMEM19 | 0.000431 | 0.012946 |
| MICAL-L1 | 0.002047 | 0.027424 | INSM1 | 0.000431 | 0.012946 |
| CHD9 | 0.001293 | 0.022448 | MYO5A | 0.000215 | 0.010104 |
| CYB5A | 0.003232 | 0.034676 | CHMP4B | 0.00474 | 0.042105 |
| DKFZp761I2123 | 0.003232 | 0.034676 | MRPS5 | 0.001225 | 0.022448 |
| TXNRD2 | 0.001293 | 0.022448 | POU3F3 | 0.000108 | 0.007806 |
| HNRPH2 | 0.000108 | 0.007806 | CERK | 0.002047 | 0.027424 |
| C1orf80 | 0.003232 | 0.034676 | NSMCE1 | 0.00474 | 0.042105 |
| ZDHHC14 | 0.002047 | 0.027424 | CXXC1 | 0.000108 | 0.007806 |
| DQX1 | 0.000754 | 0.017438 | NTS | 0.000215 | 0.010104 |
| ANKRD53 | 0.000754 | 0.017438 | SPARCL1 | 0.001293 | 0.022448 |
| CBX5 | 0.001293 | 0.022448 | LGI1 | 0.00474 | 0.042105 |
| KIAA0553 | 0.002047 | 0.027424 | TCEAL7 | 0.000215 | 0.010104 |
| PSRC1 | 0.001293 | 0.022448 | SLC26A9 | 0.001293 | 0.022448 |
| ARG1 | 0.003232 | 0.034676 | LOC652536 | 0.000754 | 0.017438 |
| TULP3 | 0.003232 | 0.034676 | RBM7 | 0.00474 | 0.042105 |
| APRIN | 0.003232 | 0.034676 | CBS | 0.000754 | 0.017438 |
| PPP1R9B | 0.003232 | 0.034676 | SH3BGRL | 0.000108 | 0.007806 |
| RB1 | 0.000108 | 0.007806 | C17orf44 | 0.002047 | 0.027424 |
| ZAN | 0.003232 | 0.034676 | ZNF31 | 0.002047 | 0.027424 |
| C18orf25 | 0.003232 | 0.034676 | PRR3 | 0.00474 | 0.042105 |
| SUMF1 | 0.000431 | 0.012946 | SLC7A2 | 0.001293 | 0.022448 |
| MAPRE1 | 0.000108 | 0.007806 | YWHAH | 0.002047 | 0.027424 |
| MLLT10 | 0.000431 | 0.012946 | LGALS7 | 0.002047 | 0.027424 |
| KIAA1754L | 0.000754 | 0.017438 | ART3 | 0.000431 | 0.012946 |
| RAD23A | 0.000431 | 0.012946 | BMP8A | 0.002047 | 0.027424 |
| UBE2Q2 | 0.003232 | 0.034676 | FLJ39370 | 0.003674 | 0.039253 |
| C9orf18 | 0.000754 | 0.017438 | BSG | 0.000431 | 0.012946 |
| CD63 | 0.002047 | 0.027424 | TLK2 | 0.000108 | 0.007806 |
| CDX4 | 0.003232 | 0.034676 | TSC22D1 | 0.003232 | 0.034676 |
| IL20RA | 0.00474 | 0.042105 | KIFC1 | 0.000754 | 0.017438 |
| OMD | 0.003232 | 0.034676 | GNB2L1 | 0.00474 | 0.042105 |
| AHNAK | 0.003232 | 0.034676 | PHGDH | 0.002047 | 0.027424 |
| SEMG1 | 0.001293 | 0.022448 | PROZ | 0.001293 | 0.022448 |
| BARX1 | 0.002047 | 0.027424 | TNRC6B | 0.000108 | 0.007806 |
| SCCPDH | 0.003232 | 0.034676 | SMOX | 0.00474 | 0.042105 |
| EHBP1 | 0.000108 | 0.007806 | CILP | 0.003232 | 0.034676 |
| ORC6L | 0.003232 | 0.034676 | UBR2 | 0.00474 | 0.042105 |
| LOC388397 | 0.000431 | 0.012946 | LOC652740 | 0.001293 | 0.022448 |
| ZC3H7A | 0.000431 | 0.012946 | LRIG2 | 0.000754 | 0.017438 |
| EVI5 | 0.001694 | 0.027424 | C19orf31 | 0.002047 | 0.027424 |
| RAB6IP2 | 0.003232 | 0.034676 | POLS | 0.000215 | 0.010104 |
| C21orf66 | 0.002047 | 0.027424 | CTNND1 | 0.003232 | 0.034676 |
| IGBP1 | 0.003232 | 0.034676 | LMBRD1 | 0.001293 | 0.022448 |
| FTCD | 0.000215 | 0.010104 | C7orf10 | 0.000754 | 0.017438 |
| MAT1A | 0.000754 | 0.017438 | CCDC50 | 0.004262 | 0.042105 |
| IMPAD1 | 0.001293 | 0.022448 | ANUBL1 | 0.001293 | 0.022448 |
| CART1 | 0.000108 | 0.007806 | CCNB2 | 0.002047 | 0.027424 |
| GSPT1 | 0.000108 | 0.007806 | ITLN2 | 0.000431 | 0.012946 |
| NOTCH2NL | 0.003232 | 0.034676 | RP4-691N24.1 | 0.00474 | 0.042105 |
| KLHL17 | 0.000754 | 0.017438 | LZTFL1 | 0.00474 | 0.042105 |
| LIX1 | 0.00474 | 0.042105 | TAF3 | 0.001442 | 0.024998 |
| REP15 | 0.003232 | 0.034676 | MIR16 | 0.002323 | 0.0309 |
| OSBP | 0.001694 | 0.027424 | RAB11B | 0.003232 | 0.034676 |
| PTK9 | 0.001293 | 0.022448 | GOLGA8B | 0.002047 | 0.027424 |
| UBE2I | 0.000754 | 0.017438 | UNQ689 | 0.002047 | 0.027424 |
| LRP10 | 0.000108 | 0.007806 | HSMPP8 | 0.001293 | 0.022448 |
| RORB | 0.000215 | 0.010104 | ELA3B | 0.000108 | 0.007806 |
| DGCR8 | 0.002047 | 0.027424 | NUDC | 0.000108 | 0.007806 |
| C17orf85 | 0.00474 | 0.042105 | HSPD1 | 0.003232 | 0.034676 |
| GLTSCR1 | 0.00474 | 0.042105 | THRAP1 | 0.000879 | 0.019929 |
| DIP2B | 0.000108 | 0.007806 | C12orf22 | 0.000754 | 0.017438 |
| LOC285958 | 0.000754 | 0.017438 | KIAA1919 | 0.001293 | 0.022448 |
| ATP11B | 0.001293 | 0.022448 | PRR7 | 0.003232 | 0.034676 |
| LPP | 0.000754 | 0.017438 | HOXA11 | 0.001293 | 0.022448 |
| UNC13C | 0.000215 | 0.010104 | TAF1L | 0.000108 | 0.007806 |
| C6orf25 | 0.003232 | 0.034676 | ZNF140 | 0.001293 | 0.022448 |
| SPANXD | 0.000215 | 0.010104 | CDC2L1 | 0.00474 | 0.042105 |
| ST3GAL6 | 0.001225 | 0.022448 | GNL1 | 0.000431 | 0.012946 |
| CT45-6 | 0.003232 | 0.034676 | C1orf61 | 0.000431 | 0.012946 |
| BTN3A1 | 0.000215 | 0.010104 | MSH5 | 0.002047 | 0.027424 |
| CPEB3 | 0.00474 | 0.042105 | MMP14 | 0.000431 | 0.012946 |
| GOLPH4 | 0.000215 | 0.010104 | LOC146325 | 0.002047 | 0.027424 |
| TBN | 0.000108 | 0.007806 | FLJ36492 | 0.000754 | 0.017438 |
| LARS | 0.000108 | 0.007806 | DPF1 | 0.000215 | 0.010104 |
| PNPLA2 | 0.003232 | 0.034676 | ADAMTS7 | 0.003232 | 0.034676 |
| ATRX | 0.000215 | 0.010104 | CBX1 | 0.000431 | 0.012946 |
| DYRK1B | 0.00474 | 0.042105 | LOC51057 | 0.000754 | 0.017438 |
| ASAM | 0.002047 | 0.027424 | LOC339344 | 0.003232 | 0.034676 |
| SYCP3 | 0.000754 | 0.017438 | TMEM35 | 0.001293 | 0.022448 |
| ANKRD43 | 0.001293 | 0.022448 | GSTP1 | 0.000431 | 0.012946 |
| C12orf29 | 0.001293 | 0.022448 | RPS4Y1 | 0.000754 | 0.017438 |
| ANKHD1 | 0.00474 | 0.042105 | LOC644380 | 0.000215 | 0.010104 |
| C15orf20 | 0.000108 | 0.007806 | LARP4 | 0.000431 | 0.012946 |
| KIAA0507 | 0.002047 | 0.027424 | SLC38A1 | 0.00474 | 0.042105 |
| CCDC75 | 0.003232 | 0.034676 | LETM2 | 0.003232 | 0.034676 |
| RABGGTB | 0.000754 | 0.017438 | BIRC4 | 0.000431 | 0.012946 |
| C20orf108 | 0.002047 | 0.027424 | BRWD1 | 0.003232 | 0.034676 |
| TIMELESS | 0.000108 | 0.007806 | MUC5AC | 0.000215 | 0.010104 |
| CLINT1 | 0.001293 | 0.022448 | DYDC2 | 0.00474 | 0.042105 |
| HMGN1 | 0.000754 | 0.017438 | ARFIP1 | 0.000431 | 0.012946 |
| LOC653125 | 0.000754 | 0.017438 | MGC42105 | 0.000108 | 0.007806 |
| RAXL1 | 0.00474 | 0.042105 | TMEM46 | 0.003232 | 0.034676 |
| PHOSPHO2 | 0.00474 | 0.042105 | LOC339778 | 0.000108 | 0.007806 |
| PRND | 0.00474 | 0.042105 | GLIS2 | 0.002047 | 0.027424 |
| C15orf51 | 0.000431 | 0.012946 | CDH24 | 0.001293 | 0.022448 |
| FAM26B | 0.00474 | 0.042105 | LAMA2 | 0.00474 | 0.042105 |
| NPM2 | 0.00474 | 0.042105 | TRAF7 | 0.001293 | 0.022448 |
| FAM60A | 0.00474 | 0.042105 | C14orf145 | 0.001293 | 0.022448 |
| HNRPA3P1 | 0.000215 | 0.010104 | C6orf85 | 0.00474 | 0.042105 |
| C8orf53 | 0.00474 | 0.042105 | MARCKSL1 | 0.00474 | 0.042105 |
| SETDB2 | 0.00474 | 0.042105 | CDH15 | 0.00474 | 0.042105 |
| GPR20 | 0.000431 | 0.012946 | CYorf15A | 0.000431 | 0.012946 |
| DPPA2 | 0.000108 | 0.007806 | MIP | 0.002047 | 0.027424 |
| WDR79 | 0.000108 | 0.007806 | LIF | 0.00474 | 0.042105 |
| OXSR1 | 0.000431 | 0.012946 | MUTED | 0.002047 | 0.027424 |
| LAT | 0.00474 | 0.042105 | SHOC2 | 0.000215 | 0.010104 |
| ERCC8 | 0.00474 | 0.042105 | IVL | 0.002047 | 0.027424 |
| TRIM35 | 0.003232 | 0.034676 | P2RX4 | 0.000215 | 0.010104 |
| LOC653805 | 0.000215 | 0.010104 | GABRB1 | 0.000108 | 0.007806 |
| HIPK1 | 0.000108 | 0.007806 | LOC652683 | 0.000431 | 0.012946 |
| CBARA1 | 0.000215 | 0.010104 | CTCFL | 0.000215 | 0.010104 |
| AHI1 | 0.000754 | 0.017438 | TAF2 | 0.000431 | 0.012946 |
| HHIP | 0.002047 | 0.027424 | LOC653789 | 0.000431 | 0.012946 |
| MBL2 | 0.000215 | 0.010104 | C17orf51 | 0.003232 | 0.034676 |
| LMX1B | 0.000431 | 0.012946 | MOSC2 | 0.003232 | 0.034676 |
| RNF165 | 0.000754 | 0.017438 | FALZ | 0.001293 | 0.022448 |
| POLR2C | 0.000431 | 0.012946 | MYH10 | 0.000215 | 0.010104 |
| ZCCHC7 | 0.00474 | 0.042105 | CXCR3 | 0.002047 | 0.027424 |
| FLJ20464 | 0.000215 | 0.010104 | AKT1S1 | 0.000431 | 0.012946 |
| SUGT1 | 0.000879 | 0.019929 | GPR135 | 0.002047 | 0.027424 |
| C8orf16 | 0.003232 | 0.034676 | TEX15 | 0.00474 | 0.042105 |
| BTBD14A | 0.00474 | 0.042105 | FCN3 | 0.000108 | 0.007806 |
| NRP1 | 0.000215 | 0.010104 | LSM12 | 0.000431 | 0.012946 |
| ZFPM1 | 0.00474 | 0.042105 | UPP1 | 0.000431 | 0.012946 |
| LOC653349 | 0.003232 | 0.034676 | VCL | 0.002047 | 0.027424 |
| NHLH1 | 0.000754 | 0.017438 | C10orf57 | 0.003232 | 0.034676 |
| UPF3A | 0.00474 | 0.042105 | FAM119B | 0.000431 | 0.012946 |
| FRS3 | 0.002047 | 0.027424 | LRP16 | 0.000108 | 0.007806 |
| FLJ32549 | 0.00474 | 0.042105 | LMAN1 | 0.002047 | 0.027424 |
| RGS14 | 0.000215 | 0.010104 | CLK2P | 0.003232 | 0.034676 |
| SPINK5L3 | 0.000431 | 0.012946 | LOC641814 | 0.000431 | 0.012946 |
| FLJ10154 | 0.001293 | 0.022448 | HOXD10 | 0.001293 | 0.022448 |
| C3orf51 | 0.001293 | 0.022448 | DNAJB14 | 0.00474 | 0.042105 |
| UBE2C | 0.001293 | 0.022448 | VPREB1 | 0.001225 | 0.022448 |
| C1orf113 | 0.003232 | 0.034676 | LOC90113 | 0.001225 | 0.022448 |
| NFKBIL1 | 0.000754 | 0.017438 | SFRS15 | 0.003232 | 0.034676 |
| PGBD4 | 0.000431 | 0.012946 | SMARCC1 | 0.004262 | 0.042105 |
| PYGB | 0.003232 | 0.034676 | SYT3 | 0.00474 | 0.042105 |
| SOX1 | 0.003232 | 0.034676 | ACSM3 | 0.003232 | 0.034676 |
| RTN1 | 0.002047 | 0.027424 | AFAR3 | 0.00474 | 0.042105 |
| VEPH1 | 0.000108 | 0.007806 | MED10 | 0.002047 | 0.027424 |
| TBC1D22B | 0.000108 | 0.007806 | RPA4 | 0.002047 | 0.027424 |
| RIPK3 | 0.000431 | 0.012946 | FOLR3 | 0.000754 | 0.017438 |
| MGC21644 | 0.002047 | 0.027424 | ANKRD54 | 0.001293 | 0.022448 |
| FOXM1 | 0.000108 | 0.007806 | RSN | 0.000108 | 0.007806 |
| EXOSC9 | 0.003232 | 0.034676 | RPS4Y2 | 0.002047 | 0.027424 |
| SNX19 | 0.000108 | 0.007806 | COVA1 | 0.000108 | 0.007806 |
| S100PBP | 0.001293 | 0.022448 | TTL | 0.000108 | 0.007806 |
| GMEB1 | 0.001225 | 0.022448 | TYW3 | 0.000431 | 0.012946 |
| GPR78 | 0.002047 | 0.027424 | FLJ43276 | 0.000874 | 0.019929 |
| SERPINB4 | 0.003232 | 0.034676 | AGPAT4 | 0.001293 | 0.022448 |
| CLSPN | 0.000215 | 0.010104 | C15orf41 | 0.003232 | 0.034676 |
| C6orf15 | 0.000108 | 0.007806 | GABRR2 | 0.00474 | 0.042105 |
| RAB11A | 0.001293 | 0.022448 | WIPI2 | 0.003232 | 0.034676 |
| SCNN1D | 0.002047 | 0.027424 | ATG4A | 0.00474 | 0.042105 |
| SAC3D1 | 0.001293 | 0.022448 | ZCCHC6 | 0.000754 | 0.017438 |
| C6orf151 | 0.000215 | 0.010104 | CCDC120 | 0.003232 | 0.034676 |
| CCNF | 0.000431 | 0.012946 | TAF1C | 0.000215 | 0.010104 |
| NDRG4 | 0.002047 | 0.027424 | PAK1IP1 | 0.000108 | 0.007806 |
| LOC646264 | 0.002047 | 0.027424 | AGTR1 | 0.000431 | 0.012946 |
| PIGH | 0.004936 | 0.043629 | POLR3C | 0.000754 | 0.017438 |
| CD74 | 0.002047 | 0.027424 | TINP1 | 0.002047 | 0.027424 |
| FLJ21657 | 0.002047 | 0.027424 | LOC652565 | 0.000108 | 0.007806 |
| C14orf129 | 0.000215 | 0.010104 | METTL3 | 0.00474 | 0.042105 |
| PRDX2 | 0.00474 | 0.042105 | CTRL | 0.00474 | 0.042105 |
| KIAA0446 | 0.002047 | 0.027424 | VISA | 0.002047 | 0.027424 |
| LOC402176 | 0.003232 | 0.034676 | CXYorf2 | 0.003232 | 0.034676 |
| GJC1 | 0.001293 | 0.022448 | TRIM26 | 0.000754 | 0.017438 |
| EGFR | 0.003232 | 0.034676 | NBEAL1 | 0.00474 | 0.042105 |
| REEP5 | 0.003232 | 0.034676 | RASGRP1 | 0.00474 | 0.042105 |
| NFIC | 0.000754 | 0.017438 | CC2D1B | 0.002047 | 0.027424 |
| PCDHB4 | 0.003232 | 0.034676 | LOC389199 | 0.000754 | 0.017438 |
| DDX25 | 0.000879 | 0.019929 | KPNA6 | 0.000431 | 0.012946 |
| PLAGL2 | 0.001293 | 0.022448 | SYNGR4 | 0.003232 | 0.034676 |
| MITF | 0.001293 | 0.022448 | NSUN5C | 0.000754 | 0.017438 |
| TGOLN2 | 0.000108 | 0.007806 | FOXA1 | 0.000431 | 0.012946 |
| CENPO | 0.000874 | 0.019929 | MYBL1 | 0.000431 | 0.012946 |
| NOTCH2 | 0.000215 | 0.010104 | LOC648979 | 0.000754 | 0.017438 |
| SSFA2 | 0.000108 | 0.007806 | SERPINA7 | 0.000754 | 0.017438 |
| PVALB | 0.002047 | 0.027424 | LOC440595 | 0.004262 | 0.042105 |
| SLC29A3 | 0.000754 | 0.017438 | GTF2A1 | 0.000108 | 0.007806 |
| ORC1L | 0.003232 | 0.034676 | EFTUD1 | 0.001986 | 0.027424 |
| RIPK1 | 0.000108 | 0.007806 | LOC283755 | 0.003232 | 0.034676 |
| G3BP2 | 0.000108 | 0.007806 | C6orf106 | 0.000108 | 0.007806 |
| LOC201175 | 0.002047 | 0.027424 | AASS | 0.00474 | 0.042105 |
| MAFA | 0.00474 | 0.042105 | STXBP5L | 0.000108 | 0.007806 |
| NLF2 | 0.00474 | 0.042105 | UTF1 | 0.001293 | 0.022448 |
| GTPBP3 | 0.001293 | 0.022448 | ST8SIA2 | 0.001293 | 0.022448 |
| DNAJB11 | 0.000754 | 0.017438 | PTPN11 | 0.000108 | 0.007806 |
| WBSCR23 | 0.00474 | 0.042105 | IGF2BP3 | 0.000431 | 0.012946 |
| ZBTB11 | 0.003232 | 0.034676 | LOC653596 | 0.002047 | 0.027424 |
| CPE | 0.002047 | 0.027424 | FEM1C | 0.000108 | 0.007806 |
| PIP5K1C | 0.002047 | 0.027424 | SNURF | 0.00474 | 0.042105 |
| DKK4 | 0.003674 | 0.039253 | LIMD1 | 0.000431 | 0.012946 |
| SOX21 | 0.000431 | 0.012946 | DKK2 | 0.000431 | 0.012946 |
| MGC5509 | 0.000108 | 0.007806 | ODAM | 0.000215 | 0.010104 |
| PRPF40A | 0.001293 | 0.022448 | OSBPL3 | 0.00474 | 0.042105 |
| POLD1 | 0.000754 | 0.017438 | ZRANB2 | 0.000108 | 0.007806 |
| APOB | 0.00474 | 0.042105 | ABCC4 | 0.003232 | 0.034676 |
| MVK | 0.000215 | 0.010104 | MYH3 | 0.00474 | 0.042105 |
| ARL13B | 0.000215 | 0.010104 | OTOR | 0.001293 | 0.022448 |
| NFE2 | 0.002047 | 0.027424 | MGC10334 | 0.00474 | 0.042105 |
| RNU17D | 0.003232 | 0.034676 | SFRS4 | 0.000108 | 0.007806 |
| FBXO34 | 0.00474 | 0.042105 | CLOCK | 0.001293 | 0.022448 |
| USP33 | 0.000754 | 0.017438 | RNF111 | 0.000754 | 0.017438 |
| NAPA | 0.000754 | 0.017438 | SMEK2 | 0.003232 | 0.034676 |
| TTC28 | 0.002047 | 0.027424 | CD2BP2 | 0.001694 | 0.027424 |
| TMEM44 | 0.00474 | 0.042105 | BCL2 | 0.000215 | 0.010104 |
| ANKS6 | 0.000431 | 0.012946 | CD1B | 0.002047 | 0.027424 |
| PCNT | 0.000108 | 0.007806 | PLCG1 | 0.000431 | 0.012946 |
| MAGEA10 | 0.000215 | 0.010104 | JSRP1 | 0.000754 | 0.017438 |
| LOC642209 | 0.00474 | 0.042105 | GUCA1B | 0.000108 | 0.007806 |
| ZNF318 | 0.00474 | 0.042105 | RPL10 | 0.00474 | 0.042105 |
| USP19 | 0.001293 | 0.022448 | C14orf24 | 0.000431 | 0.012946 |
| BLNK | 0.00474 | 0.042105 | GPR55 | 0.003232 | 0.034676 |
| STX1A | 0.002047 | 0.027424 | KL | 0.002047 | 0.027424 |
| ELMO3 | 0.00474 | 0.042105 | BCLAF1 | 0.000108 | 0.007806 |
| NPY | 0.000431 | 0.012946 | SPTB | 0.003232 | 0.034676 |
| C17orf50 | 0.002047 | 0.027424 | IPO13 | 0.000431 | 0.012946 |
| ZBED1 | 0.000754 | 0.017438 | AHCYL1 | 0.00474 | 0.042105 |
| DNAJC18 | 0.003232 | 0.034676 | CCNG2 | 0.004936 | 0.043629 |
| FAT2 | 0.002047 | 0.027424 | ZBTB8OS | 0.00474 | 0.042105 |
| IGF2BP2 | 0.000108 | 0.007806 | MAGI2 | 0.002047 | 0.027424 |
| NEDD8 | 0.002047 | 0.027424 | CASC5 | 0.00474 | 0.042105 |
| LOC653551 | 0.002047 | 0.027424 | PARN | 0.000215 | 0.010104 |
| CTA-216E10.6 | 0.002047 | 0.027424 | PAMCI | 0.002047 | 0.027424 |
| KIN | 0.000431 | 0.012946 | UBE2N | 0.002047 | 0.027424 |
| ZNF594 | 0.002047 | 0.027424 | MGC39584 | 0.00474 | 0.042105 |
| PRIM2A | 0.000754 | 0.017438 | PTDSS2 | 0.003674 | 0.039253 |
| CSF1R | 0.000754 | 0.017438 | ACBD4 | 0.001293 | 0.022448 |
| FMN2 | 0.000431 | 0.012946 | CENPN | 0.003232 | 0.034676 |
| LOC645296 | 0.002047 | 0.027424 | ATXN10 | 0.000215 | 0.010104 |
| GABARAP | 0.000754 | 0.017438 | RAN | 0.000754 | 0.017438 |
| FAM24A | 0.000754 | 0.017438 | UBE2G1 | 0.000879 | 0.019929 |
| SNAPAP | 0.001293 | 0.022448 | PTPLB | 0.00474 | 0.042105 |
| WDR75 | 0.00316 | 0.034676 | RASSF4 | 0.000754 | 0.017438 |
| SSX4B | 0.000215 | 0.010104 | LOC126661 | 0.00474 | 0.042105 |
| AP2B1 | 0.00316 | 0.034676 | PSMA2 | 0.000431 | 0.012946 |
| LOC387753 | 0.001293 | 0.022448 | NUDT12 | 0.003232 | 0.034676 |
| MGC2749 | 0.000108 | 0.007806 | UCP3 | 0.003232 | 0.034676 |
| CTGLF1 | 0.003232 | 0.034676 | RNPEP | 0.000431 | 0.012946 |
| C3orf40 | 0.002047 | 0.027424 | KIAA0528 | 0.003232 | 0.034676 |
| PSAT1 | 0.000108 | 0.007806 | HOXD11 | 0.001293 | 0.022448 |
| POLR2J2 | 0.001293 | 0.022448 | STARD5 | 0.002047 | 0.027424 |
| HM13 | 0.000215 | 0.010104 | C6orf170 | 0.002047 | 0.027424 |
| LOC57149 | 0.000215 | 0.010104 | C3orf19 | 0.000215 | 0.010104 |
| THRAP3 | 0.000108 | 0.007806 | ZNF206 | 0.003232 | 0.034676 |
| KIAA1212 | 0.000215 | 0.010104 | AARSL | 0.001293 | 0.022448 |
| GAPVD1 | 0.002047 | 0.027424 | LAX1 | 0.00474 | 0.042105 |
| ZNF483 | 0.003232 | 0.034676 | RAB6B | 0.002047 | 0.027424 |
| DDX24 | 0.003232 | 0.034676 | CAPN1 | 0.002047 | 0.027424 |
| CCDC64 | 0.003232 | 0.034676 | DEAF1 | 0.000754 | 0.017438 |
| LOC255783 | 0.002047 | 0.027424 | MEIS2 | 0.003232 | 0.034676 |
| UBE2O | 0.003232 | 0.034676 | YWHAG | 0.002047 | 0.027424 |
| LEMD3 | 0.00474 | 0.042105 | SGPL1 | 0.002047 | 0.027424 |
| HMGB1 | 0.003232 | 0.034676 | SLC6A12 | 0.003232 | 0.034676 |
| RGS20 | 0.000754 | 0.017438 | DSCAM | 0.004262 | 0.042105 |
| LOC653577 | 0.002323 | 0.0309 | HDAC8 | 0.002047 | 0.027424 |
| KIAA1109 | 0.000431 | 0.012946 | FADS1 | 0.003232 | 0.034676 |
| SPANXD | 0.001293 | 0.022448 | RAVER1 | 0.00474 | 0.042105 |
| JMJD1B | 0.000431 | 0.012946 | C6orf208 | 0.000431 | 0.012946 |
| TRA2A | 0.00474 | 0.042105 | MYST2 | 0.000431 | 0.012946 |
| PIP5K1A | 0.002047 | 0.027424 | CDC2L1 | 0.000431 | 0.012946 |
| GJB3 | 0.001293 | 0.022448 | LOC93349 | 0.000754 | 0.017438 |
| TAC3 | 0.00474 | 0.042105 | ATF4 | 0.000431 | 0.012946 |
| SLC16A8 | 0.00474 | 0.042105 | FBXL3 | 0.000108 | 0.007806 |
| SETD8 | 0.000431 | 0.012946 | SCARF2 | 0.00474 | 0.042105 |
| MCTP2 | 0.000754 | 0.017438 | POLR3G | 0.000108 | 0.007806 |
| DUSP10 | 0.000215 | 0.010104 | CBLN1 | 0.001293 | 0.022448 |
| CCDC93 | 0.00474 | 0.042105 | FBXO4 | 0.002047 | 0.027424 |
| ABI3 | 0.002047 | 0.027424 | FTSJ3 | 0.003232 | 0.034676 |
| CST6 | 0.00474 | 0.042105 | C20orf134 | 0.003232 | 0.034676 |
| ACOXL | 0.002047 | 0.027424 | TMEM62 | 0.000215 | 0.010104 |
| TSPY1 | 0.000108 | 0.007806 | CHUK | 0.002047 | 0.027424 |
| ASPN | 0.00474 | 0.042105 | CALCR | 0.00474 | 0.042105 |
| DCAKD | 0.001293 | 0.022448 | MYT1 | 0.003232 | 0.034676 |
| WNT4 | 0.001293 | 0.022448 | VIL1 | 0.000108 | 0.007806 |
| NBPF4 | 0.000879 | 0.019929 | PDCL3 | 0.000754 | 0.017438 |
| SOX18 | 0.002047 | 0.027424 | WNT10A | 0.00474 | 0.042105 |
| SUSD4 | 0.00474 | 0.042105 | ZNF581 | 0.00474 | 0.042105 |
| LOC645296 | 0.00474 | 0.042105 | H2AFB1 | 0.00474 | 0.042105 |
| LRRFIP2 | 0.002047 | 0.027424 | LOC648303 | 0.000215 | 0.010104 |
| ADAMTS13 | 0.003232 | 0.034676 | SDAD1 | 0.000108 | 0.007806 |
| PNPLA6 | 0.001293 | 0.022448 | IER5 | 0.002047 | 0.027424 |
| ITSN2 | 0.000215 | 0.010104 | ZNF28 | 0.002047 | 0.027424 |
| NAP1L1 | 0.002047 | 0.027424 | MUM1 | 0.003232 | 0.034676 |
| LOC132321 | 0.003232 | 0.034676 | LOC348262 | 0.002047 | 0.027424 |
| CDC40 | 0.000108 | 0.007806 | FAM63A | 0.000431 | 0.012946 |
| TBC1D23 | 0.000108 | 0.007806 | LXN | 0.002047 | 0.027424 |
| BTN2A2 | 0.000754 | 0.017438 | CENPM | 0.003232 | 0.034676 |
| BTG3 | 0.002047 | 0.027424 | F2 | 0.004913 | 0.04362 |
| ENTPD1 | 0.001293 | 0.022448 | DACH1 | 0.000108 | 0.007806 |
| OR4K17 | 0.003232 | 0.034676 | CDC20 | 0.000754 | 0.017438 |
| PPIG | 0.000108 | 0.007806 | KISS1R | 0.003232 | 0.034676 |
| NR2C2 | 0.000754 | 0.017438 | VPS4B | 0.001293 | 0.022448 |
| DKFZp762E1312 | 0.001225 | 0.022448 | ZNF419 | 0.001293 | 0.022448 |
| TOE1 | 0.003232 | 0.034676 | VCX-C | 0.00474 | 0.042105 |
| PLK4 | 0.001293 | 0.022448 | RET | 0.000754 | 0.017438 |
| C22orf24 | 0.000431 | 0.012946 | KIAA0040 | 0.000215 | 0.010104 |
| C1orf21 | 0.001293 | 0.022448 | UBE2V1 | 0.001293 | 0.022448 |
| TAF4 | 0.00474 | 0.042105 | FADS2 | 0.00474 | 0.042105 |
| CEBPA | 0.001293 | 0.022448 | KIAA1545 | 0.003232 | 0.034676 |
| RKHD1 | 0.00474 | 0.042105 | MMEL1 | 0.000108 | 0.007806 |
| WHDC1L1 | 0.000108 | 0.007806 | TMEM153 | 0.002047 | 0.027424 |
| FBP1 | 0.00474 | 0.042105 | ARHGAP22 | 0.000754 | 0.017438 |
| PRAMEF18 | 0.000431 | 0.012946 | CHD6 | 0.003232 | 0.034676 |
| LOC644362 | 0.002047 | 0.027424 | EP400NL | 0.002047 | 0.027424 |
| APOE | 0.000754 | 0.017438 | GNB1L | 0.000754 | 0.017438 |
| STX6 | 0.002047 | 0.027424 | SLC25A24 | 0.00474 | 0.042105 |
| GH2 | 0.000754 | 0.017438 | MACF1 | 0.000431 | 0.012946 |
| LOC653193 | 0.00474 | 0.042105 | FXYD2 | 0.003144 | 0.034676 |
| OTUD3 | 0.001293 | 0.022448 | C5orf24 | 0.001293 | 0.022448 |
| YARS | 0.001293 | 0.022448 | PPM1K | 0.001293 | 0.022448 |
| EPB41L2 | 0.002047 | 0.027424 | PPIE | 0.000431 | 0.012946 |
| C16orf57 | 0.002047 | 0.027424 | ZNF273 | 0.001293 | 0.022448 |
| ARPC5 | 0.000108 | 0.007806 | NAPG | 0.000108 | 0.007806 |
| CHRM3 | 0.00474 | 0.042105 | LEPREL1 | 0.000108 | 0.007806 |
| FLJ12949 | 0.00474 | 0.042105 | FXR2 | 0.000431 | 0.012946 |
| HLXB9 | 0.00474 | 0.042105 | ITGB1BP2 | 0.001293 | 0.022448 |
| BRWD3 | 0.000215 | 0.010104 | TBL1XR1 | 0.001293 | 0.022448 |
| C9orf156 | 0.003232 | 0.034676 | TAF1 | 0.001293 | 0.022448 |
| MUM1L1 | 0.002047 | 0.027424 | KCNQ3 | 0.002047 | 0.027424 |
| LOC644951 | 0.00474 | 0.042105 | OR2A25 | 0.002047 | 0.027424 |
| ASAH1 | 0.000108 | 0.007806 | GLT25D2 | 0.000431 | 0.012946 |
| STIP1 | 0.00474 | 0.042105 | CRR9 | 0.00474 | 0.042105 |
| SCRT2 | 0.002047 | 0.027424 | ZNF682 | 0.00474 | 0.042105 |
| XRCC5 | 0.001293 | 0.022448 | FAM98A | 0.00474 | 0.042105 |
| SERPINH1 | 0.000754 | 0.017438 | PRUNE | 0.000215 | 0.010104 |
| CCL20 | 0.003232 | 0.034676 | RAD51 | 0.001293 | 0.022448 |
| CYP4F3 | 0.000754 | 0.017438 | GSTM2 | 0.002047 | 0.027424 |
| EIF2S3 | 0.003232 | 0.034676 | SOX13 | 0.000108 | 0.007806 |
| MTMR12 | 0.003232 | 0.034676 | TPM1 | 0.000754 | 0.017438 |
| C12orf30 | 0.002047 | 0.027424 | BBS1 | 0.003232 | 0.034676 |
| DTNBP1 | 0.002047 | 0.027424 | CTSO | 0.000215 | 0.010104 |
| APCDD1L | 0.000431 | 0.012946 | SLC6A6 | 0.001225 | 0.022448 |
| FPGT | 0.000431 | 0.012946 | LOC653622 | 0.003232 | 0.034676 |
| SYNPO | 0.003232 | 0.034676 | MED18 | 0.000108 | 0.007806 |
| LOC648293 | 0.002047 | 0.027424 | DKFZP434A0131 | 0.001293 | 0.022448 |
| TXLNA | 0.003674 | 0.039253 | LOC653773 | 0.00474 | 0.042105 |
| PTPRT | 0.002323 | 0.0309 | MKL2 | 0.00474 | 0.042105 |
| MGMT | 0.003232 | 0.034676 | HMGA2 | 0.000108 | 0.007806 |
| KRTAP13-4 | 0.000754 | 0.017438 | PANK1 | 0.000215 | 0.010104 |
| CYP24A1 | 0.000431 | 0.012946 | KRT222P | 0.003232 | 0.034676 |
| EPO | 0.000754 | 0.017438 | XPA | 0.001293 | 0.022448 |
| KIAA1012 | 0.001293 | 0.022448 | PCBP1 | 0.003232 | 0.034676 |
| LOC644354 | 0.002047 | 0.027424 | VAPB | 0.001293 | 0.022448 |
| CALU | 0.002047 | 0.027424 | TMEM69 | 0.00474 | 0.042105 |
| RPL39 | 0.002047 | 0.027424 | RUFY2 | 0.00474 | 0.042105 |
| C14orf109 | 0.002047 | 0.027424 | SMARCE1 | 0.00474 | 0.042105 |
| HEMGN | 0.000108 | 0.007806 | DCXR | 0.000215 | 0.010104 |
| ASCC3 | 0.000108 | 0.007806 | DLGAP3 | 0.00474 | 0.042105 |
| SLC7A13 | 0.000108 | 0.007806 | LMBR1 | 0.001293 | 0.022448 |
| TYROBP | 0.003232 | 0.034676 | CEP63 | 0.00474 | 0.042105 |
| TRAF3 | 0.00474 | 0.042105 | PASD1 | 0.001293 | 0.022448 |
| MALAT1 | 0.00474 | 0.042105 | TGFB1I1 | 0.000431 | 0.012946 |
| MAB21L2 | 0.001293 | 0.022448 | KIAA0196 | 0.000754 | 0.017438 |
| KNG1 | 0.002047 | 0.027424 | GNA12 | 0.00474 | 0.042105 |
| PVT1 | 0.000431 | 0.012946 | BRD2 | 0.003232 | 0.034676 |
| LOC653125 | 0.002047 | 0.027424 | TRHDE | 0.00474 | 0.042105 |
| MGC16597 | 0.000108 | 0.007806 | DOC1 | 0.00474 | 0.042105 |
| ASGR1 | 0.000431 | 0.012946 | BBC3 | 0.003232 | 0.034676 |
| IL1RAP | 0.000215 | 0.010104 | SLC40A1 | 0.003232 | 0.034676 |
| HES7 | 0.00474 | 0.042105 | PNPLA4 | 0.000754 | 0.017438 |
| MMP17 | 0.002047 | 0.027424 | MGC87631 | 0.000754 | 0.017438 |
| LOC389517 | 0.002047 | 0.027424 | ZNF626 | 0.000754 | 0.017438 |
| SLC12A3 | 0.000431 | 0.012946 | CENPC1 | 0.000108 | 0.007806 |
| LOC347273 | 0.00474 | 0.042105 | ARHGAP29 | 0.000108 | 0.007806 |
| C14orf100 | 0.003232 | 0.034676 | TM9SF2 | 0.003232 | 0.034676 |
| INS | 0.00474 | 0.042105 | FBXO11 | 0.000754 | 0.017438 |
| PAPD1 | 0.000215 | 0.010104 | MCM3AP | 0.004936 | 0.043629 |
| LOC389906 | 0.00474 | 0.042105 | EIF2AK2 | 0.003232 | 0.034676 |
| SUMO1 | 0.000754 | 0.017438 | HSZFP36 | 0.00474 | 0.042105 |
| MGC16384 | 0.003232 | 0.034676 | PRKAG1 | 0.000754 | 0.017438 |
| NRG4 | 0.000431 | 0.012946 | ZNF507 | 0.000754 | 0.017438 |
| LOC641738 | 0.000108 | 0.007806 | CDC42BPB | 0.00474 | 0.042105 |
| PSME1 | 0.00474 | 0.042105 | DYNLRB2 | 0.000431 | 0.012946 |
| SBK1 | 0.002047 | 0.027424 | COL7A1 | 0.002047 | 0.027424 |
| ERAL1 | 0.004262 | 0.042105 | SPAG1 | 0.002047 | 0.027424 |
| LGALS1 | 0.002047 | 0.027424 | INHBE | 0.000108 | 0.007806 |
| SFXN3 | 0.00474 | 0.042105 | FOXL2 | 0.00474 | 0.042105 |
| CNTROB | 0.003232 | 0.034676 | CCNA2 | 0.003232 | 0.034676 |
| BZW1 | 0.001293 | 0.022448 | TLOC1 | 0.002047 | 0.027424 |
| KCNJ13 | 0.000431 | 0.012946 | MEG3 | 0.00474 | 0.042105 |
| PRDM4 | 0.001293 | 0.022448 | POU5F1 | 0.000215 | 0.010104 |
| RAB11FIP1 | 0.000431 | 0.012946 | PRSS2 | 0.000108 | 0.007806 |
| MPHOSPH10 | 0.000108 | 0.007806 | MINPP1 | 0.002047 | 0.027424 |
| KCNG1 | 0.000754 | 0.017438 | LOC284422 | 0.000215 | 0.010104 |
| SMARCA3 | 0.000431 | 0.012946 | MCC | 0.002047 | 0.027424 |
| CXorf23 | 0.001293 | 0.022448 | ACIN1 | 0.002047 | 0.027424 |
| CCNK | 0.000108 | 0.007806 | TES | 0.000431 | 0.012946 |
| TMTC3 | 0.000215 | 0.010104 | MGC40405 | 0.003232 | 0.034676 |
| RIPK4 | 0.00474 | 0.042105 | RAPGEF4 | 0.000431 | 0.012946 |
| CDH4 | 0.001293 | 0.022448 | RLN1 | 0.002047 | 0.027424 |
| NR2E1 | 0.000215 | 0.010104 | ZNF425 | 0.00474 | 0.042105 |
| C21orf91 | 0.003232 | 0.034676 | LARGE | 0.000215 | 0.010104 |
| C20orf59 | 0.001293 | 0.022448 | REXO4 | 0.000879 | 0.019929 |
| MICB | 0.000754 | 0.017438 | CDC5L | 0.000215 | 0.010104 |
| LRP1 | 0.00474 | 0.042105 | CARF | 0.00474 | 0.042105 |
| ZCCHC17 | 0.003232 | 0.034676 | DENND3 | 0.00474 | 0.042105 |
| ZNF641 | 0.000431 | 0.012946 | ZNF197 | 0.000754 | 0.017438 |
| EIF4B | 0.003232 | 0.034676 | ZNF2 | 0.003232 | 0.034676 |
| IFNAR2 | 0.000754 | 0.017438 | NOL7 | 0.003232 | 0.034676 |
| SURF6 | 0.000108 | 0.007806 | SEC23B | 0.000431 | 0.012946 |
| NKD2 | 0.003232 | 0.034676 | FAM11A | 0.000754 | 0.017438 |
| PRSS7 | 0.000108 | 0.007806 | RPL9 | 0.001293 | 0.022448 |
| OPRM1 | 0.003232 | 0.034676 | C1orf43 | 0.003656 | 0.039201 |
| POLH | 0.002047 | 0.027424 | OR8D4 | 0.003232 | 0.034676 |
| C9orf98 | 0.00474 | 0.042105 | SSBP2 | 0.001293 | 0.022448 |
| NFKBIZ | 0.002047 | 0.027424 | PTER | 0.003232 | 0.034676 |
| MGC45922 | 0.003232 | 0.034676 | CLEC4M | 0.003232 | 0.034676 |
| ZNF614 | 0.000431 | 0.012946 | SPANXB2 | 0.000754 | 0.017438 |
| CSPP1 | 0.00474 | 0.042105 | MAGEA3 | 0.00474 | 0.042105 |
| HDLBP | 0.000431 | 0.012946 | C3orf25 | 0.000215 | 0.010104 |
| TMEM66 | 0.000108 | 0.007806 | GCGR | 0.003232 | 0.034676 |
| LOC648294 | 0.003232 | 0.034676 | LOC645721 | 0.001293 | 0.022448 |
| SPANXA2 | 0.002047 | 0.027424 | LOC646576 | 0.000108 | 0.007806 |
| CDK6 | 0.000108 | 0.007806 | ALB | 0.000108 | 0.007806 |
| BRI3BP | 0.000754 | 0.017438 | NDUFAB1 | 0.003232 | 0.034676 |
| MDC1 | 0.000108 | 0.007806 | ZNF415 | 0.00474 | 0.042105 |
| ADAM23 | 0.000215 | 0.010104 | C10orf84 | 0.002047 | 0.027424 |
| FGF10 | 0.00474 | 0.042105 | CCDC109A | 0.002047 | 0.027424 |
| RASGEF1C | 0.00474 | 0.042105 | PTK2 | 0.002047 | 0.027424 |
| LSAMP | 0.000754 | 0.017438 | ST3GAL4 | 0.001293 | 0.022448 |
| FLJ11903 | 0.000108 | 0.007806 | FTS | 0.003232 | 0.034676 |
| USP20 | 0.003232 | 0.034676 | DNASE2B | 0.001293 | 0.022448 |
| PPARD | 0.002047 | 0.027424 | THAP1 | 0.00474 | 0.042105 |
| ARMC3 | 0.000215 | 0.010104 | MOAP1 | 0.003232 | 0.034676 |
| PRDM15 | 0.002047 | 0.027424 | DES | 0.00474 | 0.042105 |
| C15orf28 | 0.001293 | 0.022448 | TRPM7 | 0.000108 | 0.007806 |
| C8orf70 | 0.000754 | 0.017438 | KIAA0179 | 0.001293 | 0.022448 |
| PMS2CL | 0.000754 | 0.017438 | PYCR2 | 0.000431 | 0.012946 |
| UBE1L2 | 0.000108 | 0.007806 | DLX1 | 0.000215 | 0.010104 |
| LOC652226 | 0.00474 | 0.042105 | ATP8A2 | 0.002047 | 0.027424 |
| HES2 | 0.000431 | 0.012946 | FLJ21816 | 0.003232 | 0.034676 |
| KIAA0152 | 0.000108 | 0.007806 | LOC649279 | 0.003232 | 0.034676 |
| C2orf30 | 0.000431 | 0.012946 | C9orf37 | 0.000215 | 0.010104 |
| GSG1 | 0.001293 | 0.022448 | LOC645156 | 0.003232 | 0.034676 |
| MGC16121 | 0.003232 | 0.034676 | HIST1H2BA | 0.001293 | 0.022448 |
| MDFI | 0.00474 | 0.042105 | LOC648176 | 0.000431 | 0.012946 |
| RBM28 | 0.000215 | 0.010104 | CTSZ | 0.000215 | 0.010104 |
| SEC63 | 0.001293 | 0.022448 | F8A1 | 0.002047 | 0.027424 |
| LBH | 0.003232 | 0.034676 | GDEP | 0.002047 | 0.027424 |
| SRP46 | 0.000754 | 0.017438 | C18orf49 | 0.001293 | 0.022448 |
| SPTLC1 | 0.001293 | 0.022448 | CGGBP1 | 0.000431 | 0.012946 |
| ARL4A | 0.00474 | 0.042105 | CD1A | 0.001293 | 0.022448 |
| DIDO1 | 0.002047 | 0.027424 | ILDR1 | 0.003232 | 0.034676 |
| FLJ10781 | 0.003232 | 0.034676 | VPS52 | 0.000431 | 0.012946 |
| FOXC1 | 0.000754 | 0.017438 | C6orf211 | 0.000431 | 0.012946 |
| NEUROG1 | 0.002047 | 0.027424 | GALNT7 | 0.00474 | 0.042105 |
| CSPG4 | 0.003232 | 0.034676 | LRBA | 0.002047 | 0.027424 |
| IFNA4 | 0.000108 | 0.007806 | CD46 | 0.000108 | 0.007806 |
| CHEK2 | 0.000431 | 0.012946 | EIF2AK1 | 0.002047 | 0.027424 |
| SLC25A6 | 0.002047 | 0.027424 | VCX-C | 0.003232 | 0.034676 |
| DOCK3 | 0.002047 | 0.027424 | MYO6 | 0.003232 | 0.034676 |
| GALK2 | 0.000431 | 0.012946 | LOC388642 | 0.001293 | 0.022448 |
| RAB12 | 0.003232 | 0.034676 | VGLL2 | 0.002047 | 0.027424 |
| MYCNOS | 0.002047 | 0.027424 | ZNF70 | 0.000108 | 0.007806 |
| KBTBD3 | 0.00474 | 0.042105 | GDA | 0.000215 | 0.010104 |
| TMEM30A | 0.000754 | 0.017438 | TRABD | 0.003232 | 0.034676 |
| SUPT7L | 0.003232 | 0.034676 | SGOL1 | 0.000754 | 0.017438 |
| GRIN2D | 0.001293 | 0.022448 | Septin 2 | 0.004262 | 0.042105 |
| LOC645124 | 0.001293 | 0.022448 | LOC401904 | 0.00474 | 0.042105 |
| TPP1 | 0.00474 | 0.042105 | MRAS | 0.00474 | 0.042105 |
| CCDC81 | 0.000215 | 0.010104 | JTB | 0.002047 | 0.027424 |
| MGC12966 | 0.002047 | 0.027424 | OR8G1 | 0.000431 | 0.012946 |
| PARP10 | 0.003232 | 0.034676 | STK33 | 0.003232 | 0.034676 |
| BIN1 | 0.001293 | 0.022448 | PDAP1 | 0.000108 | 0.007806 |
| CREB3L2 | 0.002047 | 0.027424 | UACA | 0.000754 | 0.017438 |
| NPAS3 | 0.002047 | 0.027424 | SLFN5 | 0.003232 | 0.034676 |
| FGF1 | 0.00474 | 0.042105 | ELA2 | 0.000431 | 0.012946 |
| MGC10701 | 0.001293 | 0.022448 | ROBO1 | 0.000108 | 0.007806 |
| CXorf15 | 0.000108 | 0.007806 | ZBTB39 | 0.000754 | 0.017438 |
| SET | 0.001293 | 0.022448 | RNASET2 | 0.000754 | 0.017438 |
| TWIST1 | 0.00474 | 0.042105 | HRK | 0.001293 | 0.022448 |
| UIP1 | 0.000754 | 0.017438 | OSTbeta | 0.002047 | 0.027424 |
| DOCK5 | 0.000108 | 0.007806 | SP3 | 0.001293 | 0.022448 |
| CNN2 | 0.000754 | 0.017438 | FLJ22709 | 0.000215 | 0.010104 |
| LOC653048 | 0.000108 | 0.007806 | ROS1 | 0.000431 | 0.012946 |
| TLN2 | 0.003232 | 0.034676 | TAF9B | 0.00474 | 0.042105 |
| TAX1BP3 | 0.000754 | 0.017438 | SCN3A | 0.00474 | 0.042105 |
| RSRC1 | 0.000215 | 0.010104 | MRPL21 | 0.00474 | 0.042105 |
| RHOBTB1 | 0.001293 | 0.022448 | PODXL2 | 0.000431 | 0.012946 |
| CA5B | 0.003232 | 0.034676 | TAPBP | 0.000754 | 0.017438 |
| C3orf21 | 0.000108 | 0.007806 | SKIV2L | 0.001293 | 0.022448 |
| KCNH6 | 0.002047 | 0.027424 | COMMD3 | 0.000431 | 0.012946 |
| PSEN1 | 0.000108 | 0.007806 | SEMA3C | 0.002047 | 0.027424 |
| ATF6 | 0.000754 | 0.017438 | TACC3 | 0.000108 | 0.007806 |
| DNALI1 | 0.003232 | 0.034676 | L1TD1 | 0.000108 | 0.007806 |
| DDX3Y | 0.00474 | 0.042105 | C22orf26 | 0.000215 | 0.010104 |
| ATG4D | 0.00474 | 0.042105 | KIAA0251 | 0.00474 | 0.042105 |
| TNPO1 | 0.002047 | 0.027424 | POLR3A | 0.002047 | 0.027424 |
| TCOF1 | 0.003232 | 0.034676 | AGMAT | 0.000108 | 0.007806 |
| PRKAB2 | 0.003232 | 0.034676 | RPL23AP13 | 0.00474 | 0.042105 |
| RIMS3 | 0.000431 | 0.012946 | CENPB | 0.00474 | 0.042105 |
| ACOT2 | 0.002047 | 0.027424 | PPP1R12A | 0.003232 | 0.034676 |
| FOXD3 | 0.003232 | 0.034676 | ARPC2 | 0.000108 | 0.007806 |
| C20orf151 | 0.000215 | 0.010104 | C14orf115 | 0.000108 | 0.007806 |
| WDR40B | 0.003232 | 0.034676 | C9orf85 | 0.003232 | 0.034676 |
| C1orf34 | 0.00474 | 0.042105 | BTBD7 | 0.000215 | 0.010104 |
| GRIN2C | 0.000754 | 0.017438 | HTRA3 | 0.000215 | 0.010104 |
| DIAPH3 | 0.000754 | 0.017438 | GJA7 | 0.000108 | 0.007806 |
| NFAT5 | 0.001293 | 0.022448 | MYO18B | 0.00474 | 0.042105 |
| LOC654351 | 0.000431 | 0.012946 | RHOA | 0.003232 | 0.034676 |
| MKI67 | 0.003232 | 0.034676 | ZNF613 | 0.002047 | 0.027424 |
| MXD1 | 0.000108 | 0.007806 | TADA1L | 0.000108 | 0.007806 |
| KIAA1539 | 0.000431 | 0.012946 | BAGE | 0.000215 | 0.010104 |
| TEGT | 0.003232 | 0.034676 | TCEAL5 | 0.002047 | 0.027424 |
| IGBP1 | 0.000431 | 0.012946 | TNPO2 | 0.00474 | 0.042105 |
| ZNF585A | 0.003232 | 0.034676 | LOC116143 | 0.000754 | 0.017438 |
| IRAK3 | 0.002047 | 0.027424 | KBTBD7 | 0.003232 | 0.034676 |
| PCSK1N | 0.002047 | 0.027424 | ZNF575 | 0.000754 | 0.017438 |
| SULT2A1 | 0.000108 | 0.007806 | WDR63 | 0.000108 | 0.007806 |
| RIF1 | 0.002323 | 0.0309 | SNAPC1 | 0.000215 | 0.010104 |
| PLOD1 | 0.000215 | 0.010104 | RBM3 | 0.003232 | 0.034676 |
| LOC647474 | 0.000108 | 0.007806 | RAPGEFL1 | 0.002047 | 0.027424 |
| PRM2 | 0.00474 | 0.042105 | C19orf22 | 0.000431 | 0.012946 |
| NR1H4 | 0.000108 | 0.007806 | SCUBE2 | 0.001293 | 0.022448 |
| MTHFD1L | 0.000108 | 0.007806 | LOC201895 | 0.000108 | 0.007806 |
| HPS1 | 0.00474 | 0.042105 | ZZZ3 | 0.002047 | 0.027424 |
| S100A2 | 0.001293 | 0.022448 | KIAA1370 | 0.000754 | 0.017438 |
| NPBWR2 | 0.000431 | 0.012946 | LOC644037 | 0.000431 | 0.012946 |
| SRCRB4D | 0.002047 | 0.027424 | NAT2 | 0.000754 | 0.017438 |
| SIDT1 | 0.000215 | 0.010104 | HSPC268 | 0.000754 | 0.017438 |
| YPEL5 | 0.003232 | 0.034676 | ALCAM | 0.000108 | 0.007806 |
| BHLHB4 | 0.003232 | 0.034676 | C12orf51 | 0.000431 | 0.012946 |
| ZIC5 | 0.003232 | 0.034676 | CLDN10 | 0.00474 | 0.042105 |
| FLJ21736 | 0.001293 | 0.022448 | FANCE | 0.002047 | 0.027424 |
| RBM9 | 0.001293 | 0.022448 | RAB10 | 0.000754 | 0.017438 |
| ABLIM2 | 0.003232 | 0.034676 | LOC253842 | 0.001293 | 0.022448 |
| PAGE5 | 0.001293 | 0.022448 | PEX11G | 0.003232 | 0.034676 |
| SUCLA2 | 0.00474 | 0.042105 | S100A3 | 0.00474 | 0.042105 |
| LCE5A | 0.003232 | 0.034676 | ZNF701 | 0.000754 | 0.017438 |
| FAM74A1 | 0.000431 | 0.012946 | REEP3 | 0.000431 | 0.012946 |
| KIFC3 | 0.002047 | 0.027424 | KIAA0828 | 0.00474 | 0.042105 |
| GALNT1 | 0.002047 | 0.027424 | LOC643426 | 0.000108 | 0.007806 |
| CD24 | 0.003232 | 0.034676 | CXorf43 | 0.000215 | 0.010104 |
| CCDC18 | 0.004262 | 0.042105 | RPL15 | 0.002047 | 0.027424 |
| GCN1L1 | 0.003232 | 0.034676 | MPP5 | 0.00474 | 0.042105 |
| OS9 | 0.000431 | 0.012946 | HBG2 | 0.002047 | 0.027424 |
| TUSC1 | 0.001293 | 0.022448 | GLUD2 | 0.000754 | 0.017438 |
| SLC41A3 | 0.000431 | 0.012946 | EDG2 | 0.001293 | 0.022448 |
| PPAPDC1B | 0.003232 | 0.034676 | RARB | 0.00474 | 0.042105 |
| FLJ30277 | 0.00474 | 0.042105 | STK32A | 0.001293 | 0.022448 |
| GAGE10 | 0.000431 | 0.012946 | ACSL3 | 0.00474 | 0.042105 |
| EML4 | 0.000431 | 0.012946 | FXR1 | 0.00474 | 0.042105 |
| LOC644380 | 0.000215 | 0.010104 | RAPGEF5 | 0.000431 | 0.012946 |
| CUGBP1 | 0.000431 | 0.012946 | CORIN | 0.003232 | 0.034676 |
| FLJ36874 | 0.000108 | 0.007806 | BAZ1A | 0.000754 | 0.017438 |
| SLC30A10 | 0.001293 | 0.022448 | LOC137886 | 0.001293 | 0.022448 |
| SDK1 | 0.000754 | 0.017438 | HNRPR | 0.000215 | 0.010104 |
| PTPLAD2 | 0.002047 | 0.027424 | SLC19A2 | 0.000431 | 0.012946 |
| UCK1 | 0.003232 | 0.034676 | SAFB | 0.004936 | 0.043629 |
| NT5C | 0.002047 | 0.027424 | CCDC98 | 0.002047 | 0.027424 |
| LARP5 | 0.002047 | 0.027424 | LMTK3 | 0.00474 | 0.042105 |
| PLA2G12B | 0.000108 | 0.007806 | OR7D2 | 0.001293 | 0.022448 |
| C2orf28 | 0.002047 | 0.027424 | WDR46 | 0.003232 | 0.034676 |
| ZNF571 | 0.001293 | 0.022448 | RBM25 | 0.00474 | 0.042105 |
| UBXD3 | 0.000754 | 0.017438 | C21orf93 | 0.00474 | 0.042105 |
| AMD1 | 0.000215 | 0.010104 | VARSL | 0.002047 | 0.027424 |
| LOC126208 | 0.00474 | 0.042105 | CASP8 | 0.000431 | 0.012946 |
| HSP90AA1 | 0.000431 | 0.012946 | VGF | 0.00474 | 0.042105 |
| RHBDL1 | 0.00474 | 0.042105 | FAM9C | 0.00474 | 0.042105 |
| PRPF38B | 0.000108 | 0.007806 | SOCS6 | 0.00474 | 0.042105 |
| NCF1 | 0.001293 | 0.022448 | TTC5 | 0.003232 | 0.034676 |
| RCOR1 | 0.000431 | 0.012946 | ZFAND2B | 0.000108 | 0.007806 |
| CECR2 | 0.00474 | 0.042105 | FLJ39660 | 0.000108 | 0.007806 |
| CUEDC1 | 0.001293 | 0.022448 | ZNF702 | 0.00474 | 0.042105 |
| SIX6 | 0.000108 | 0.007806 | ASL | 0.000754 | 0.017438 |
| OPRS1 | 0.00474 | 0.042105 | DNMT3B | 0.003232 | 0.034676 |
| MIPEP | 0.00474 | 0.042105 | SLC30A3 | 0.00474 | 0.042105 |
| WDR85 | 0.003232 | 0.034676 | C4B | 0.001293 | 0.022448 |
| HS2ST1 | 0.00474 | 0.042105 | ATF7IP | 0.000754 | 0.017438 |
| MATN4 | 0.000431 | 0.012946 | GINS4 | 0.000108 | 0.007806 |
| DYNC1H1 | 0.002047 | 0.027424 | MRPL4 | 0.00316 | 0.034676 |
| HOMER1 | 0.001293 | 0.022448 | CCDC115 | 0.002047 | 0.027424 |
| TBC1D9 | 0.000215 | 0.010104 | TA-NFKBH | 0.00474 | 0.042105 |
| CRTC2 | 0.000754 | 0.017438 | RPP38 | 0.003232 | 0.034676 |
| LOC653751 | 0.002047 | 0.027424 | MAGEH1 | 0.003232 | 0.034676 |
| ATAD3C | 0.00474 | 0.042105 | RANBP1 | 0.000215 | 0.010104 |
| PQLC3 | 0.002047 | 0.027424 | DDX6 | 0.002047 | 0.027424 |
| C17orf58 | 0.000431 | 0.012946 | RNASE9 | 0.001293 | 0.022448 |
| ARNTL2 | 0.000108 | 0.007806 | CYFIP1 | 0.002047 | 0.027424 |
| GUSBL2 | 0.001293 | 0.022448 | LOC644617 | 0.000431 | 0.012946 |
| ATCAY | 0.001293 | 0.022448 | THRAP5 | 0.000879 | 0.019929 |
| XTP3TPA | 0.00474 | 0.042105 | CSAG2 | 0.00474 | 0.042105 |
| FLJ38663 | 0.002047 | 0.027424 | ZNF441 | 0.002047 | 0.027424 |
| SUPT3H | 0.001293 | 0.022448 | LOC653067 | 0.000108 | 0.007806 |
| NEUROG3 | 0.000754 | 0.017438 | SRR | 0.000108 | 0.007806 |
| PPP1R8 | 0.000431 | 0.012946 | PWP2H | 0.001293 | 0.022448 |
| KIF14 | 0.00474 | 0.042105 | ERCC1 | 0.000431 | 0.012946 |
| INPP4A | 0.000431 | 0.012946 | FKSG2 | 0.003232 | 0.034676 |
| TNRC15 | 0.000108 | 0.007806 | NACA | 0.000215 | 0.010104 |
| UTP14A | 0.000108 | 0.007806 | C1orf56 | 0.00474 | 0.042105 |
| DHX16 | 0.000431 | 0.012946 | C20orf26 | 0.000431 | 0.012946 |
| PTCD1 | 0.000431 | 0.012946 | PSCD3 | 0.000754 | 0.017438 |
| PTPN18 | 0.000754 | 0.017438 | C1QDC1 | 0.00474 | 0.042105 |
| ANP32E | 0.000108 | 0.007806 | SLC16A14 | 0.002047 | 0.027424 |
| C1GALT1 | 0.000431 | 0.012946 | TNFRSF8 | 0.002047 | 0.027424 |
| ZNF74 | 0.001293 | 0.022448 | STAU1 | 0.001293 | 0.022448 |
| HP1BP3 | 0.000879 | 0.019929 | NASP | 0.000215 | 0.010104 |
| PDE7A | 0.001293 | 0.022448 | FLJ39575 | 0.001293 | 0.022448 |
| ABCG5 | 0.000431 | 0.012946 | PNMA1 | 0.00474 | 0.042105 |
| SPANXD | 0.000754 | 0.017438 | NALP11 | 0.002047 | 0.027424 |
| CACNA1E | 0.00474 | 0.042105 | LOC642826 | 0.003232 | 0.034676 |
| UPF3B | 0.000108 | 0.007806 | SH2D5 | 0.003232 | 0.034676 |
| C20orf6 | 0.000754 | 0.017438 | RPL10L | 0.000754 | 0.017438 |
| TTC9B | 0.002047 | 0.027424 | C19orf21 | 0.003232 | 0.034676 |
| STAT2 | 0.000754 | 0.017438 | ZMAT5 | 0.003674 | 0.039253 |
| CDKAL1 | 0.000108 | 0.007806 | CX3CR1 | 0.001293 | 0.022448 |
| DDC | 0.002047 | 0.027424 | CHCHD5 | 0.004936 | 0.043629 |
| NRAS | 0.00474 | 0.042105 | SLC15A4 | 0.003232 | 0.034676 |
| RBM22 | 0.00474 | 0.042105 | SEMA3F | 0.00474 | 0.042105 |
| PCSK1 | 0.003232 | 0.034676 | GRPEL1 | 0.000108 | 0.007806 |
| ZNF597 | 0.000431 | 0.012946 | IPP | 0.000431 | 0.012946 |
| SLCO1B1 | 0.000108 | 0.007806 | SYNGR2 | 0.003232 | 0.034676 |
| KLK15 | 0.001293 | 0.022448 | C1orf93 | 0.00474 | 0.042105 |
| UGT2A3 | 0.000215 | 0.010104 | C12orf24 | 0.001293 | 0.022448 |
| RPN2 | 0.00474 | 0.042105 | ARTN | 0.00474 | 0.042105 |
| C1orf149 | 0.000215 | 0.010104 | DHX9 | 0.002047 | 0.027424 |
| RFC1 | 0.000215 | 0.010104 | NTN4 | 0.000431 | 0.012946 |
| MTMR1 | 0.002323 | 0.0309 | NKIRAS2 | 0.00474 | 0.042105 |
| KCNJ14 | 0.002047 | 0.027424 | AP3B1 | 0.002047 | 0.027424 |
| TRAF3IP1 | 0.00474 | 0.042105 | ZNF709 | 0.002047 | 0.027424 |
| PDCD6 | 0.00474 | 0.042105 | ANKRD1 | 0.000108 | 0.007806 |
| KCND2 | 0.000108 | 0.007806 | NDUFA10 | 0.00474 | 0.042105 |
| ITPKA | 0.000431 | 0.012946 | EIF4EBP3 | 0.002047 | 0.027424 |
| NARG2 | 0.000431 | 0.012946 | GPR160 | 0.000879 | 0.019929 |
| DOCK7 | 0.000108 | 0.007806 | ZNF691 | 0.001293 | 0.022448 |
| MAPK14 | 0.000215 | 0.010104 | C12orf56 | 0.00474 | 0.042105 |
| C1orf50 | 0.00474 | 0.042105 | PCNP | 0.002047 | 0.027424 |
| CDC42EP3 | 0.000431 | 0.012946 | GNA13 | 0.000215 | 0.010104 |
| C1orf163 | 0.00474 | 0.042105 | FLJ25801 | 0.00474 | 0.042105 |
| C20orf30 | 0.003232 | 0.034676 | BHLHB2 | 0.002047 | 0.027424 |
| SMN1 | 0.002047 | 0.027424 | UBE2R2 | 0.000108 | 0.007806 |
| PAPOLA | 0.000108 | 0.007806 | GLRB | 0.002047 | 0.027424 |
| FNDC6 | 0.000431 | 0.012946 | SH3KBP1 | 0.002047 | 0.027424 |
| MAGEA1 | 0.000108 | 0.007806 | MAN2B2 | 0.002047 | 0.027424 |
| LOC375133 | 0.003232 | 0.034676 | PPARA | 0.000108 | 0.007806 |
| FIP1L1 | 0.00474 | 0.042105 | MID1IP1 | 0.001293 | 0.022448 |
| NFKBIL2 | 0.000431 | 0.012946 | CNIH | 0.003232 | 0.034676 |
| HBEGF | 0.001293 | 0.022448 | PSRC2 | 0.000108 | 0.007806 |
| LOC644676 | 0.002047 | 0.027424 | EFHD1 | 0.00474 | 0.042105 |
| SMPX | 0.000431 | 0.012946 | C21orf99 | 0.003232 | 0.034676 |
| MYLK2 | 0.003232 | 0.034676 | PRKAR1B | 0.001293 | 0.022448 |
| C1orf33 | 0.000108 | 0.007806 | SMAD4 | 0.000108 | 0.007806 |
| EIF4EBP1 | 0.003232 | 0.034676 | RKHD2 | 0.000754 | 0.017438 |
| MSL2L1 | 0.00474 | 0.042105 | FYCO1 | 0.000215 | 0.010104 |
| ZNF525 | 0.002047 | 0.027424 | LOH11CR2A | 0.000108 | 0.007806 |
| UBR1 | 0.002047 | 0.027424 | ARNTL | 0.002047 | 0.027424 |
| SP8 | 0.000108 | 0.007806 | AKR1D1 | 0.00474 | 0.042105 |
| TYRP1 | 0.000754 | 0.017438 | LRRC4C | 0.000754 | 0.017438 |
| ITIH2 | 0.00474 | 0.042105 | LYAR | 0.000754 | 0.017438 |
| GPR156 | 0.002047 | 0.027424 | C9orf68 | 0.002047 | 0.027424 |
| UBE2S | 0.003232 | 0.034676 | LOC388621 | 0.001694 | 0.027424 |
| NSD1 | 0.000215 | 0.010104 | MGC17330 | 0.00474 | 0.042105 |
| FGFR1OP2 | 0.001293 | 0.022448 | PSMC6 | 0.00474 | 0.042105 |
| ANKRD24 | 0.000108 | 0.007806 | LOC647054 | 0.003232 | 0.034676 |
| WDR67 | 0.000108 | 0.007806 | ZFPM2 | 0.000108 | 0.007806 |
| TAS2R14 | 0.003232 | 0.034676 | C4orf18 | 0.001293 | 0.022448 |
| C6orf120 | 0.000754 | 0.017438 | DYNC1I2 | 0.001694 | 0.027424 |
| LOC649407 | 0.003232 | 0.034676 | ATP5A1 | 0.000215 | 0.010104 |
| WDR7 | 0.002047 | 0.027424 | CAB39 | 0.003232 | 0.034676 |
| BDP1 | 0.000431 | 0.012946 | ASZ1 | 0.000431 | 0.012946 |
| IGHMBP2 | 0.00474 | 0.042105 | OGN | 0.002047 | 0.027424 |
| RIOK1 | 0.002323 | 0.0309 | GLUD1 | 0.001293 | 0.022448 |
| ANKRD13D | 0.000431 | 0.012946 | RHOC | 0.000108 | 0.007806 |
| CCDC86 | 0.001293 | 0.022448 | TH1L | 0.000215 | 0.010104 |
| CCDC77 | 0.003232 | 0.034676 | LOC652683 | 0.000431 | 0.012946 |
| ARMET | 0.00474 | 0.042105 | KRT77 | 0.002047 | 0.027424 |
| MSN | 0.000108 | 0.007806 | KIAA1737 | 0.000754 | 0.017438 |
| CBWD6 | 0.000431 | 0.012946 | PJCG6 | 0.000431 | 0.012946 |
| CD8B | 0.000754 | 0.017438 | RASSF8 | 0.001225 | 0.022448 |
| KLHL9 | 0.003232 | 0.034676 | NDRG1 | 0.001694 | 0.027424 |
| RNF103 | 0.00474 | 0.042105 | DLGAP4 | 0.001293 | 0.022448 |
| YRDC | 0.002047 | 0.027424 | DC-UbP | 0.000108 | 0.007806 |

**Table S5 Detailed description of 12 genes in MIRS related to tumorigenesis or carcinogenesis.**

| **Gene ID** | **Annotation** |
| --- | --- |
| **APOA5** | Encode an apolipoprotein that plays an important role in regulating the plasma triglyceride levels. |
| **FAM9C** | This gene is a member of a gene family which arose through duplication on the X chromosome., which plays anti-apoptotic role through activation of the PI3K/Akt pathway |
| **IVL** | IVL is one of the precursor proteins of the cornified cell envelope and is markedly increased in inflammatory skin diseases. |
| **PAGE5** | This gene is expressed in a variety of tumors and in some fetal and reproductive tissues, and implicated in programmed cell death |
| **CACNA1E** | Voltage-dependent calcium channels, involved in a variety of calcium-dependent processes, including muscle contraction, hormone or neurotransmitter release, gene expression, cell motility, cell division and cell death |
| **CCL25** | This gene promotes the migration and invasion |
| **CD1A** | mediate the presentation of primarily lipid and glycolipid antigens of self or microbial origin to T cells |
| **CD1B** | mediate the presentation of primarily lipid and glycolipid antigens of self or microbial origin to T cells |
| **GPR55** | G-protein-coupled receptor superfamily, May be involved in hyperalgesia associated with inflammatory and neuropathic pain. |
| **LAX1** | Lymphocyte Transmembrane Adaptor 1, protein kinase binding and SH2 domain binding, Negatively regulates TCR (T-cell antigen receptor)-mediated signaling in T-cells and BCR (B-cell antigen receptor)-mediated signaling in B-cells |
| **TNFRSF8** | A member of the TNF-receptor superfamily. This receptor is expressed by activated, but not by resting, T and B cells |
| **WNT10A** | implicated in oncogenesis and in several developmental processes, including regulation of cell fate and patterning during embryogenesis |
